# Cardiovascular Signatures of COVID-19 Predict Mortality and Identify Barrier Stabilizing Therapies

**DOI:** 10.1101/2022.02.08.22270636

**Authors:** Dakota Gustafson, Michelle Ngai, Ruilin Wu, Huayun Hou, Alice Schoffel, Clara Erice, Serena Mandla, Filio Billia, Michael D. Wilson, Milica Radisic, Eddy Fan, Uriel Trahtemberg, Andrew Baker, Chris McIntosh, Chun-Po S. Fan, Claudia C. dos Santos, Kevin C. Kain, Kate Hanneman, Paaladinesh Thavendiranathan, Jason E. Fish, Kathryn L. Howe

## Abstract

**Background:** Endothelial cell (EC) activation, endotheliitis, vascular permeability, and thrombosis have been observed in patients with severe COVID-19, indicating that the vasculature is affected during the acute stages of SARS-CoV-2 infection. It remains unknown whether circulating vascular markers are sufficient to predict clinical outcomes, are unique to COVID-19, and if vascular permeability can be therapeutically targeted.

**Methods:** Evaluating the prevalence of circulating inflammatory, cardiac and EC activation markers, and the development of a microRNA atlas in 241 patients with suspected SARS-CoV-2 infection, allowed their prognostic value to be assessed by a Random Forest model machine learning approach. Subsequent ex vivo experiments assessed EC permeability responses to patient plasma and were used to uncover modulated gene regulatory networks from which rational therapeutic design was inferred.

**Findings:** Multiple inflammatory and EC activation biomarkers were associated with mortality in COVID-19 patients and in severity-matched SARS-CoV-2-negative patients, while dysregulation of specific microRNAs at presentation was specific for poor COVID-19-related outcomes and revealed disease-relevant pathways. Integrating the datasets using a machine learning approach further enhanced clinical risk prediction for in-hospital mortality. Exposure of ECs to COVID-19 patient plasma resulted in severity-specific gene expression responses and EC barrier dysfunction which was ameliorated using angiopoietin-1 mimetic or recombinant Slit2-N.

**Interpretation:** Integration of multi-omics data identified microRNA and vascular biomarkers prognostic of in-hospital mortality in COVID-19 patients and revealed that vascular stabilizing therapies should be explored as a treatment for endothelial dysfunction in COVID-19, and other severe diseases where endothelial dysfunction has a central role in pathogenesis.

**RESEARCH IN CONTEXT:** *Evidence before this study:* While diagnostic testing has allowed for the rapid identification of COVID-19 cases, the lack of post-diagnosis risk assessment metrics, especially among the highest-risk subgroups, thereby undermined the cascade and allocation of care. To date, the integration of clinical data with broad omics technologies has opened up new avenues for efficiently delineating complex patient phenotypes and their associations with clinical outcomes, with circulating profiles of plasma microRNAs (miRNA), in particular, having been shown to be tightly associated with disease, and capable of providing not only detailed prognostic information but also mechanistic insight.

*Added value of this study:* Markers of endothelial dysfunction at presentation, while indicative of poor outcomes in COVID-19-positive patients, likely reflect systemic vascular dysfunction in critically ill patients and are not specific to SARS-CoV-2 infection. More so, the generation of a plasma microRNA atlas uncovers COVID-19-specific prognostic markers and multiple disease-specific pathways of interest, including endothelial barrier dysfunction. Furthermore, synthesis of electronic health record data with clinically relevant multi-omic datasets using a machine learning approach provides substantially better metrics by which mortality can be estimated in patients with severe COVID-19. Finally, targeted stabilization of the endothelial barrier with Q-Peptide and Slit2-N are novel therapeutic avenues that should be explored in COVID-19 patients.

*Implications of all the available evidence:* Together, our work provides biological insight into the role of the endothelium in SARS-CoV-2 infection, the importance of miRNA as disease- and pathway-specific biomarkers, and the exciting possibility that endothelial barrier stabilizing treatments might hold promise in COVID-19.

## INTRODUCTION

Severe acute respiratory syndrome coronavirus 2 (SARS-CoV-2) is a highly contagious betacoronavirus, which results in coronavirus disease 2019 (COVID-19).(1) While the majority of infected individuals manifest mild to moderate illness, 14-31% of symptomatic unvaccinated patients eventually require hospitalization, with intensive care unit (ICU) admission rates ranging from 2-26% among those hospitalized.(2) Select populations, particularly older individuals and those with underlying comorbidities (including cardiovascular disease [CVD]) have high rates of morbidity and mortality.(3) Substantially higher rates of morbidity and mortality have been observed as variants of concern become more predominant.(4) While diagnostic testing has allowed for the rapid identification of COVID-19 cases, the lack of post-diagnosis risk assessment metrics, especially among the highest-risk subgroups, have undermined the cascade and allocation of care.

It has been proposed that the use of existing cardiovascular and respiratory parameters could serve as a metric of risk prediction.(5, 6) However, case-fatality rates of those with comorbidities remain particularly high (e.g., preexisting CVD at ∼10.5%) with cardiorespiratory the aforementioned markers having limited utility.(7) In this regard, while standard metrics including measures of cardiac damage (e.g., troponin values above the 99^th^ percentile), the extent of inflammatory activation (e.g., C-reactive protein expression), and cardiovascular imaging have elucidated the spectrum of COVID-19 complications, they have only modestly elucidated the risk of adverse in-hospital outcomes and often provide limited insight into disease mechanism.(8–10) Albeit encouraging, current COVID-19 therapeutics focus on stemming aberrant immune responses and controlling viral reproduction (e.g., tocilizumab and remdesevir, respectively), which may neglect key elements of the host response contributing to outcomes.(11) From this perspective, clinical data and autopsy studies revealing endotheliitis and thrombosis have raised the possibility that endothelial dysfunction, particularly fluctuations in vascular integrity and coagulative capacity, could be a driver of clinical outcomes.(12) Thrombosis, fluid extravasation, and microangiopathy observed in the small vessels and capillaries of the lungs directly support the notion that an intense vascular reaction takes place in those with severe disease.(13)

Early data suggested that viral tropism towards angiotensin-converting enzyme 2 and acetylated sialic acid residues, which are highly expressed by vascular endothelial cells (EC), could instigate cardiovascular dysfunction.(14–19) However, recent *in vitro* evidence has alternatively suggested that SARS-CoV-2 may have limited infectious potential and replicative ability in ECs.(20) In this respect, mechanisms secondary to direct infection, such as cascading immunological activation may instead be the driving factor behind the observed endothelial dysfunction; particularly among the populations with coexisting conditions where EC dysfunction is already evident.(21, 22) In fact, recent single cohort analysis of select endothelium-related biomarkers such as thrombomodulin(23), von Willebrand Factor(24), angiopoietin-2 (Ang-2)(25), and soluble triggering receptor expressed on myeloid cells-1 (sTREM-1)(26) have shown utility in prognostication, being associated with both disease severity and in-hospital mortality. While markers of endothelial function may aid in prognostication, it seems unlikely that a simple combination of markers can provide insight significant enough to adjudicate the level of care a patient will need, nor is it apparent that these markers associate specifically with COVID-19 pathology. To date, the integration of clinical data with broad omics technologies has opened up new avenues for efficiently delineating complex patient phenotypes and their associations with clinical outcomes.(27, 28) Circulating profiles of plasma microRNAs (miRNA), in particular, have been shown to be tightly associated with disease, and capable of providing not only detailed prognostic information but also mechanistic insight.(29, 30)

In this study, we sought to broadly characterize markers of endothelial function and inflammation and to develop for the first time an atlas of miRNA expression across both a spectrum of individuals diagnosed with COVID-19 as well as SARS-CoV-2-negative patients from the ICU. To enhance clinical utility, we focused on understanding how these markers are associated with in-hospital mortality of high-risk patients, particularly those requiring the highest levels of care.

## METHODS

### Contact for Reagent and Resource Sharing

The authors declare that all supporting data are available within the article and its supplementary data files. Transcriptomic data (i.e., messenger RNA and miRNA sequencing) have been deposited at the Gene Expression Omnibus and the RStudio analysis pipeline is contained within the Online Data Supplement. Further information and requests for resources, reagents, or patient-level datasets from qualified researchers trained in human subject confidentiality protocols may be sent to the lead contact, Dr. Kathryn Howe.

### Synopsis of Study Design, Patient Demographics, and Clinical Severity

Two hundred and forty-one patients with suspected, community-acquired SARS-CoV-2 (acute infection) were enrolled prospectively in the emergency departments or upon admission at two urban, quaternary-care hospitals in Toronto, Canada (University Health Network and St. Michael’s Hospital, Table 1 and Online Figure I) between May 2020 to December 2020 (prior to vaccine availability). Infection status of admitted patients was confirmed by at least two SARS-CoV-2 polymerase-chain reaction tests. Patients with SARS-CoV-2 but noninfectious etiologies were not enrolled (e.g., blunt force trauma). Medical history, physical examination, clinical laboratory values, and acute illness scores (Acute Physiologic Assessment and Chronic Health Evaluation II and Sequential Organ Failure Assessment) were recorded upon admission (day 0 or 1; t_0-1_), two to three days later (t_2-3_), and up to five days (t_4-5_), along with the synchronous collection of blood samples (Online Figure I). As a result of sampling logistics, t_0_ or t_1_ (days) were grouped as the earliest timepoint available for admitted patients. Similarly, collapsed sampling timepoints at t_2-3_ and t_4-5_ allowed evaluation of patient trajectories. Since the analysis was conducted retrospectively, clinical care was dictated by individual care providers with the primary outcome being mortality.

**Table 1.**
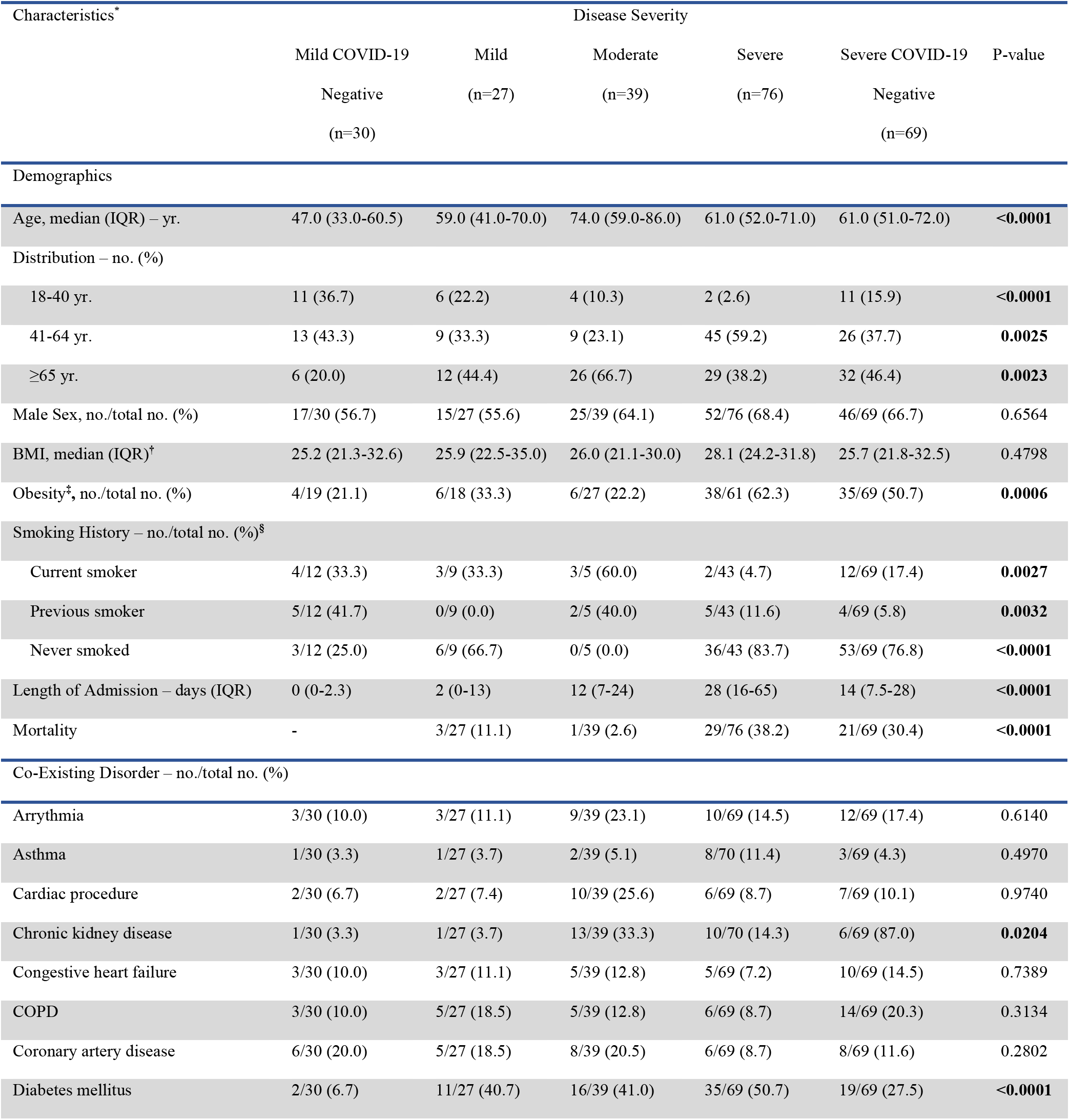
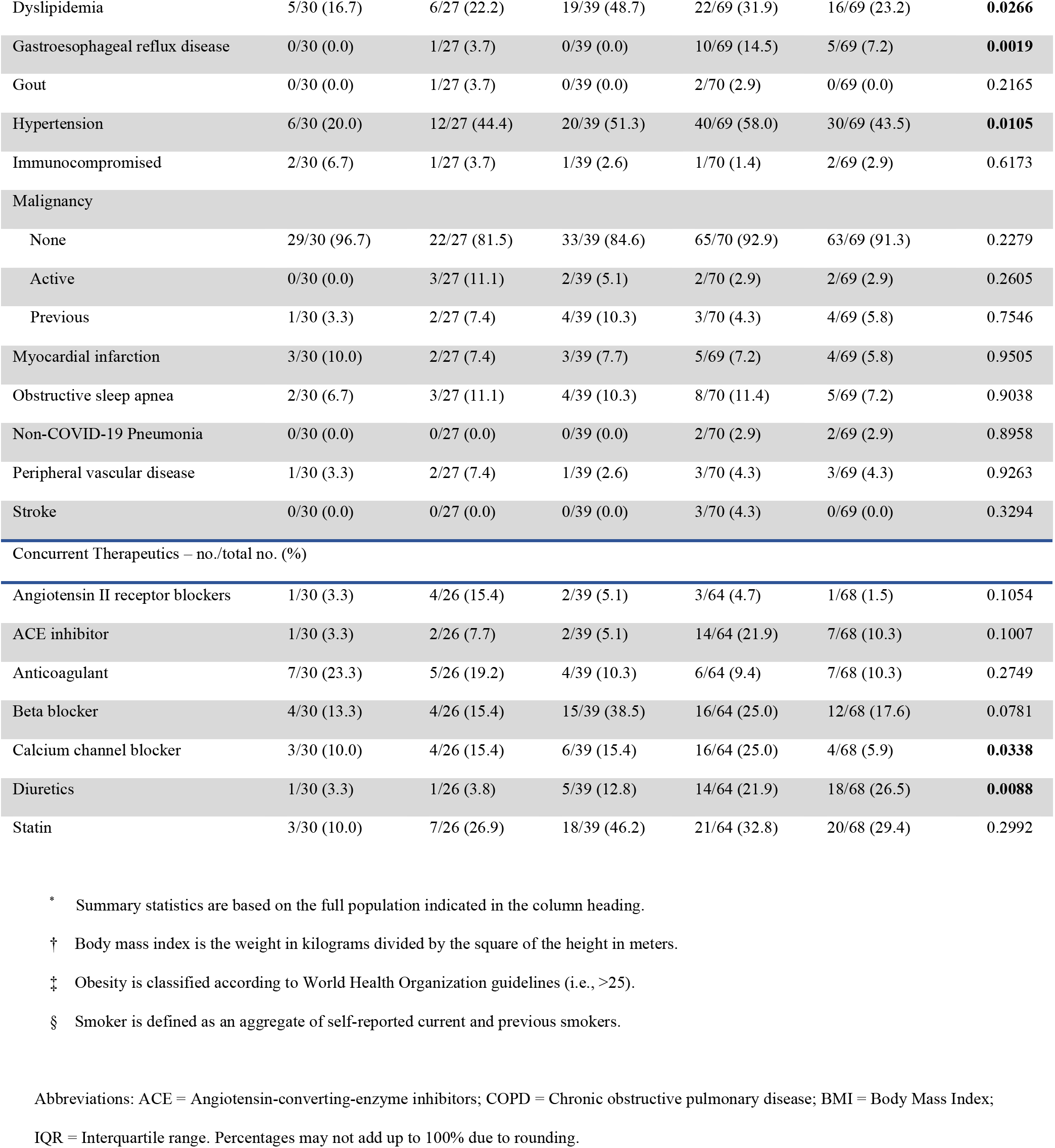
Patient Demographics and Clinical Characteristics of Patients at T_0-1_.

| Characteristics* | Disease Severity |  |  |  |  | P-value |
| --- | --- | --- | --- | --- | --- | --- |
|  | Mild COVID-19 | Mild | Moderate | Severe | Severe COVID-19 |  |
|  | Negative<br>(n=30) | (n=27) | (n=39) | (n=76) | Negative<br>(n=69) |  |
| Demographics |  |  |  |  |  |  |
| Age, median (IQR) – yr. | 47.0 (33.0-60.5) | 59.0 (41.0-70.0) | 74.0 (59.0-86.0) | 61.0 (52.0-71.0) | 61.0 (51.0-72.0) | <0.0001 |
| Distribution – no. (%) |  |  |  |  |  |  |
| 18-40 yr. | 11 (36.7) | 6 (22.2) | 4 (10.3) | 2 (2.6) | 11 (15.9) | <0.0001 |
| 41-64 yr. | 13 (43.3) | 9 (33.3) | 9 (23.1) | 45 (59.2) | 26 (37.7) | 0.0025 |
| ≥65 yr. | 6 (20.0) | 12 (44.4) | 26 (66.7) | 29 (38.2) | 32 (46.4) | 0.0023 |
| Male Sex, no./total no. (%) | 17/30 (56.7) | 15/27 (55.6) | 25/39 (64.1) | 52/76 (68.4) | 46/69 (66.7) | 0.6564 |
| BMI, median (IQR)† | 25.2 (21.3-32.6) | 25.9 (22.5-35.0) | 26.0 (21.1-30.0) | 28.1 (24.2-31.8) | 25.7 (21.8-32.5) | 0.4798 |
| Obesity‡, no./total no. (%) | 4/19 (21.1) | 6/18 (33.3) | 6/27 (22.2) | 38/61 (62.3) | 35/69 (50.7) | 0.0006 |
| Smoking History – no./total no. (%)§ |  |  |  |  |  |  |
| Current smoker | 4/12 (33.3) | 3/9 (33.3) | 3/5 (60.0) | 2/43 (4.7) | 12/69 (17.4) | 0.0027 |
| Previous smoker | 5/12 (41.7) | 0/9 (0.0) | 2/5 (40.0) | 5/43 (11.6) | 4/69 (5.8) | 0.0032 |
| Never smoked | 3/12 (25.0) | 6/9 (66.7) | 0/5 (0.0) | 36/43 (83.7) | 53/69 (76.8) | <0.0001 |
| Length of Admission – days (IQR) | 0 (0-2.3) | 2 (0-13) | 12 (7-24) | 28 (16-65) | 14 (7.5-28) | <0.0001 |
| Mortality | - | 3/27 (11.1) | 1/39 (2.6) | 29/76 (38.2) | 21/69 (30.4) | <0.0001 |
| Co-Existing Disorder – no./total no. (%) |  |  |  |  |  |  |
| Arrythmia | 3/30 (10.0) | 3/27 (11.1) | 9/39 (23.1) | 10/69 (14.5) | 12/69 (17.4) | 0.6140 |
| Asthma | 1/30 (3.3) | 1/27 (3.7) | 2/39 (5.1) | 8/70 (11.4) | 3/69 (4.3) | 0.4970 |
| Cardiac procedure | 2/30 (6.7) | 2/27 (7.4) | 10/39 (25.6) | 6/69 (8.7) | 7/69 (10.1) | 0.9740 |
| Chronic kidney disease | 1/30 (3.3) | 1/27 (3.7) | 13/39 (33.3) | 10/70 (14.3) | 6/69 (8.7) | 0.0204 |
| Congestive heart failure | 3/30 (10.0) | 3/27 (11.1) | 5/39 (12.8) | 5/69 (7.2) | 10/69 (14.5) | 0.7389 |
| COPD | 3/30 (10.0) | 5/27 (18.5) | 5/39 (12.8) | 6/69 (8.7) | 14/69 (20.3) | 0.3134 |
| Coronary artery disease | 6/30 (20.0) | 5/27 (18.5) | 8/39 (20.5) | 6/69 (8.7) | 8/69 (11.6) | 0.2802 |
| Diabetes mellitus | 2/30 (6.7) | 11/27 (40.7) | 16/39 (41.0) | 35/69 (50.7) | 19/69 (27.5) | <0.0001 |
| Dyslipidemia | 5/30 (16.7) | 6/27 (22.2) | 19/39 (48.7) | 22/69 (31.9) | 16/69 (23.2) | **0.0266** |
| Gastroesophageal reflux disease | 0/30 (0.0) | 1/27 (3.7) | 0/39 (0.0) | 10/69 (14.5) | 5/69 (7.2) | **0.0019** |
| Gout | 0/30 (0.0) | 1/27 (3.7) | 0/39 (0.0) | 2/70 (2.9) | 0/69 (0.0) | 0.2165 |
| Hypertension | 6/30 (20.0) | 12/27 (44.4) | 20/39 (51.3) | 40/69 (58.0) | 30/69 (43.5) | **0.0105** |
| Immunocompromised | 2/30 (6.7) | 1/27 (3.7) | 1/39 (2.6) | 1/70 (1.4) | 2/69 (2.9) | 0.6173 |
| **Malignancy** |  |  |  |  |  |  |
| None | 29/30 (96.7) | 22/27 (81.5) | 33/39 (84.6) | 65/70 (92.9) | 63/69 (91.3) | 0.2279 |
| Active | 0/30 (0.0) | 3/27 (11.1) | 2/39 (5.1) | 2/70 (2.9) | 2/69 (2.9) | 0.2605 |
| Previous | 1/30 (3.3) | 2/27 (7.4) | 4/39 (10.3) | 3/70 (4.3) | 4/69 (5.8) | 0.7546 |
| Myocardial infarction | 3/30 (10.0) | 2/27 (7.4) | 3/39 (7.7) | 5/69 (7.2) | 4/69 (5.8) | 0.9505 |
| Obstructive sleep apnea | 2/30 (6.7) | 3/27 (11.1) | 4/39 (10.3) | 8/70 (11.4) | 5/69 (7.2) | 0.9038 |
| Non-COVID-19 Pneumonia | 0/30 (0.0) | 0/27 (0.0) | 0/39 (0.0) | 2/70 (2.9) | 2/69 (2.9) | 0.8958 |
| Peripheral vascular disease | 1/30 (3.3) | 2/27 (7.4) | 1/39 (2.6) | 3/70 (4.3) | 3/69 (4.3) | 0.9263 |
| Stroke | 0/30 (0.0) | 0/27 (0.0) | 0/39 (0.0) | 3/70 (4.3) | 0/69 (0.0) | 0.3294 |
| **Concurrent Therapeutics – no./total no. (%)** |  |  |  |  |  |  |
| Angiotensin II receptor blockers | 1/30 (3.3) | 4/26 (15.4) | 2/39 (5.1) | 3/64 (4.7) | 1/68 (1.5) | 0.1054 |
| ACE inhibitor | 1/30 (3.3) | 2/26 (7.7) | 2/39 (5.1) | 14/64 (21.9) | 7/68 (10.3) | 0.1007 |
| Anticoagulant | 7/30 (23.3) | 5/26 (19.2) | 4/39 (10.3) | 6/64 (9.4) | 7/68 (10.3) | 0.2749 |
| Beta blocker | 4/30 (13.3) | 4/26 (15.4) | 15/39 (38.5) | 16/64 (25.0) | 12/68 (17.6) | 0.0781 |
| Calcium channel blocker | 3/30 (10.0) | 4/26 (15.4) | 6/39 (15.4) | 16/64 (25.0) | 4/68 (5.9) | **0.0338** |
| Diuretics | 1/30 (3.3) | 1/26 (3.8) | 5/39 (12.8) | 14/64 (21.9) | 18/68 (26.5) | **0.0088** |
| Statin | 3/30 (10.0) | 7/26 (26.9) | 18/39 (46.2) | 21/64 (32.8) | 20/68 (29.4) | 0.2992 |

\* Summary statistics are based on the full population indicated in the column heading.
† Body mass index is the weight in kilograms divided by the square of the height in meters.
‡ Obesity is classified according to World Health Organization guidelines (i.e., >25).
§ Smoker is defined as an aggregate of self-reported current and previous smokers.
Abbreviations: ACE = Angiotensin-converting-enzyme inhibitors; COPD = Chronic obstructive pulmonary disease; BMI = Body Mass Index;
IQR = Interquartile range. Percentages may not add up to 100% due to rounding.

The cohort of 241 patients was categorized into three groups that reflected conventional concepts of COVID-19 severity (National Institutes of Health, ‘Clinical Spectrum of SARS-CoV-2 Infection’), as well as two analogous symptom/severity matched control groups.(31) The resulting five groups were: SARS-CoV-2 negative patients presenting to outpatient clinics with symptoms consistent with a respiratory tract illness (n=30, “mild negative”); SARS-CoV-2 positive patients presenting to outpatient clinics with symptoms consistent with a respiratory tract illness (n=27, “mild COVID-19”); admitted SARS-CoV-2 positive patients requiring supplemental oxygenation (n=39, “moderate COVID-19”); SARS-CoV-2 positive patients requiring high-level in-patient ICU care (n=76, “severe COVID-19”); and SARS-CoV-2 negative patients who exhibited symptoms of a severe respiratory disease requiring high-level in-patient ICU care (n=69, “severe negative”). Patients testing negative by nasopharyngeal SARS-CoV-2 polymerase chain reaction had immunological history (i.e., antibody reactivity) ascertained through spike antigen cross-reactivity using a Federal Drug Administration approved enzyme-linked immunosorbent sandwich assay (Online Figure II).

### Data Visualization and Statistical Analysis

All data generated and analyzed which support the findings of this study are included in this article. Associated supplementary information files are available on a publicly accessible archive (see below). *Descriptive Analysis* - Clinical characteristics were characterized using summary statistics. Continuous variables were described using median and inter-quartile range (IQR), and dichotomous or polytomous variables were described using frequencies. Between-group differences were evaluated using Wilcoxon rank-sum tests for continuous variables and Fisher’s exact tests for dichotomous/polytomous variables. Correlation between continuous variables were quantified using Spearman rank correlation. *Descriptive outcome analysis* - The Kaplan-Meier survival method was applied to assess the in-hospital death, and the between-group differences in the freedom from death were evaluated using log-rank tests. The length of hospitalization/ICU was characterized using competing risk models in terms of cumulative incidence rate function. Univariable Cox proportional hazard regression were applied to assess and quantify the association of the baseline clinical characteristics with in-hospital/ICU death. The associations of continuous variables were modeled using natural cubic splines. *Biomarker Analysis* - Comparisons between two independent groups were made using *t-* tests for normally distributed continuous variables or Wilcoxon rank-sum tests non-normally distributed continuous variables. When more than two groups were compared, either a one-way ANOVA with a Tukey or Bonferroni post-hoc test (where appropriate) for multiple testing correction, Kruskal-Wallis one-way analysis of variance with Dunn’s multiple comparison correction. Two-way ANOVA was used to estimate how the mean quantitative variable changes according to time and group differences in leak experiments. Where appropriate, Benjamini-Hochberg false discovery rate (FDR) was utilized with adjusted P values (or Q value where stated) of <0.05 being considered statistically significant and indicated in the graphs as reported by the analysis software with significance thresholds of P<0.05, P<0.01, P<0.001, and P<0.0001 indicated as *, **, ***, **** respectively. MiRNA pathway analysis was conducted using BioCarta/KEGG/Reactome databases and tested for enrichment by a hypergeometric test with adjustment for multiple comparisons using the Benjamini-Hochberg FDR, with P ≤0.05 considered to be statistically enriched in a gene set of interest.(32–34) Although many hypotheses were tested throughout the manuscript, no experiment-wide multiple test correction was applied. Unless indicated otherwise, graphs depict averaged values of independent data points with technical replicates and have error bars displayed as mean +/- standard deviation (±S.D.). Data were analyzed with GraphPad Prism 9.0.0 for MacOS (GraphPad Software, Inc., La Jolla, CA, USA; Biomarker Multiple Comparisons), R(35) (v4.0.3; Spearman Correlation Plots), and FIJI(36) (v2.1.0/1.53c; Quantifying Image Intensities). Final figures were assembled for publication purposes using Adobe Illustrator (v25.4.1).

### Study Approval

This is a multicenter, secondary analysis of a prospectively recruited longitudinal cohort study enrolling consecutive patients with suspected SARS-CoV-2 infection who were referred to two Canadian quaternary care networks in Toronto, Canada from May 2020 to December 2020: University Health Network and St. Michael’s Hospital. All participants who were 18 years of age or older, provided either direct written informed consent or were consented into the study by a lawfully entitled substitute decision-maker on behalf of a participant when lacking the capacity to make the decision. The study, and consenting, was conducted in accordance with protocols approved by the Research Ethics Board (REB) of the University Health Network (REB#: 20-5453.6; Cardiovascular Disease and Outcomes among Patients with SARS-CoV**-**2 Infection During Admission and Post-Discharge [The COVID study]) or St. Michael’s Hospital (REB#: 20-078; COVID-19 Longitudinal Biomarkers in Lung Injury [COLOBILI]). SARS-CoV-2-negative patients with severe respiratory illness symptoms were enrolled within the COLOBILI study.

Detailed materials and methods can be found in the Online Supplement and the Major Resource Tables (Online Tables I and II).

## RESULTS

The baseline characteristics for the cohort are indicated in Table 1. The median age (interquartile range [IQR]) of the entire cohort (n=241) was 61 [51-72] years (155 male patients [64.3%]), and of those 201 were admitted, having a median hospital stay of 18 (IQR, 8-40) days within which 54 (26.9%) died (Table 1). Regardless of admission or SARS-CoV-2 status, co-existing medical conditions were common amongst the population including 83 (out of a total of 234; 35.5%) with diabetes mellitus, 80 (34.4%) with underlying CVD, 31 (12.9%) having chronic kidney disease, and 159 (66.0%) patients having more than one co-existing condition. In this respect, there were significant differences in age (P<0.0001), and the frequency of obesity (P=0.0006), smoking history (P<0.0001), chronic kidney disease (P=0.0204), diabetes mellitus (P<0.0001), dyslipidemia (P=0.0266), gastroesophageal reflux disease (P=0.0019), hypertension (P=0.0105), calcium channel blockers (P=0.0338), and diuretics (P=0.0088) between groups (Table 1). Although a small proportion of patients had documented smoking status, there were 40 (total 138; 29.0%) current or former smokers, with 33 (total 234; 14.1%) patients having chronic obstructive pulmonary disease.

On admission to hospital, clinical lab data highlighted disparities between the groups with leukocytosis and lymphopenia seen amongst more severe groups (e.g., white blood cell count [P<0.0001]; lymphocytes [P<0.0023, Online table III]). Amongst cardiovascular markers, there were significant differences in the frequency of elevated creatine kinase (P<0.0001) and D-dimers (P<0.0001). Although the etiology of admissions was predominantly extracardiac in nature, high-sensitivity troponin I (hs-CnTI) values at t_0-1_ were detectable (>3pg/mL) in 157 patients (65.1%), with values significantly elevated (>15pg/mL in males; >10pg/mL in females) in 90 patients (37.3%, Online Table III), predominantly in the severe groups (P<0.0001).

### COVID-19 Outcomes During Hospital Admission

Among admitted patients with COVID-19, 33 died, with Kaplan-Meier survival curves indicating that patients with severe disease higher mortality risk than did patients with less severe phenotypes (P<0.001, Figure 1A). Furthermore, the only baseline clinical metrics associated with mortality within admitted patients were the history of coronary artery disease (Log-rank, P=0.011) and age at hospital admission (Log-rank, P=0.046, Online Table IV and Online Figure III). During hospitalization, 76 COVID-19 patients (44.2%) were transferred to the ICU (severe COVID-19) immediately upon admission. Of these, 23 (30.3%) were treated with non-invasive ventilation, 53 (69.7%) with invasive ventilation, and 21 (27.6%) underwent extracorporeal membrane oxygenation. When compared to severity-matched SARS-CoV-2-negative controls, patients with severe COVID-19 had on average longer ICU stays (mean 13 [IQR, 7-35] days versus mean 8 [IQR, 3-15] days, P<0.0001), were more likely to display acute respiratory distress syndrome (ARDS, P=0.0195), and had overall worse oxygenation, requiring higher FiO2 (P<0.0001) as well as having lower PaO_2_/FiO_2_ ratios (P=0.0003, Online Table V). Interestingly, although many cardiovascular metrics were unchanged, admitted COVID-19 patients had an increase in the number of noted arrhythmic events (P=0.007) and a higher number of secondary cardiovascular events (P=0.001, Online Table V, Defined in Online Methods). However, when compared to severity matched SARS-CoV-2 negative patients, there were no differences in overall survival (P=0.42; Figure 1B).

**Fig. 1.**
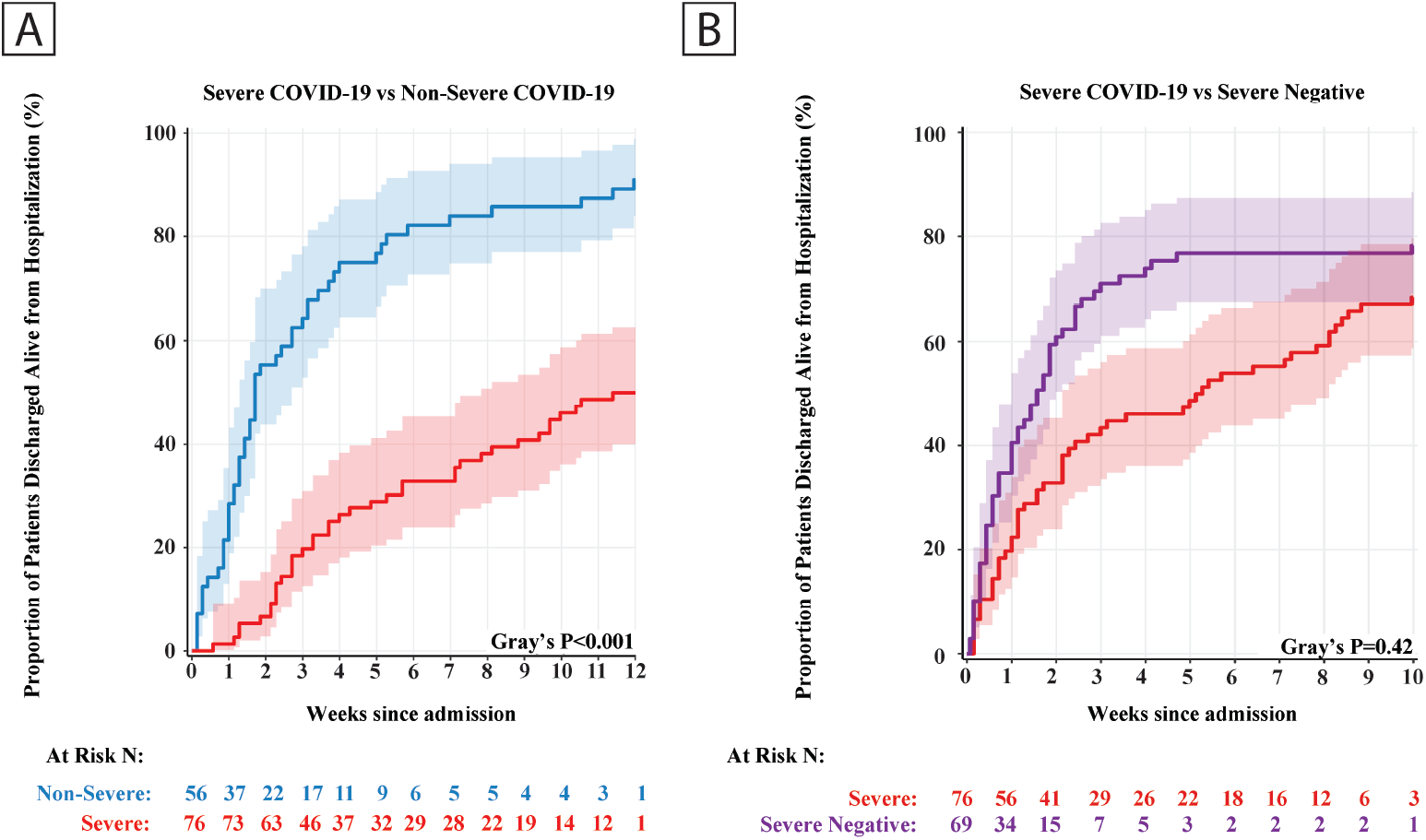
Unadjusted Kaplan-Meier Estimates of Survival. (a) Log-rank test comparing curves of all admitted patients with severe COVID-19 to all admitted non-severe COVID-19 patients (i.e., mild and moderate vs severe, log-rank, P<0.001). (b) Log-rank test comparing curves of those with severe COVID-19 to severity matched SARS-CoV-2 negative patients (P=0.42). Refer to **Online Figure III as well as Online Tables IV-VI for more information.**

### Associations of Cardiovascular and Inflammatory Biomarkers with Outcomes

The vascular endothelium acts as the crucial interface between blood components and tissues, displaying a series of properties that maintain homeostasis (i.e., maintenance of vascular barrier, coagulative capacity, and modulation of immunological responses).(22, 37, 38) While these functions participate in the dynamic regulation of cardiovascular function and coordinate many host defense mechanisms, proinflammatory cytokines are known to elicit a change in endothelial phenotype, promoting thrombosis, local tissue injury, propagating inflammation and potentially contributing to mortality.(39) In order to test the hypothesis that metrics of cardiac damage and inflammatory endothelial dysfunction/activation may better reflect COVID-19 severity and subsequent mortality than standard clinical metrics, a custom Simple Plex assay for markers robustly associated with inflammation and endothelial dysfunction was performed (i.e., Ang-2, endothelin-1 [ET-1], soluble intercellular adhesion molecule [sICAM], soluble vascular cell adhesion molecule [sVCAM], soluble E-Selectin [sE-selectin], sTREM-1, interleukin-6 [IL], and IL-8), along with standalone assays measuring myeloperoxidase (MPO) and high-sensitivity cardiac troponin (hs-cTnI). Correlation analysis revealed that Ang-2, sE-Selectin, and sICAM had moderate correlations with numerous other inflammatory markers, with correlation coefficients ranging from -0.89 (IL-6:MPO) to 0.89 (sICAM-1:Ang-2; Online Figures IV-IX). Myeloperoxidase was the only biomarker without a significant correlation. Among these markers, two were significantly different between mild SARS-CoV-2 positive patients and SARS-CoV-2 negative patients with mild illness at t_0-1_ (elevated Ang-2, P=0.0200; elevated MPO, P=0.0439, Figure 2A, B, Online Figure X). In contrast, nine of the markers (Ang-2, ET-1, sICAM-1, sVCAM-1, sE-Selectin, sTREM-1, IL-6, IL-8, and MPO, Figure 2A, B, Online Figure X) reflected differences among severity within the SARS-CoV-2 positive cohort but failed to demonstrate significance between the critically ill patient groups that did or did not have COVID-19.

**Fig. 2:**
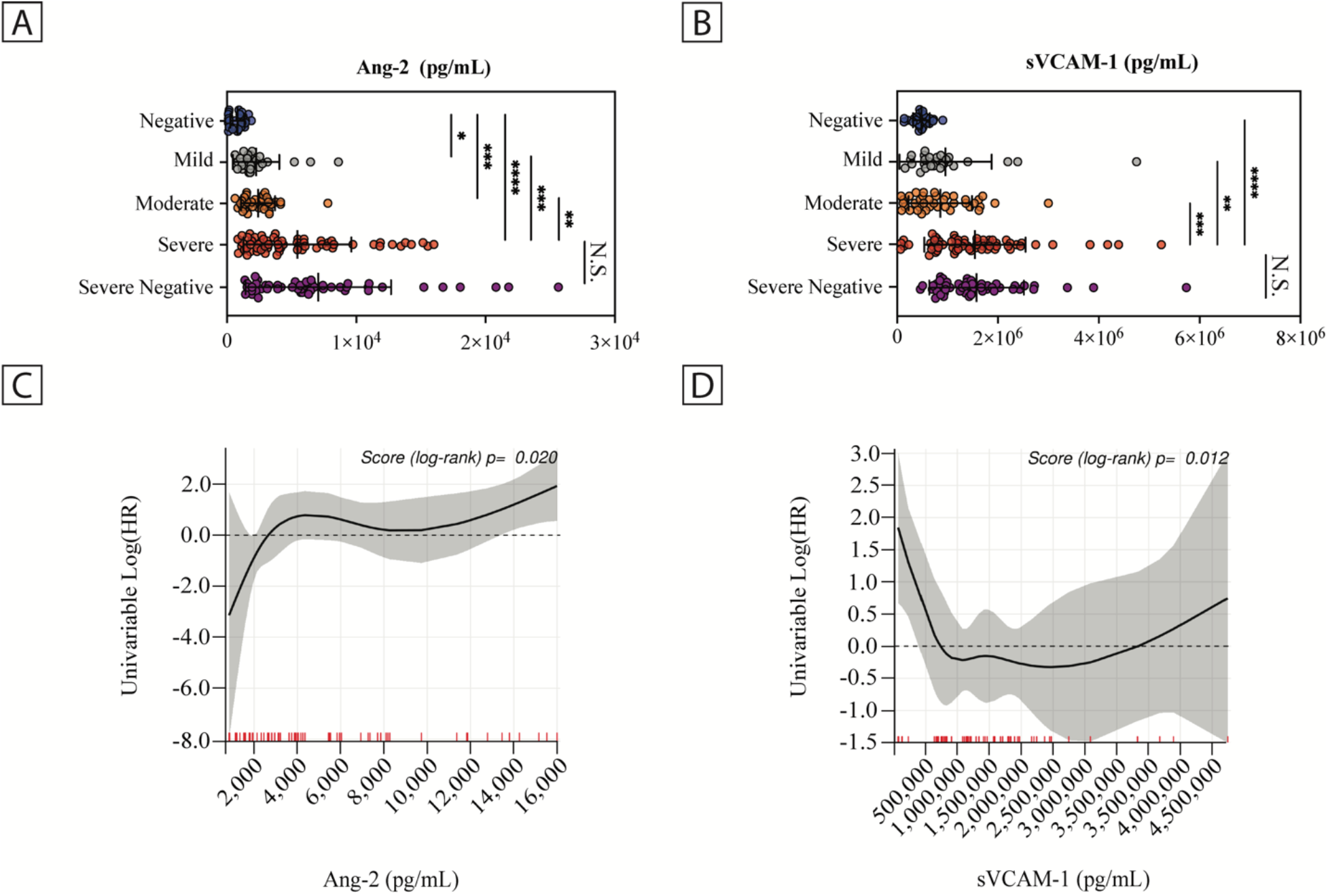
Plasma Concentration of Endothelial Dysfunction and Inflammatory Markers at T_0-1_. (a) Levels of Ang-2 and (b) sVCAM-1 stratified among disease severity. Data shown are for all patients with an available t_0-1_ sample (n=210), with values representing the mean and error bars are (±S.D.). (c) Unadjusted log hazard ratio of Ang-2 and (d) sVCAM-1 in severe disease patients (n=76) with univariable p-value in association with mortality. P values for multiple group comparisons were determined by Kruskal-Wallis test with Dunn’s multiple comparisons test. Severe negative statistical comparisons are only shown in reference to the concordant severe group. Red hash marks indicate individual samples. Abbreviations: Ang-2, Angiopoietin-2; VCAM-1, Vascular cell adhesion molecule 1; N.S., non-significant. Refer to **Online Figure IV-XIII for more information and additional endothelial and inflammatory markers.**

Amongst all patients admitted with COVID-19, univariable analysis revealed that only Ang-2 was associated with mortality (P=0.015; Online Table IV), suggesting higher concentrations are associated with higher mortality, while both Ang-2 (P=0.020) and sVCAM-1 (P=0.012) were associated with mortality when looking specifically at the severe COVID-19 patients (Figure 2C, D, Online Table VI). Sub-stratifying these markers by severity, there were significantly higher t_0-1_ concentrations of Ang-2, IL-6, and MPO in non-survivors with COVID-19 when compared to the severe SARS-CoV-2 negative patients (Online Figure XI). Over time, only IL-6 and MPO remained significantly different between severe COVID-19 and severe negative patients (Online Figure XII and XIII). Taken together, while markers of inflammation/endothelial dysfunction were observed at early timepoints and associated with severity, the majority were not specific to either COVID-19 status or mortality.

### Plasma MiRNA Atlas of SARS-CoV-2 Infection Reveals Markers Specific to COVID-19 and Mortality

While select inflammatory/EC activation markers were informative for ICU mortality, they lacked the ability to distinguish between COVID-19 and non-COVID-19 pathology. We postulated that assessment of the circulating miRNA transcriptome may provide further precision with respect to patient subgroups, since miRNA profiling (in contrast to circulating protein markers) has been shown to effectively differentiate complex disease etiologies.(40, 41) Using whole transcriptome miRNA sequencing (2,083 mature miRNAs) we screened plasma obtained from the differing groups of disease severity to identify meaningful differences in miRNA composition (n=30, negative mild; n=14, mild COVID-19; n=15, moderate COVID-19; n=36, severe COVID-19; n=33, negative severe). Comparative analyses indicated that there were substantially higher numbers of differentially expressed miRNA as disease severity progressed (Figure 3A; Fold ≥±1.5 and Q<0.05). Comparing severe COVID-19 to severe negative patients revealed 765 differentially expressed miRNAs that could subsequently be used for group differentiation (Figure 3B). DIANA-mirPath pathway analysis on the differentially expressed miRNAs suggested broad enrichment of pathways including those related to cardiomyocyte function (i.e., ErbB2 signaling and arrhythmogenic right ventricular cardiomyopathy) as well as adherens junctions (Figure 3C). Sub-analysis of the severe COVID-19 patients for mortality revealed 207 differentially expressed miRNA between survivors and non-survivors (Figure 3D), including pathway enrichment for platelet activation, extracellular matrix-receptor interactions, Ras, and ErbB2 (Figure 3E). A full list of differentially expressed miRNA and predicted KEGG pathways is available in Online Data Files IV and V as well as Online Figures XIV and XV.

**Fig. 3:**
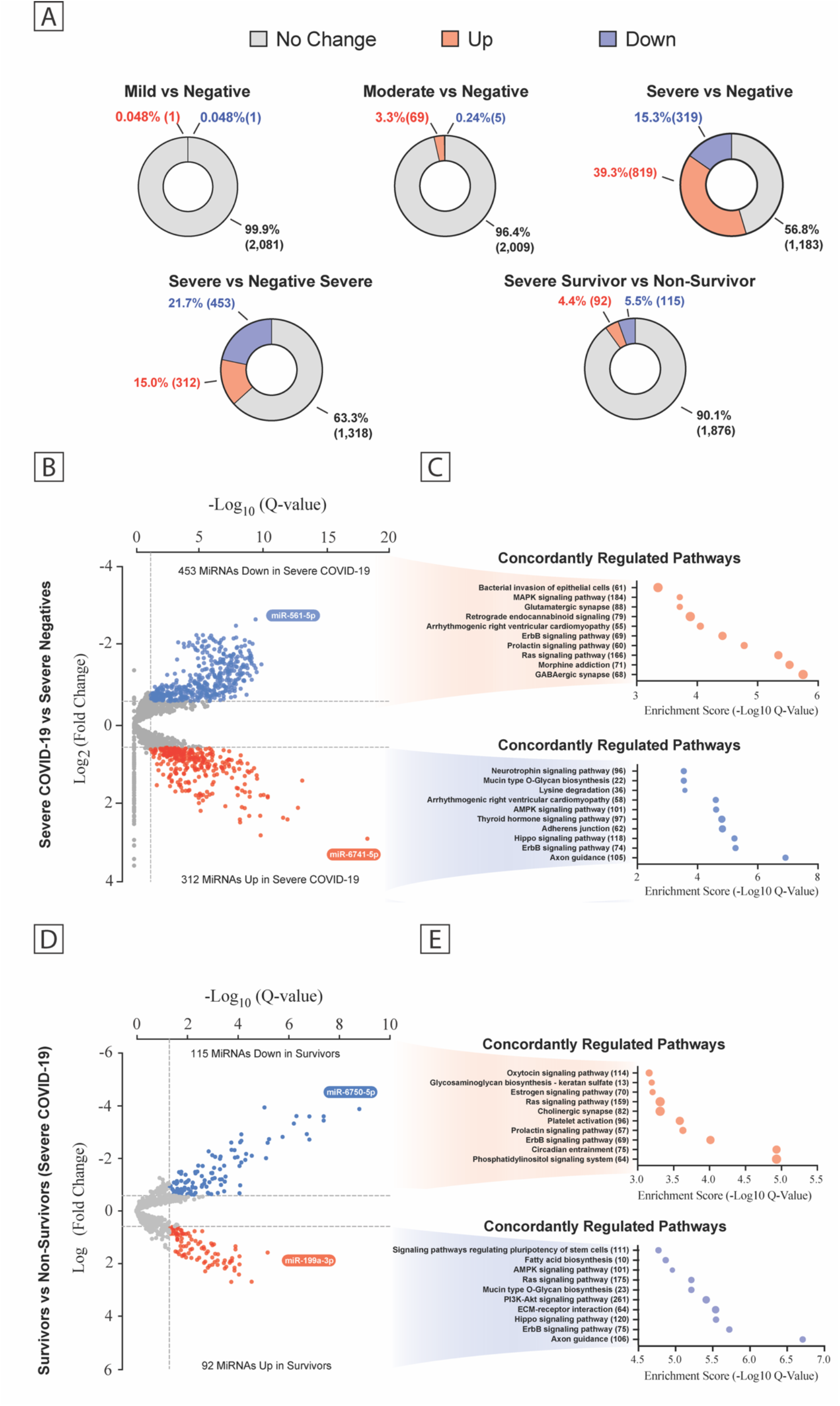
Plasma MiRNA Transcriptome Across the COVID-19 Severity. (a) Pie chart percent modulation of the transcriptome for all subgroups studied. Volcano plots of differentially expressed miRNA between patient groups (b, d) with predicted KEGG terms (with enrichment score below and number of genes to the right) for pathways of deregulated miRNAs shown beside each corresponding region of the volcano plot (c, e). Data are displayed as false discovery rate (FDR) adjusted P values (Q values) vs the log_2_ fold change, with dashed lines drawn to define restriction boundaries. Refer to **Online Figure XIV-XV for more information.**

### Predictive Power of Clinical, Protein, and miRNA Data on Mortality using Machine Learning

Given the high dimensionality of the dataset generated, we sought to examine the utility of models developed using machine learning to predict in-hospital mortality of COVID-19 patients at admission, based on common clinical data, protein expression data, and miRNA expression data. We performed 250 experiments using repeated randomized stratified sub-sampling cross-validation into disjoint sets of 80% training and 20% testing, to train a set of Random Forest models.(42) We assessed model performance by the area under the receiver operating characteristic (AUROC) calculated on the testing sets. Aggregate statistics on AUROC were calculated across the 250 experiments. We observed a low AUROC using only clinical features available at the time of admission (AUROC 0.44, 95% CI 0.22-0.69, Figure 4A).(42) However, incorporation of either the protein expression data or miRNA data improved the performance over the models that used conventional clinical predictors alone, having AUROCs of 0.82 (95% CI 0.64-0.98) and 0.76 (95% CI 0.56-0.96) respectively (Figure 4B, C). In the clinical data only models, ranking input variables that contributed to model predictivity revealed age and body mass index were amongst the most important (Online Figure XVI). Furthermore, in contrast to the traditional univariate statistical analyses, MPO, TREM-1, and Ang-2 were listed as the top contributing features in the synthesized clinical and protein multivariate machine learning model, coming before any other traditional clinical factors (Online Figure XVII). Owing to the exceedingly high dimensionality (2,083 features) machine learning did not highly rank specific variables within the synthesized clinical and miRNA model with statistical reliability.

**Fig. 4:**
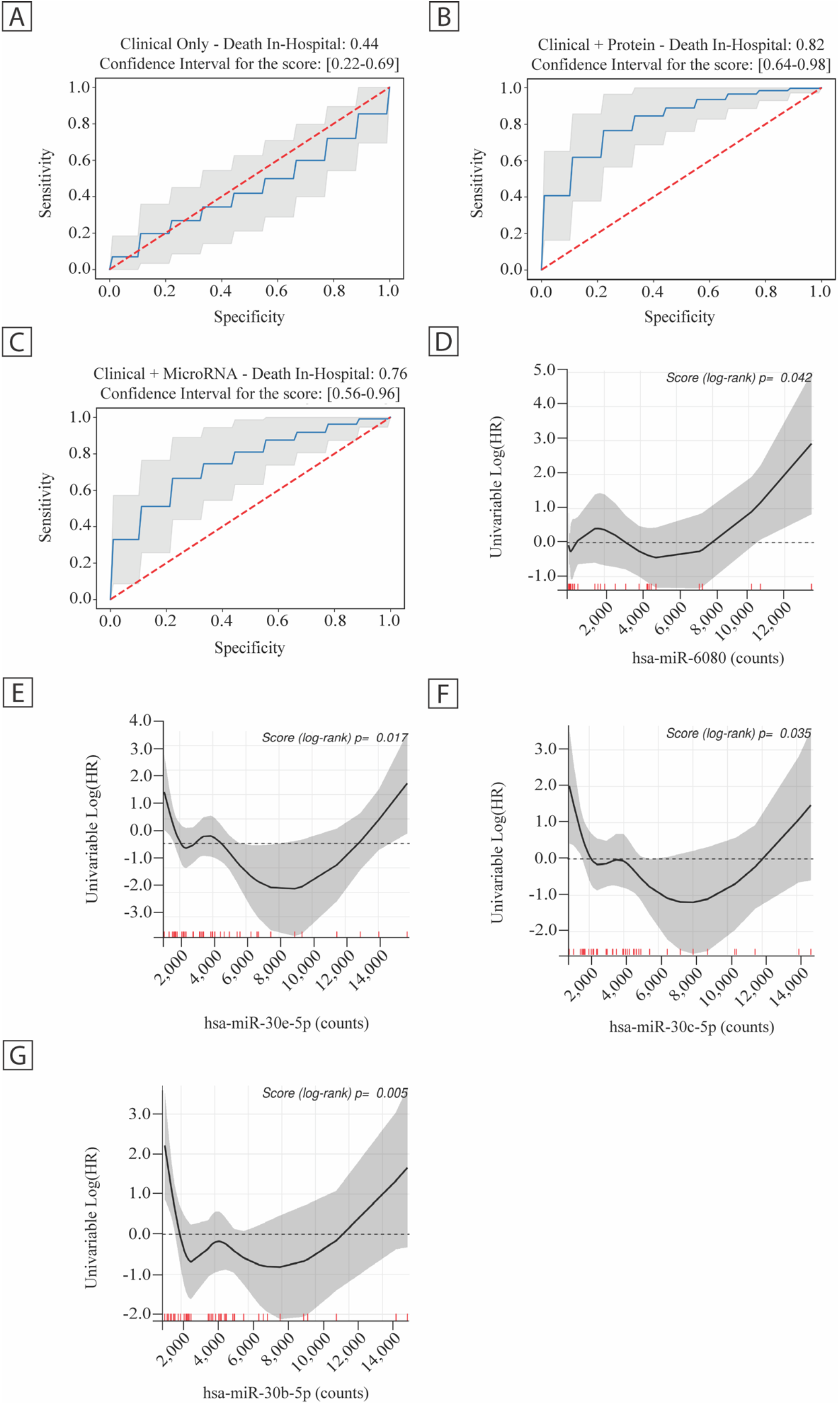
Machine Learning Approach to Risk Assessment and Association of Biomarkers with In-Hospital Mortality for Severe COVID-19 Patients. Assessment of datasets using repeated randomized stratified sub-sampling cross-validation (Random Forest machine learning) for (a) clinical data, (b) clinical data and protein expression metrics, and (c) clinical data and miRNA atlas expression metrics. Univariable log hazard ratios of candidate microRNAs (d) hsa-miR-6080, (e) hsa-miR-30e-5p, (f) hsa-miR-30c-5p, and (g) hsa-miR-30b-5p in relation to mortality. Red hash marks indicate individual samples. Refer to **Online Figure XVI-XVII for more information.**

### Identification of miRNAs Associated with Severe COVID-19 Mortality

Since miRNAs provided superior specificity for COVID-19 mortality compared to protein biomarkers (Figure 2, 3), we next sought to identify candidate miRNA markers that significantly contribute to mortality risk. We considered miRNAs within the top 50% of abundance and cross-examined the differential expression between the both survivors and non-survivors of COVID-19 as well as SARS-CoV-2 severe negatives. More so, we took into consideration existing biological relevance, thereby specifically analyzing miR-1 which is associated with myocardial injury(43, 44), miR-199a-3p which has been shown to be cardioprotective(45), miR-181a-5p which has been shown to restrict vascular inflammation(46), along with members of the miR-30 family which are enriched in ECs and capable of modulating inflammation(47–49), as well miR-339-3p and miR-6080 which were among the highest differentially expressed. Univariable hazard ratios and log-rank P-values were generated to determine the relationship of the miRNA expression measured in plasma with mortality. When ranking by significance of independent association with mortality, miRNAs are among the highest ranking factors comparing to other clinical metrics (Online Table IV and VI) highlighting that miRs-30b/c/e, -6080, -181a-5p, -199a-3p, and -339 (Figure 4D-E, Online Figures

XVIII) are specific for COVID-19 severity and mortality. In contrast, miR-1, failed to show a significant association with mortality (Online Table VI).

### Endothelial Barrier Disruption is Rapidly Induced During Co-incubation with Plasma from Patients With Moderate to Severe COVID-19

With studies showing conflicting evidence that SARS-CoV-2 can directly infect the endothelium in a physiologically meaningful fashion(12, 50, 51), we reasoned that modulation of the extracellular milieu (i.e., changes to circulating plasma components such as sTREM-1, Ang-2, and MPO) as a result of the systemic immune response may be a driving factor behind the observed endothelial dysfunction. In this context, data from previous studies conducted in systemic inflammatory response syndrome and sepsis have underscored the impact of endothelial barrier disruption on disease outcomes.(52) The data presented so far suggested that dysfunction of the endothelium may also accompany COVID-19 and may be associated with poor outcomes. To further examine this hypothesis, we utilized two validated permeability platforms using an endothelial monolayer model: continuous monitoring of transendothelial electrical resistance (Figure 5A) using the xCelligence platform, and a transwell system consisting of a confluent monolayer of pooled human umbilical vein ECs on a permeable membrane. In a timeframe that would exclude viral replication(20, 53), cells were treated with 20% (v/v) plasma from the t_0-1_ samples from across the disease severity spectrum of COVID positive and negative patients. Moderate and severe COVID-19 patient plasma induced significant endothelial barrier dysfunction, while the mild and mild negative patient plasma did not induce significant EC leak in the xCelligence assay (Figure 5B). Similarly, using a validation cohort, the integrity of the monolayer was gauged through leakage of a large dextran tracer across the EC barrier in both acute (i.e., one-hour) and longer-term (i.e., six-hour) treatment. This revealed barrier disruption in response to moderate and severe COVID-19 patient plasma, but not in response to plasma from severe negative patients (Online Figure XIX). Of the mediators tested, EC permeability correlated with levels of Ang-2, hs-cTnI, ET-1, IL-6, IL-8, sTREM-1, and (Online Figure XX). These experiments provide *in vitro* evidence that barrier dysfunction can be independent of direct SARS-CoV-2 infection. We reasoned that elucidating the unique pathways through which the more severe COVID-19 phenotypes exert their barrier disruptive effects may reveal potential therapeutic approaches to maintain EC barrier.

**Fig. 5:**
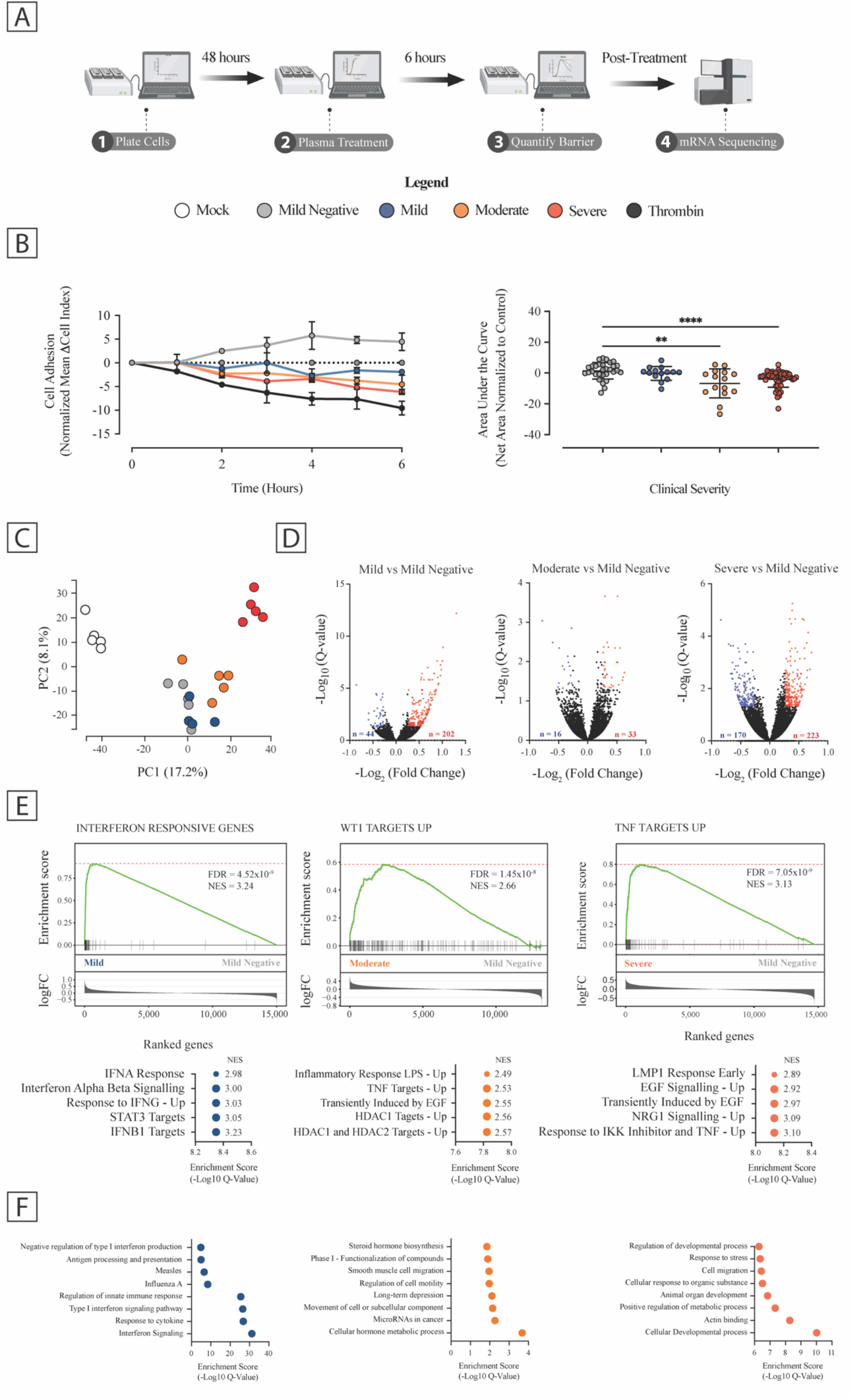
Endothelial Barrier Disruption Driven by Modulation of Inflammatory and Cytoskeletal Pathways is Disease Severity Dependent. (a) Overview of parallel endothelial phenotype monitoring. The overall scheme is shown, representing the real-time monitoring of EC barrier function (xCelligence) followed by both mRNA sequencing and multiplexed immunohistochemistry. (b) Time-course monitoring of pHUVEC barrier function during coincubation with 20% (v/v) plasma (left) sampled from t_0-1_ (negative, n=30; mild, n=14; moderate disease, n=15; severe disease, n=52; thrombin, n=7) compared to ‘mock’ treatment (i.e., PBS control) and quantification of area under the curve across the six-hour coincubation period (right). Thrombin treatment was included as a barrier disrupting positive control. Barrier data displayed depicts t_0-1_ adjusted values (media only) that were subsequently normalized to mock-treated cells (PBS, dashed line) and displayed as the mean of the change in cell index from experiment initiation. Note: some error bars are too small to be visible on the figure. Quantified values are relative to ‘mock’ treatment and represent mean and error bars (±S.D.). P values determined by a Kruskal-Wallis one-way analysis of variance with Dunn’s multiple comparison correction. moderate vs negative, p=3.1×10^-3^; severe vs negative, p=4.7×10^-5^. (c) Principal component analysis plot, with the 2D coordinates of each profile based on the scores of the first two principal components. (d) Volcano plot displaying the −log_10_ of the adjusted P values vs the log_2_ fold change of respective disease severities compared with COVID-19-negative control transcript expression. Red and blue markers indicate adjusted FDR-adjusted P values <0.05 for up- and down-regulation, respectively, based on a log fold-change of >±0.58. (e) Gene set enrichment plot of the top-ranked gene set, TNF Targets Up (FDR = 7.05×10^-9^, NES = 3.13), WT1 Targets Up (FDR = 1.45×10^-8^, NES = 2.66), and interferon responsive genes (FDR = 4.52×10^-9^, NES = 3.24) using all genes ranked by their magnitude of association with each respective disease severity group (the enrichment P value shown was computed from the GSEA test) along with top ranked gene sets (below). The tick marks denote the location of the genes in each respective module. Fold change of all genes between the compared conditions are shown as bar plots in the bottom panels (x axis: genes ranked by -log_2_ fold change; y axis: -log_10_ fold change). (f) Top ten significantly enriched pathways based on all genes ranked by fold change identified by gProfiler are shown for each comparison. See Online Data Files IV, V, and VI for a full list of differentially expressed genes, GSEAs, and pathways. Refer to **Online Figures XIX-XX for more information.**

### Modulation of Inflammatory and Cytoskeletal Processes are Coincident with Barrier Dysfunction

To gain a better understanding of how the endothelium is modulated by plasma components, we investigated the gene-level changes using RNA-sequencing of ECs after six hours of co-incubation with patient-derived plasma. On the transcriptome level (16,285 total quantified genes, having ≥10 reads in at least five samples), biological replicates within groups were tightly correlated (Pearson r=0.975-0.988, n=4-5), suggesting robust intra-group clustering even among a heterogeneous patient group, similar to previous studies.(27) Principal component analysis of the normalized transcriptome showed segregation between experimental groups, with the severe group being clearly distinct from the mild and moderate groups (Figure 5C). Pair-wise differential expression analysis with the mild SARS-CoV-2 negative cohort as the control revealed 393, 49, and 246 genes are differentially expressed (FDR < 0.05, log_2_ fold change >±0.58) in the mild, moderate, and severe COVID-19 cohorts, respectively (Figure 5D). Gene set enrichment analysis revealed that co-incubation with either severe or moderate COVID-19 patient plasma altered the expression of endothelial genes related to acute inflammatory response, angiogenic programs, or histone deacetylase activity, whereas administration of plasma from mild COVID-19 patients altered the expression of genes involved in priming of antiviral responses (Figure 5E and Online Data Files VI, VII, and VIII). Similarly, pathway enrichment analysis using gProfiler with significant differentially expressed genes highlighted a predominance of KEGG pathways relating to interferon in the mild group, while in contrast, pathways in ECs exposed to moderate and severe COVID-19 plasma related more broadly to cell motility, developmental processes, cell stress responses, cell structure reorganization, and actin mobilization (Figure 5F). Collectively, these results suggested endothelial structural changes could be occurring that might be amenable to treatment with barrier stabilizing agents.

### Vascular Barrier Stabilizing Drugs Prevent EC Permeability Induced by COVID-19 Patient Plasma Exposure *in vitro*

We next examined two principal structural contributors to the EC barrier, vascular endothelial-cadherin (VE-Cadherin) and Claudin-5. VE-cadherin is an essential adherens junction protein that regulates cell-cell junctional stability, and Claudin-5 is a tight junction protein that regulates size-dependent paracellular permeability pathways.(54) Immunostaining confirmed significant disruptions to both VE-cadherin and Claudin-5 expression as well as junctional localization, particularly in ECs exposed to the moderate and severe COVID-19 plasma (Figure 6A). To gauge the potential clinical importance of these changes in relation to vascular leak, we next investigated whether targeted therapeutics that are known to stabilize the vascular barrier or suppress EC activation can prevent COVID-19 plasma-induced permeability *in vitro*. The following drugs that have been reported to reduce endothelial dysfunction were utilized: Q-peptide, a synthetic integrin-binding motif of angiopoietin-1(55); Slit2-N, a recombinant member of the Slit family of secreted extracellular matrix glycoproteins that stabilizes adherens junctions(56); Nangibotide, a TREM-1 inhibitor(57); and dexamethasone, a potent synthetic adrenal corticosteroid currently used in COVID-19 treatment(58). Co-treatment with either Q-peptide or Slit2-N prevented the disruption of the EC barrier, as examined through junctional protein expression (Figure 6B, C, left) and permeability as examined by xCelligence readout (Figure 6B, C, right). While other agents could maintain junctional protein expression (e.g., Nangibotide, dexamethasone) in specific settings, no other agents were universally able to maintain barrier protein expression in the face of moderate or severe disease plasma exposure and prevent the physiologic barrier disruption measured by electrical resistance.

**Fig. 6.**
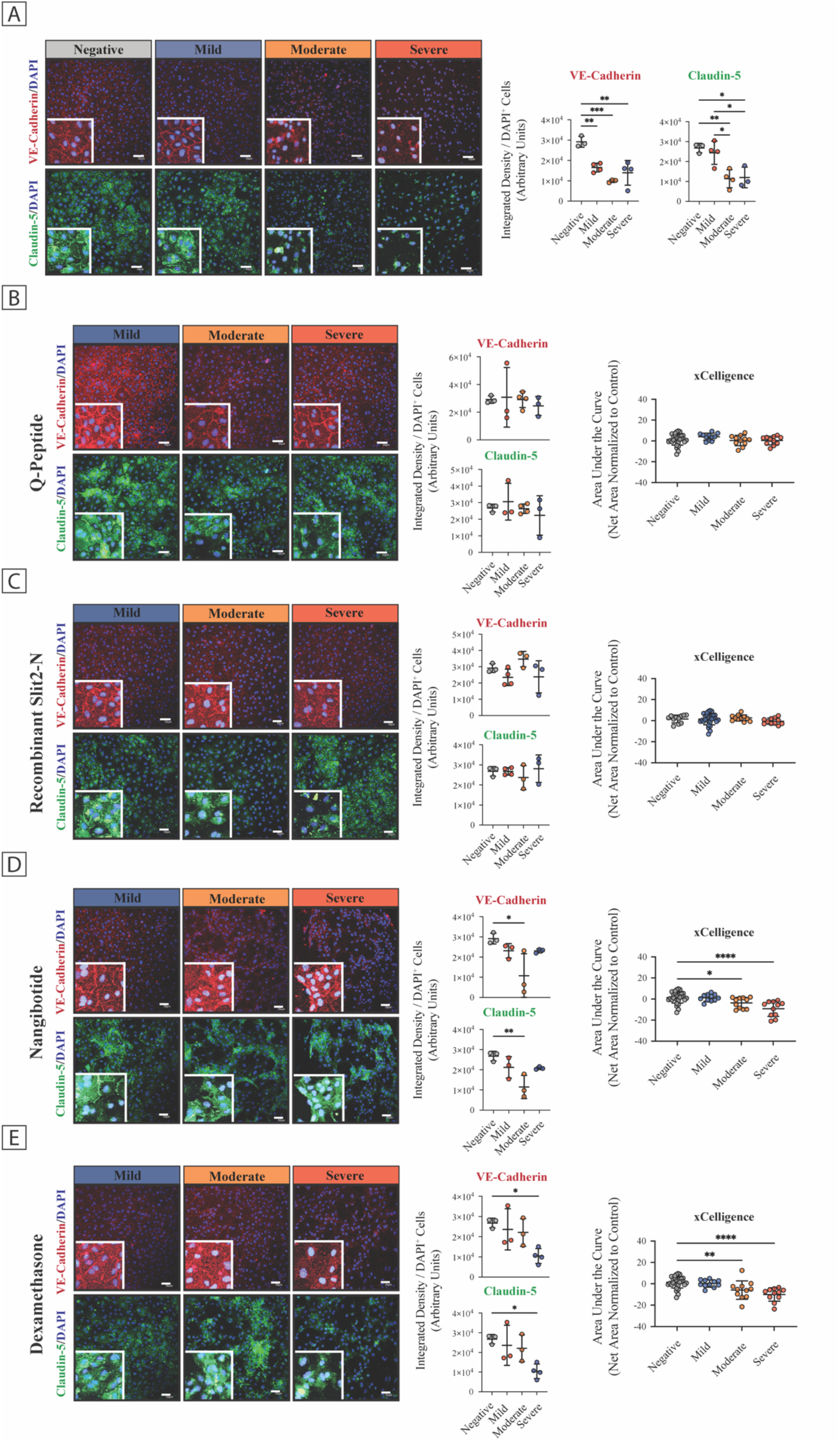
Targeted Modulators of Endothelial Barrier Protect Against COVID-19 Plasma-Induced Endothelial Barrier Dysfunction. (a) Left; Representative confocal microscopy images and brightness enhanced zooms depicting VE-cadherin (red) and Claudin-5 (green) junctional staining after six hours of treatment of pHUVECs with t_0-1_ COVID-19 plasma and matched controls; n=3-4 per group. Scale bars: 50 µm. Right; Quantification of VE-cadherin and Claudin-5 pixel intensity for each respective group. Each dot represents mean protein expression across at least four representative sections of a biological replicate. Center bars represent mean and error bars are (±S.D.). P values were determined by one-way ANOVA with Tukey’s multiple comparisons test. Mild vs negative: VE-cadherin, P=6.0×10^-3^. Moderate vs. negative: VE-cadherin, P=4.0×10^-4^; Claudin-5, p=1.6×10^-2^. Negative vs severe: VE-cadherin, P=1.6×10^-3^; Claudin-5, P=8.0×10^-3^. Mild vs moderate: Claudin-5, P=2.9×10^-2^. Mild vs severe: Claudin-5, P=1.4×10^-2^. (b-e) Left; Representative confocal microscopy images depicting a combinatorial screen of COVID-19 plasma and matched controls treated with either (b) Q-peptide, (c) Slit2-N, (d) Nangibotide, and (e) Dexamethasone on pHUVECs reveals modulated maintenance of VE-cadherin (red) and Claudin-5 (green) expression; n=3-4 per group. Scale bars: 50 µm. Middle; Quantification of VE-cadherin and claudin-5 for each respective group. Imaging P values were determined by one-way ANOVA with Dunnett’s multiple comparisons test (all compared to control). Nangibotide; Moderate vs negative: VE-cadherin *P=1.3×10^-2^, Claudin-5 **P=8.2×10^-3^. Dexamethasone; Severe vs negative: VE-cadherin *P=2.4×10^-2^, Claudin-5 *P=3.3×10^-3^. Right; Quantification of VE-cadherin and Claudin-5 for each respective group. Each dot represents mean protein expression across at least four representative sections of a biological replicate. Center bars represent mean and error bars are (±S.D.). Right; TEER quantification of corresponding treatment groups. P values were determined by Kruskal-Wallis test with Dunn’s multiple comparisons test (all compared to control). Nangibotide; Negative vs moderate (P): *P=3.9×10^-2^; Severe vs negative ****P=4.3×10^-5^. Dexamethasone; Moderate vs negative (P): **P=6.2×10^-3^; Severe vs negative****P=2.2×10^-5^. Refer to **Online Figure XXI for more information.**

## DISCUSSION

Recent literature on SARS-CoV-2 pathogenesis has suggested that the induction of substantial acute respiratory distress phenotypes is driven by a mismatched inflammatory response together with broad vascular dysfunction.(59, 60) While several detailed reports implementing multi-omic approaches have provided insight into the immune cell phenotypes involved in these processes(61–63), risk stratifying immune markers specific to COVID-19 have not been fully elucidated. Herein, we provide a comprehensive, multi-omics-based description of the molecular antecedents to COVID-19 mortality, yielding new insights pertaining to the vasculature. Our study further delineates the gradient of vascular dysfunction observed in patients across the spectrum of COVID-19 severity, particularly among those with severe illness. While our findings are consistent with smaller cohort studies examining single markers of cardiovascular dysfunction(26, 64), our report is the first to use proper, disease-negative controls, which allowed us to ask etiological questions. Our analysis revealed that although markers of cardiovascular dysfunction (such as Ang-2 and sVCAM-1, Figure 2A, B) tracked well with COVID-19 severity and outcomes, these markers were similarly elevated in ICU patients without COVID-19, which indicates they are more appropriate as general markers of disease severity, and they are not COVID-19 specific.

In recent years, circulating miRNAs have emerged as exquisite biomarkers for complex pathological conditions, including influenza and sepsis.(65, 66) The inherent stability of plasma microRNAs under harsh conditions and reproducible quantification makes them attractive candidates for use as noninvasive biomarkers.(30) Incorporating whole transcriptome sequencing as an added metric allowed for the effective differentiation between severities of COVID-19 and severe SARS-CoV-2 negative controls using samples gathered on admission. Through this study, we provide further empirical evidence for the value of data from novel biomarkers over metrics traditionally utilized in healthcare systems (i.e., clinical demographics and laboratory data). Notably, we show that the data from electronic medical records fails to adequately capture the risk of in-hospital mortality. Using a Random Decision Forest machine learning model on our multi-omic biologically relevant datasets we were able to develop an exploratory prognostic risk prediction model that incorporates markers that are COVID-19 and vascular specific. Importantly, there are several platforms that allow the interrogation of candidate miRNAs within clinically relevant timeframes.(67, 68) While still investigational, given the extent of pre-clinical and clinical research on miRNAs(40), reasonable to expect miRNA datasets will soon be amenable for clinical implementation. As an example of the importance of a multi-omic approach that includes clinically relevant disease negative controls, miR-1 (which has been intensively studied as a cardiac enriched miRNA and associated with numerous cardiac etiologies(41, 43, 44, 69)) failed to show an association with in-hospital mortality in our study. If the promising miRs-30b/c/e, -6080, -181a-5p, -199a-3p, and -339 identified here are validated in larger populations they would represent new biomarkers that could be utilized for rapid in-hospital risk assessment. As miRNAs represent functional biomarkers, having active roles in gene regulation, they also present an important opportunity to understand the pathophysiological relevance of endothelial-based processes affected by SARS-CoV-2 infection.

Given that the endothelium is the gatekeeper of vascular permeability, which is a main pathophysiological process in systemic inflammatory conditions, there is intense research on their role in ARDS, systemic inflammatory response syndrome, and sepsis (among others).(70–72) Several lines of evidence support the hypothesis that endothelial function is a major determinant of COVID-19 outcome.(64, 73) Studies have shown endothelial derangements (i.e., pathological sprouting angiogenesis), increased endothelial apoptosis, modified metabolism, and a strong correlation between underlying CVD and mortality as COVID-19 severity increases.(23, 25, 70) Often forgotten, endothelial dysfunction–associated molecular patterns are broad and strong activators of innate immune responses, leading to innate immunity-mediated organ injury. To this end, we observed that plasma from patients with moderate and severe COVID-19 induced profound barrier disruption, as assessed through the modulation of VE-Cadherin and Claudin-5 at EC-EC junctions. Transcriptomic analysis revealed that COVID-19 plasma from moderate to severe patients appears to preferentially induce pro-inflammatory immune gene processes, while plasma from mild patients induces an interferon (IFN) response. This is in line with several studies on the immune response of mild vs severe COVID-19 patients, and confirmed by the over-representation of type I IFN genes in ECs exposed to mild COVID-19 plasma.(64, 74, 75) Type I IFNs are critical mediators of the antiviral immune response and subverting the early type I IFN response has been shown repeatedly to be a contributor to coronavirus infection.(74) Looking at the moderate and severe phenotypes where endothelial pro-inflammatory responses (e.g., TNF) and histone deacetylase dysregulation occurred, both epigenetic modifications as well as TNF responses have been documented in COVID-19.(64, 75)

Increased pro-inflammatory cytokine expression, specifically TNF and IL-6, has been shown to upregulate trypsin, resulting in the loss of endothelial tight junctions, subsequently inducing vascular hyperpermeability.(76, 77) We are encouraged by our findings that both Q-peptide and recombinant Slit2-N, can effectively prevent the induction of endothelial permeability *in vitro*. With the known roles of Ang-1 and Ang-2 as ligands of the endothelial cell-specific integrins, and their antagonistic relationship as modulators of endothelial survival, the barrier maintenance seen through Q-peptide could be due to antagonism of higher Ang-2 levels in more severe COVID-19.(78, 79) Additionally, measurement of the levels of endogenous sSlit2 in SARS-CoV-2 patient plasma revealed a significant upregulation only in patients with severe disease suggesting a possible compensatory mechanism and furthering the biological relevance of Slit2-N treatment (Online Figure XXI). Therefore, as a proof-of-concept, these data support the hypothesis that disruption of endothelial barrier function, particularly as a result of loss of adherens and tight junctions, is induced during COVID-19. These findings support a model in which systemic induction of pro-inflammatory processes, rather than direct infection, may be the primary driver of systemic endothelial perturbations observed in COVID-19 patients, and that relative to current treatments aimed at dampening immune responses, preserving endothelial barrier may be a viable adjuvant to reduce the mortality of SARS-CoV-2.

Furthermore, Q-peptide and Slit2N act via different pharmacological pathways, suggesting that preservation of endothelial barrier itself is the key therapeutic mechanism. It will nonetheless be important to determine whether pre-existing and potentially chronic endothelial permeability – layered on top of a permeabilizing stimuli such as infection – can be reversed through biologics that interact directly at the endothelial interface. If the significant induction of endothelial permeability can be overridden, even in patients with an ongoing inflammatory insult, then pharmacological intervention at the level of the vasculature may be an effective adjuvant therapy for patients at a higher risk of adverse events. Indeed, previous studies using animal models of sepsis, influenza, and malaria have reaffirmed that pharmacological treatments that enhance vascular resiliency in the face of an aberrant host immune response may be a therapeutically feasible approach for managing vascular dysfunction without compromising immunity.(80–82) Although stabilizing the vasculature may be more practical than attenuating individual cytokines, it remains to be determined whether severe vascular damage can be reversed in a clinically meaningful way. Anecdotally, recent reports of adrenomedullin, a peptide hormone capable of endothelial stabilization, have suggested high *in vivo* efficacy and positive results from both Phase I and II trials in patients with sepsis.(83, 84) A prospective study comparing endothelial function in critical illness of non-viral etiology and viral etiology may provide clearer insights as to the translatability of these findings.

Taken together, we have provided the first highly salient comparison of a diverse group of cardiovascular markers between COVID-19 positive and negative patients, highlighting that well-known markers of inflammation, cardiac damage, and endothelial dysfunction are not specific to COVID-19 pathology. Notably, with a targeted miRNA transcriptomic approach, we were able to discern specific markers that showed better discrimination between these two groups. Using machine learning, incorporation of protein and miRNA markers improved the prediction of in-hospital mortality over baseline clinical variables. While exploratory, this is a clinically feasible approach that has the advantage of using pathophysiologically relevant SARS-CoV-2’s markers, as opposed to the surrogate markers used in most other published risk stratification models.(10, 26) Finally, our data provide several lines of evidence supporting the notion that endothelial barrier function is affected in a SARS-CoV-2 specific manner, that is distinct from the pathways involved in critically ill patients with non-COVID-19 severe respiratory illnesses. Our data reinforce the idea that barrier dysfunction is likely independent of direct viral infection and instead secondary to yet undiscovered mediators in the plasma. We further pursued our observations using assays of EC function using an *in vitro* model, which served as a platform for rational therapeutic choice. Here, we show that EC barrier is reduced by the addition of COVID-19 patient plasma in a disease severity-dependent manner and that this can be prevented by stabilization with synthetic Ang-1 (Q-peptide) and Slit2-N. Together, our work provides biological insight into the role of the endothelium in SARS-CoV-2 infection, the importance of miRNA as disease- and pathway-specific biomarkers, and the exciting possibility that endothelial barrier stabilizing treatments might hold promise in COVID-19. Moreover, we provide insights into the use of this approach to find therapeutic options that might prove useful in other critical illnesses and emerging infectious diseases where endothelial permeability is central to disease pathophysiology.

## LIMITATIONS

The results of this study should be viewed in the context of its design, sample size, and socio-geographical context (i.e., access to advanced care life support). Variation between studies may represent temporal differences in circulating viral strains or vaccination status (notably, this study was done in a timeframe exclusive of B.1.1.7 [Alpha], B.1.351 [Beta], B.1.617.2 [Delta], and P.1 [Gamma] circulating strains and prior to public vaccination programs), which can impact severity and mortality. Findings from the miRNA atlas and machine learning algorithms should be validated in additional cohorts before implementation in clinical practice. The *in vitro* system used in this manuscript represents a reductionist approach to disease modelling, and as such, vascular cell co-culture or examination of specific endothelial beds (i.e., coronary or pulmonary) may better inform us about microenvironment changes in COVID-19. Putative therapies could be further assessed in co-culture models (e.g., organ on a chip) that better mimic tissue complexity and will require testing in animal models. Nevertheless, our findings provide important knowledge relating to the pathophysiology and risk stratification of betacoronavirus infection and provide avenues for future research in infectious disease.

## PERSPECTIVES (CLINICAL COMPETENCIES/TRANSLATIONAL OUTLOOK)

- Bedside microRNA-based diagnostics could significantly enhance clinical care of those with acute care diseases.
- Vascular stabilizing therapies represent an attractive supplemental modality for treating critical illnesses and emerging infectious diseases where endothelial permeability is central to disease pathophysiology and should be further explored.

## AUTHOR CONTRIBUTIONS

D.G., M.N., R.W., A.S., C.E., S.M., F.B., M.W., M.R., E.F., K.K., C.D., K.H., P.T., J.E.F, and K.L.H. contributed to the study design. D.G., A.S., P.B., S.F., U.T., A.B., C.D., P.T., and K.H. contributed to patient recruitment, data collection, and clinical analysis. D.G., M.N., R.W., C.E., and S.M. contributed to wet-lab data generation and *in vitro* analysis. S.F., and C.M. contributed to clinical data analysis, machine learning model construction, model testing, and model comparisons. H.H. and M.W. contributed to the mRNA sequencing analysis. D.G., M.N., R.W., C.E., J.E.F, and K.L.H. wrote and revised the manuscript. All authors discussed and commented on the manuscript. K.H., P.T., J.E.F., and K.L.H. provided funding to support this study.

## Data Availability

All data produced in the present study are available upon reasonable request to the authors.

## ACKNOWLEDGEMENTS

We would thank Katherine Tsang for her support in biospecimen collection and consent facilitation. This work utilized infrastructure generously provided by the Peter Munk Cardiac Centre Biobank and the University Health Network’s PRESERVE-Pandemic Response Biobank for coronavirus samples, University Health Network Biospecimen Services, REB # 20-5364. In addition, we would like to acknowledge all those responsible for the collection of biospecimens at St. Michael’s Hospital through the COVID-19 Longitudinal Biomarkers of Lung Injury Study (NCT04747782, REB 20-078). D.G. is funded by a Banting and Best Canadian Graduate Doctoral Scholarship and a Ted Rogers Centre for Heart Research studentship. Coronavirus Disease 2019 research in the laboratories of K.H., P.T., J.E.F., and K.L.H is supported by an innovation grant from the Peter Munk Cardiac Center and an innovation grant from the Ted Rogers Center for Heart Research. U.T., C.D., and A.B., received funding from the St. Michael’s Hospital Foundation through an internal competitive grant, the Canadian Institutes of Health Research and COVID-19 Immunity Task Force (COVID-19 Opportunity Grant), and salary support through the Dorothy and Robert Pitt Research Chair in Acute and Emergency Medicine. K.C.K. is supported by a Tier 1 Canada Research Chair, a CIHR Foundation grant (FDN-148439), a CIHR COVID-19 grant (VR3-172649), and a FAST grant. M.W. is supported by a Tier 2 Canada Research Chair in Comparative Genomics. H.H. was supported by Genome Canada and a Genome Canada, Genomics Technology Platform grant. P.T. is supported by a Tier 2 Canada Research Chair in Cardio-Oncology. Research in the laboratory of J.E.F is supported by Project Grants from the Canadian Institutes of Health Research (Grant: PJT148487 and PJT173489); and infrastructure funding from the Canada Foundation for Innovation, the John R. Evans Leaders Fund, and the Ontario Research Fund. J.E.F. is supported by a Tier 2 Canada Research Chair in Vascular Cell and Molecular Biology from the Canadian Institutes of Health. Research in the laboratory of K.H. is supported by a CIHR Project Grant (PJT178006), the Wylie Scholar Program, Peter Munk Cardiac Centre, University Health Network, and the Toronto Academic Vascular Surgeons. We would also like to gratefully acknowledge the participants who graciously contributed to the study, teams of research coordinators, as well as the clinicians, nurses, and biobank members who assisted with the collection of samples that were utilized in this study. Parts of figures (including the entire graphical abstract) was created with BioRender.com.

## NONSTANDARD ABBREVIATIONS AND ACRONYMS

Ang-2: Angiopoietin-2
ARDS: Acute respiratory distress syndrome
AUROC: Area under the receiver operating characteristic
BMI: Body mass index
COVID-19: Coronavirus Disease 19
CVD: Cardiovascular disease
EC: Endothelial cell
sE-Selectin: Soluble E-selectin
ET-1: Endothelin-1
FDR: False discovery rate
Hs-cTnI: High-sensitivity cardiac troponin
ICU: Intensive care unit
IL: Interleukin
IQR: Interquartile range
miR/miRNA: MicroRNA
MPO: Myeloperoxidase
S.D.: Standard deviation
SARS-COV-2: Severe acute respiratory syndrome coronavirus 2
sICAM-1: Soluble intercellular adhesion molecule-1
sTREM-1: Soluble triggering receptor expressed on myeloid cells-1
sVCAM-1: Soluble vascular cell adhesion molecule-1
VE-cadherin: Vascular endothelial-cadherin

