## Supplemental Materials, Tables, and Figures for "Cardiovascular Signatures of COVID-19 Predict Mortality and Identify Barrier Stabilizing Therapies"

This appendix has been provided by the authors to give readers additional information about their work.

**AFFILIATIONS:** [a]Toronto General Hospital Research Institute, University Health Network, Toronto, Canada; [b]Department of Laboratory Medicine and Pathobiology, University of Toronto, Toronto, Canada; [c]Program in Genetics and Genome Biology, The Hospital for Sick Children, Toronto, ON, Canada; [d]Peter Munk Cardiac Centre, Toronto General Hospital, University Health Network, Toronto, ON, Canada; [e]Johns Hopkins School of Medicine, Baltimore, USA; [f]Institute of Biomedical Engineering, University of Toronto, Toronto, Ontario, M5S 3G9 Canada; [g]Department of Molecular Genetics, University of Toronto; [h]Interdepartmental Division of Critical Care and Institute of Medical Sciences, University of Toronto, Toronto, Canada; [i]Institute of Medical Science, University of Toronto, Toronto, Canada; [j]Keenan Research Center for Biomedical Research, Unity Health Toronto, Toronto, ON, Canada; [k]Critical Care Department, Galilee Medical Center, Nahariya, Israel; [l]Joint Department of Medical Imaging, University Health Network, University of Toronto, Toronto, Canada; [m]Techna Institute, University Health Network, Toronto, Canada; [n]Department of Medical Biophysics, University of Toronto, Toronto, Canada; [o]Vector Institute, University of Toronto, Toronto, Canada; [p]Ted Rogers Program in Cardiotoxicity Prevention, Toronto General Hospital, Toronto, Canada; [q]Division of Vascular Surgery, Department of Surgery, University of Toronto, Toronto, Canada, Toronto, Canada

#### TABLE OF CONTENTS

**SUPPLEMENTAL ABBREVIATIONS AND ACRONYMS ... 5**

**SUPPLEMENTAL METHODS ... 7**

Rationale and Design ... 7

Patient Categorization ... 7

Study Population ... 7

Clinical Data Collection ... 8

Processing of Bloodwork ... 8

Protein Biomarker Analysis ... 8

Cell Culture ... 9

xCelligence Real-Time Cell Analysis (Transendothelial Electrical Resistance [TEER]) ... 9

Transwell Leak Assay ... 9

Bulk RNA-Seq ... 10

HTG EdgeSeq MicroRNA (miRNA) Whole Transcriptome Assay (WTA) from Plasma ... 11

HTG EdgeSeq MiRNA Analysis ... 11

Simultaneous Multiplexed In Vitro Immunofluorescence ... 11

Data Visualization and Statistical Analysis ... 12

Data Deposit ... 13

**SUPPLEMENTAL TABLES ... 14**

Online Table I. Key Reagents and Resources ... 14

Online Table II. Antibodies ... 16

Online Table III. Admission Clinical Laboratory Findings ... 17

Online Table IV. Association of Baseline Clinical Characteristics to Mortality Amongst All Admitted COVID-19 Patients. ... 18

Online Table V. Intensive Care Unit-Level Clinical Characteristics and Clinical Outcomes ... 20

Online Table VI. Association of Baseline Clinical Characteristics to Mortality Amongst All Severe COVID-19 Patients. ... 21

**SUPPLEMENTAL FIGURES ... 23**

Online Figure I; Related to Methods and Table 1: Flow diagram of patients enrolled between the COLOBILI Study (St Michael's) and the COVID Study (University Health Network). ... 23

Online Figure II; Related to Methods and Table 1: Spike (trimer) antigen serology testing from patients having a negative SARS-CoV-2 polymerase chain reaction result. ... 24

Online Figure III; Related to Figure 1: The association of coronary artery disease with mortality characterized in terms of proportion of deceased patients stratified by status. ... 25

Online Figure XXI; Related to Figure 6: Endogenous sSlit2 is upregulated in severe COVID-19 patient plasma. ... 48

**SUPPLEMENTAL DATA FILE ANNOTATIONS ... 49**

Supplementary Data File I. Quality control table for all RNA-sequencing experiments used in this study. ... 49

Supplementary Data File II. R documentation file for the analysis of the RNA-sequencing experiments. ... 49

Supplementary Data File III. R code for the analysis of the RNA-sequencing experiments. .. 49

Supplementary Data File IV. Full list of differentially expressed miRNA with pairwise comparisons between COVID-19 cohorts and the negative controls. ... 49

Supplementary Data File V. Full list of pathway enrichments for miRNA-sequencing experiment between COVID-19 cohorts and the negative controls. ... 49

Supplementary Data File VI. Full list of differentially expressed genes with pairwise comparisons between COVID-19 cohorts and the negative controls. ... 49

Supplementary Data File VII. Full list of pathway enrichments for mRNA-sequencing experiment between COVID-19 cohorts and the negative controls. ... 49

Supplementary Data File VIII. Gene set enrichment analysis for mRNA-sequencing experiment between COVID-19 cohorts and the negative controls. ... 49

**SUPPLEMENTAL REFERENCES ... 50**

### SUPPLEMENTAL ABBREVIATIONS AND ACRONYMS

| | |
|---|---|
| **ACE** | Angiotensin-converting-enzyme inhibitors |
| **Ang-2** | Angiopoietin-2 |
| **APACHE** | Acute physiologic assessment and chronic health evaluation |
| **ARBs** | Angiotensin II receptor blockers |
| **ARDS** | Acute respiratory distress syndrome |
| **BMI** | Body mass index |
| **CAD** | Coronary artery disease |
| **Cat#** | Catalogue number |
| **CCB** | Calcium channel blocker |
| **CF** | Cystic fibrosis |
| **CKD** | Chronic kidney disease |
| **COPD** | Chronic obstructive pulmonary disease |
| **COVID-19** | Coronavirus Disease 2019 |
| **CV** | Cardiovascular |
| **DAPI** | 4′,6-diamidino-2-phenylindole |
| **EBM** | Endothelial basal media |
| **ECMO** | Extracorporeal membrane oxygenation |
| **ED** | Emergency department |
| **EGM** | Endothelial growth medium |
| **ET-1** | Endothelin-1 |
| **FDR** | False discovery rate |
| **FITC** | Fluorescein isothiocyanate |
| **$FiO_2$** | Fraction of inspired oxygen |
| **GERD** | Gastroesophageal reflux disease |
| **GSEA** | Gene set enrichment analysis |
| **GSE** | Gene expression omnibus series accession number |
| **Hs-cTnI** | High-sensitivity cardiac troponin |
| **Ig** | Immunoglobulin |
| **IL** | Interleukin |
| **IQR** | Interquartile range |
| **kDa** | Kilodalton |
| **KEGG** | Kyoto encyclopedia of genes and genomes |
| **miR/miRNA** | MicroRNA |
| **MIV** | Mechanical ventilation |
| **MPO** | Myeloperoxidase |
| **NIV** | Non-invasive ventilation |
| **NSTEMI** | Non-ST-elevation myocardial infarction |
| **OSA** | Obstructive sleep apnea |
| **PAD** | Peripheral artery disease |
| **PF** | Ratio of arterial oxygen partial pressure to fractional inspired oxygen |
| **PFO** | Patent foramen ovale |
| **pHUVEC** | Pooled human umbilical vein endothelial cells |
| **QC** | Quality control |

| | |
|---|---|
| **REB** | Research ethics board |
| **RR** | Respiratory rate |
| **RRID:AB** | Research resource identifier (antibody) |
| **RTCA** | Real-time cell analyzer |
| **S.D.** | Standard deviation |
| **SARS-COV-2** | Severe acute respiratory syndrome coronavirus-2 |
| **sICAM-1** | Soluble intercellular adhesion molecule-1 |
| **SOFA** | Sequential organ failure assessment |
| **sSLIT2** | Soluble slit guidance ligand 2 |
| **sTREM-1** | Soluble triggering receptor expressed on myeloid cells-1 |
| **sVCAM-1** | Soluble vascular cell adhesion molecule-1 |
| **TIA** | Transient ischemia attack |
| **TC** | Tissue culture |
| **TEER** | Transendothelial electrical resistance |
| **TIA** | Transient ischemic attack |
| **TNFα** | Tumor necrosis factor alpha |
| **VTE** | Venous thromboembolism |
| **WBCs** | White blood cells |

### SUPPLEMENTAL METHODS

Rationale and Design: Coronavirus Disease 2019 (COVID-19) is a leading infectious etiology currently present throughout the world, for which there are limited therapeutic interventions. Understanding the pathobiology of COVID-19 outcomes can enable personalized patient management protocols, improve survival, and aid in the preparation of diagnostic tools for future pandemics. Due to the dynamic nature of clinical care guidelines, clinical care did not follow a standardized protocol but rather was determined by individual providers and patient needs. Plasma of enrolled patients was taken at presentation and at set intervals (days 2-3 [$t_{2\text{-}3}$], 4-5 [$t_{4\text{-}5}$], and 6-7 [$t_{6\text{-}7}$]) following obtainment of consent. Comprehensive, integrated analysis of plasma transcriptome data was performed to prioritize COVID-19 outcome signals.

Patient Categorization: After retrospective clinical adjudications of medical records and triaging protocols, patients were assigned to pre-defined clinical groups, centered around the National Institutes of Health, 'Clinical Spectrum of SARS-CoV-2 Infection'[1], those being: (i) severe acute respiratory syndrome coronavirus (SARS-CoV-2) negative patients with mild acute respiratory disease, (ii) mild COVID-19, (iii) moderate COVID-19, (iv) severe COVID-19, and (v) SARS-CoV-2 negative patients requiring intensive care unit (ICU) level management for severe respiratory illness. These assignments were made solely on the basis of information available in the medical record and were blind to any novel biomarker data, which had not yet been generated. For biomarker studies, where appropriate, patients were matched for age, sex, body mass index (BMI), and co-existing conditions with non-infected controls as reference groups.

Study Population*:* This is a multicenter, secondary analysis of a prospectively recruited longitudinal cohort study enrolling consecutive patients with suspected SARS-CoV-2 infection who were referred to two Canadian quaternary care networks in Toronto, Canada from May 2020 to December 2020: University Health Network and St. Michael's Hospital. All participants who were 18 years of age or older, provided either direct written informed consent or were consented into the study by a lawfully entitled substitute decision-maker on behalf of a participant when lacking the capacity to make the decision. The study, and consenting, was conducted in accordance with protocols approved by the Research Ethics Board (REB) of the University Health Network (REB#: 20-5453.6; Cardiovascular Disease and Outcomes among Patients with SARS-CoV-2 Infection During Admission and Post-Discharge [The COVID study]) or St. Michael's Hospital (REB#: 20-078; COVID-19 Longitudinal Biomarkers in Lung Injury [COLOBILI]). SARS-CoV-2-negative patients with severe respiratory illness symptoms were enrolled within the COLOBILI study. Diagnoses of COVID-19 were confirmed through real-time reverse transcription-polymerase chain reaction assays of nasopharyngeal swabs according to the Public Health Ontario guidelines for SARS-CoV-2 testing.[2] The timeframe for recruitment largely excludes community level spread for the variants of concern (i.e., B.1.1.7 [Alpha], B.1.351 [Beta], B.1.617.2 [Delta], and P.1 [Gamma]), with the assumption that all patients harbored one of the predominant unmutated G strains (i.e., GR, GH, and GV).[3] Samples were collected prior to the initiation of public vaccination programs in Ontario, with all patients assumed to be unvaccinated. Admitted patients were followed up after their COVID-19 diagnosis, with all causes of in-hospital mortality, complications, and therapeutic regimen ascertained until discharge. Patients' data were extracted from the in-hospital electronic medical records, de-identified, and assigned random identification numbers which were used throughout the project. Information on sex, age, and pertinent clinical parameters for the cohorts are provided (Table 1). The laboratory parameters were collected as reported by the individual centers, with standard international reference ranges applied to decide the cutoff point for abnormal levels.

Clinical Data Collection: Clinical characteristics, medical history, therapeutics administered during admission, complications, and outcomes were obtained and extracted from electronic medical records by clinical coordinators. Participant disease severity was quantified according to the National Institutes of Health Clinical Spectrum of SARS-CoV-2 infection which is largely based on respiratory parameters. Complete follow-up was available only for those requiring admission to hospital. Obesity was defined as BMI ≥30 kg/m$^2$ in line with World Health Organization guidelines.[4] Preexisting cardiovascular disease (i.e., atrial fibrillation, arterial hypertension, coronary artery disease, dyslipidemia, diabetes mellitus, chronic obstructive pulmonary disease, or heart failure), myocardial infarction, and concurrent active malignancy were defined and recorded from the electronic medical records at the discretion of the clinical coordinators upon availability from previously recorded histories. Smoking was collected as a self-reported variable, with social smoking being defined as less than <4 times per week, and avid smoker being anything greater than that. Cardiovascular complications were defined as a surrogate of new-onset atrial or ventricular arrhythmia (atrial fibrillation, atrial flutter, non-sustained ventricular tachycardia, sustained ventricular tachycardia), acute coronary syndrome, clinical heart failure, right ventricular failure, left ventricular failure, biventricular failure, moderate or greater pulmonary embolism, tamponade, stroke, acute non-coronary ischemia due to hypercoagulable state, or intracardiac thrombus. In patients admitted to the ICU the Acute Physiologic Assessment and Chronic Health Evaluation II and the Sequential Organ Failure Assessment scores were used.[5,6] These are validated methods for grading the severity of illness in critically ill patients based upon point-scoring systems associated with the degree of dysfunction with specific physiologic variables.

Processing of Bloodwork*:* Peripheral blood samples were collected synchronously with standard of care bloodwork at the University Health Network or St. Michael's Hospital between May 2020 and January 2021. Peripheral blood samples (10 mL) were drawn from the cubital vein into BD Vacutainer® Blood Collection Tubes (BD Bioscience, Franklin Lakes, NJ) containing $K_2EDTA$ and processed within three hours. Plasma was separated from whole blood through centrifugation (2,000×g, 24°C, 15 min) and stored at -80°C until downstream processing. At no time during the process was the plasma subjected to temperatures below 4°C or above 25°C. Samples were thawed on ice and the plasma was subjected to sequential centrifugation of (2,500×g, 4°C, 25 min) to reduce platelets and large particulate. Hemolysis was examined prior to downstream analysis by measuring the absorbance at 414 nm using a DS-11$^+$ Spectrophotometer (DeNovix, Wilmington, Delaware, United States).

Protein Biomarker Analysis*:* Circulating levels of angiopoietin-2 (Ang-2; Lower limit of quantification [LLOQ] 9.91 pg/mL), soluble CD62 antigen-like family member E (sE-Selectin; LLOQ 4.22 pg/mL), soluble CD54/intercellular adhesion molecule-1 (sICAM-1; LLOQ 4.1 pg/mL), soluble CD106/vascular cell adhesion protein-1 (sVCAM-1; LLOQ 137 pg/mL), CD105/endoglin, endothelin-1 (ET-1; LLOQ 0.250 pg/mL), interleukin-6 ([IL]-6; LLOQ 0.41 pg/mL), IL-8 (LLOQ 0.19 pg/mL), and soluble triggering receptor expressed on myeloid cells-1 (sTREM-1; LLOQ 4.19 pg/mL) were quantified in platelet free plasma samples using the Simple

Plex Ella (ProteinSimple, San Jose, CA, USA) multiplex platform according to the manufacturer's instructions; all Simple Plex values are reported as the average of triplicate readings. Soluble Slit homolog 2 protein (sSLIT2) was quantified using an enzyme-linked immunosorbent sandwich assay (ELISA; LLOQ 0.10 ng/mL, Elabscience, Wuhan, China). Myeloperoxidase was additionally quantified through ELISA (LLOQ; 0.062 ng/mL; R&D Systems Minneapolis, MN, USA). Circulating cardiac troponin I (cTnI) was quantified using a CLIA certified high sensitivity ELISA kit (LLOQ 0.92 pg/mL; Biomatik, Kitchener, ON, CAN). Cardiac injury was defined as plasma levels of high-sensitivity cTnI (hs-cTnI) greater than the 99$^{th}$ percentile of normal values, as per clinical guidelines. See Online Table I for reagents.

Cell Culture*:* Pooled human umbilical vein endothelial cells ([pHUVECs], Lonza, Basel, Switzerland) from multiple donors were cultured in EC Growth Basal Medium-2 (Lonza, Basel, Switzerland) containing the complete EC Growth Medium BulletKit$^{TM}$ (Lonza, Basel, Switzerland) at 37°C in a 5% $CO_2$ humidified incubator. Cells were maintained on 10 cm tissue culture plates (Corning, Costar, New York, NY, USA) coated with 0.1% (v/v) gelatin attachment factor (ThermoFisher, Waltham, MA, USA) and passaged every 2-4 days at 70%-80% confluency. For experimentation, pHUVECs at passages 3-7 were used. Cells tested negative for the presence of mycoplasma contamination (ThermoFisher, Waltham, MA, USA).

xCelligence Real-Time Cell Analysis (Transendothelial Electrical Resistance [TEER]): Baseline resistance measurements were acquired using 150 µL EC Growth Basal Medium-2 for 15 minutes prior to experiment initiation using xCelligence Real-Time Cell Analyzer (RTCA, ACEA Biosciences, San Diego, CA). Following this, pHUVECs were seeded at densities of $4x10^4$/well into multiple 16-well E-plates (ACEA Biosciences, San Diego, CA) in volumes of 150 µL EC Growth Basal Medium-2 and monitored on the xCelligence RTCA. Once the cells reached confluency, 20% v/v plasma or 2 U/mL thrombin (Sigma-Aldrich, St Louis, MO) was added, and the cells incubated at 37°C in a 5% carbon dioxide atmosphere for six hours. To quantify the change in permeability, cell indexes (surrogate metric for change in resistance across the monolayer) were adjusted to the resistance of reference wells (i.e., untreated cells in growth media) and normalized to the time point immediately prior to the addition of the plasma. The net area between the curve of the normalized dataset was utilized to calculate the overall change in permeability, with an increased area representing increases in TEER and decreases representing declines in TEER (i.e., loss of endothelial barrier function). During therapeutic testing, cells were co-treated with 400 µM QHREDGS peptide (Q-Peptide)[7], 1280 ng/mL recombinant Slit2-N[8], 50 µg/mL nangibotide[9], 3 µM dexamethasone[10], or a matching DMSO control with Dulbecco's Phosphate Buffered Saline without magnesium and calcium ([PBS$^{-/-}$], Gibco, Gaithersburg, MD, USA.

Transwell Leak Assay*:* Endothelial monolayer leak assays were performed as previously described.[11] Briefly, pHUVECs were seeded on 3 µm pore transwell inserts (Corning Life Sciences, Corning, NY, USA) coated with 0.1% (v/v) gelatin attachment factor (ThermoFisher, Waltham, MA, USA). The cells were grown on the inserts for two days until reaching 90%-95% confluency. Cells were subsequently treated for either one hour (acutely) *or* six hours (sub-chronically) with either 20% (v/v) human plasma or control PBS$^{-/-}$. Prior to leak quantification, cell media was changed to Hanks Buffered Salt Solution (ThermoFisher, Waltham, MA, USA) and 1 mg/mL 40 kilodalton (kDa) fluorescein isothiocyanate–dextran (FITC-dextran, Sigma-

Aldrich, St Louis, MO) was added to the top chamber with tracer flux subsequently being allowed to occur for one hour; 2 U/mL thrombin was added as a positive control to certain wells at the time of FITC addition. The experimenter was blinded to the grouping of each sample. The FITC accumulation in the bottom chamber was assessed in triplicate via excitation at 485 nm and emission at 535 nm using a Biotek Cytation 5 (Biotek, Winooski, VT, USA).

Bulk RNA-Seq*:* Total RNA was isolated from treated pHUVEC samples (n=5 control [$PBS^{-/-}$], n=5 mild SARS-CoV-2 negative, n=5 mild disease, n=5 moderate disease, and n=5 severe disease) at the six-hour timepoint using the RNeasy Plus Micro kit (Qiagen, Germantown, MD, USA), after washing twice with ice-cold $PBS^{-/-}$. RNA quantities and quality were assessed using the Agilent 2100 Bioanalyzer (Agilent Technologies, Mississauga, ON, Canada). All samples passed a quality control threshold (RNA integrity number ≥ 7.0) to proceed to library preparations and RNA-seq. A total amount of 20 ng RNA per sample was used as input material for the RNA sample preparations. Sequencing libraries were generated using NEBNext® UltraTM RNA Library Prep Kit for Illumina® (NEB, Ipswich, MA, USA) following the manufacturer's recommendations and index codes were added to attribute sequences to each sample. Briefly, mRNA was purified from total RNA using poly-T oligo-attached magnetic beads. Fragmentation was carried out using divalent cations under elevated temperature in NEBNext First Strand Synthesis Reaction Buffer (5X). First-strand cDNA was synthesized using random hexamer primer and M-MuLV Reverse Transcriptase (RNase H-). Second strand cDNA synthesis was subsequently performed using DNA Polymerase I and RNase H. Remaining overhangs were converted into blunt ends via exonuclease/polymerase activities. After adenylation of 3' ends of DNA fragments, NEBNext Adaptor with hairpin loop structure were ligated to prepare for hybridization. To select cDNA fragments of ~150-200 bp in length, the library fragments were purified with AMPure XP system (Beckman Coulter, Beverly, USA). Then 3 μL USER Enzyme (NEB, Ipswich, MA, USA) was used with size-selected, adaptor-ligated cDNA at 37 °C for 15 minutes followed by five minutes at 95 °C before PCR. PCR was performed with Phusion High-Fidelity DNA polymerase, Universal PCR primers and Index (X) Primer. The resulting PCR products were purified (AMPure XP system) and library quality was assessed on the Agilent Bioanalyzer 2100 system. Sequencing was carried out on an Illumina NovaSeq® 6000 (NovoGene; Illumina, San Diego, California, United States), using paired-end 2×150 bp chemistry at a depth of 20 million reads per sample.

Sequencing quality was examined using FastQC[12] (v0.11.2), and adaptors were subsequently trimmed with Trimmomatic[13] (v0.36) (in paired-end mode, parameters: TruSeq3-PE-2.fa:2:30:7:8:true LEADING:10 TRAILING:10 SLIDINGWINDOW:5:20 MINLEN:36). Trimmed fastq files were subsequently re-examined with FastQC again to ensure efficient adaptor and quality trimming. Reads were aligned to the hg38 genome (obtained through UCSC) with STAR[14] (v2.7.8a) using the default parameters only for quality control purposes. Aligned reads were subjected to the following quality control metrics: (i) duplication rate assessment through Picard Markduplicates (v.2.18), (ii) rRNA content, genomic read distribution and 3'-5' bias through RNA-SeQC (v.2.4.0). These metrics, together with the read alignment summary from STAR, were summarized using MultiQC[15] (v1.9), resulting in the removal of one sample on the basis of abnormal read duplication rates (n=1 mild disease). Gene quantification was performed using Salmon[16] (v1.3.0) with adaptor-trimmed reads and gene annotation obtained from GENCODE version 31 (parameters: --gcBias –validateMappings). Gene-level read counts were imported into R using R package Tximport[13](v.1.18.0) with transcript length adjustment. Read counts were first normalized by DESeq2[17] (v.1.30.0), with the svaseq() function from R package sva (v.3.38.0)[18] being used to subsequently estimate hidden batch effects with n.sv=4. Sva-normalized read counts were obtained by regressing out covariates using the following function: https://github.com/LieberInstitute/jaffelab/blob/master/R/cleaningY.R and were subsequently used for PCA analysis and sample correlation analysis. R package fgsea (v 1.16.0)[19] was used to perform Gene Set Enrichment Analysis with genes ranked based on fold change between the compared conditions. Gene set files were obtained from the Molecular Signatures Database (MSigDB, v7.0) and only C2: curated gene sets and C5: ontology gene sets were used. Pathway enrichment using significantly differentially expressed genes were performed with R package gProfilerR (v.0.7.0)[20]. R scripts and essential data files to reproduce the mRNA-seq analysis can be found in the Online Data Files.

HTG EdgeSeq MicroRNA (miRNA) Whole Transcriptome Assay (WTA) from Plasma*:* Lysis of $t_{0-1}$ plasma aliquots was facilitated by combining 30 µL plasma with equivalent (v/v) amounts of HTG Plasma Lysis Buffer (HTG Molecular, Tucson, AZ, USA) as well as $1/10^{th}$ (v/v) amounts of Proteinase K (HTG Molecular, Tucson, AZ, USA). The mixture was subsequently incubated for three hours at 50°C shaking at 1,400 rpm. From each prepared sample, 35 µL were added per well to a 96-well sample plate. Human fetal brain RNA was added to one well at 25 ng/well to serve as an internal control. Samples were run on an HTG EdgeSeq Processor using the HTG EdgeSeq miRNA WTA (HTG Molecular, Tucson, AZ, USA) to facilitate nuclease protection, whereby a pre-selected miRNA population is protected with proprietary protection probes, followed by degradation of all non-hybridized probes and non-targeted RNA by S1 nuclease. Following the processing, samples were individually barcoded (using a 16-cycle PCR reaction), individually purified using AMPure XP beads (Beckman Coulter, Brea, CA, USA), and quantified using a KAPA Library Quantification kit (KAPA Biosystem, Wilmington, MA, USA). The library was sequenced on a NextSeq (Illumina, Inc., San Diego, CA) using a V3 150-cycle kit with two index reads. PhiX (Roche, Mississauga, ON, CAN) was spiked into the library at 5%; this spike-in control is standard for Illumina sequencing libraries. Data were returned from the sequencer in the form of demultiplexed FASTQ files, with one file per original well of the assay. The HTG EdgeSeq Parser (v. 5.0.535.3181, HTG Molecular, Tucson, AZ, USA) was used to align the FASTQ files to the probe list to collate the data. Data were provided as data tables of raw, quality control (QC) raw, counts per million, and median normalized.

HTG EdgeSeq MiRNA Analysis*:* Samples were initially analyzed using three QC metrics: (i) QC0, examining degraded sample; cut-off of positive % ≥14% as failure, (ii) QC1, insufficient read depth; read depth ≤ 500k as failure, (iii) QC2, minimal expression variability; relative standard deviation of reads ≤ 0.08 as failure. Of the 156 samples sent for sequencing, 12 exhibited failure at the level of QC2 and were excluded from all downstream analyses. Normalization of miRNA expression data on the remaining samples was performed using DeSeq2[17] (v. 1.14.1) in the HTG reveal software (v.3.0.0, HTG Molecular, Tucson, AZ, USA). MiRNAs were considered detectable if they had expression levels of >5 counts per million in more than half of our samples. Expression counts were logarithmically scaled ($\log_{10}$) for data visualization.

Simultaneous Multiplexed *In Vitro* Immunofluorescence: Cells were fixed in ice-cold methanol (Sigma-Aldrich, St Louis, MO) for five minutes at room temperature. Blocking was subsequently conducted in a solution containing 1% BSA (w/v, BioShop, Burlington, ON, CAN), 22.52 mg/mL glycine (ThermoFisher, Waltham, MA, USA), and $PBS^{-/-}$ with Tween 20 (PBST, $PBS^{-/-}$ + 0.1% Tween 20) for 30 minutes. Immunostaining was then conducted with diluted antibody (Online Table II) in 1% BSA PBST for 16 hours at 4°C. The cells were subsequently washed three times in $PBS^{-/-}$, five minutes for each wash, and re-incubated with the concordant secondary antibodies in 1% BSA for one hour at room temperature in the dark. Post-washing, cells were counterstained with Vectashield Antifade Mounting Media with 4′,6-diamidino-2-phenylindole ([DAPI], Vector Laboratories, Burlingame, CA, USA) and mounted with coverslips (VWR International, Mississauga, ON, CAN) and sealed with nail polish. Confocal images were taken using an Olympus Fluoview 1000 Confocal microscope Olympus IX81 inverted stand (Olympus, CA, USA). Fluorochromes were excited using the following wavelengths: 405 nm for DAPI, 473 nm for Alexa Fluor 488, and 559 nm for Alexa Fluor 568 (ThermoFisher, Waltham, MA, USA). A 20X/0.75NA UPLSAPO super apochromat objective was used to take the 20X images while a Plan Apo 40x/1.35 NA oil immersion objective was utilized for the 40X images. Image processing was done with the FV10-ASW 4.2 Viewer (Olympus, CA, USA). Image intensities were calculated using FIJI[21] (v2.1.0/1.53c), by examining the integrated density and dividing that by the number of DAPI positive cells within each image.

Data Visualization and Statistical Analysis*:* All data generated and analyzed which support the findings of this study are included in this article. Associated supplementary information files are available on a publicly accessible archive (see below). *Descriptive Analysis* - Clinical characteristics were characterized using summary statistics. Continuous variables were described using median and inter-quartile range (IQR), and dichotomous or polytomous variables were described using frequencies. Between-group differences were evaluated using Wilcoxon rank-sum tests for continuous variables and Fisher's exact tests for dichotomous/polytomous variables. Correlation between continuous variables were quantified using Spearman rank correlation. *Descriptive outcome analysis* - The Kaplan-Meier survival method was applied to assess the in-hospital death, and the between-group differences in the freedom from death were evaluated using log-rank tests. The length of hospitalization/ICU was characterized using competing risk models in terms of cumulative incidence rate function. Univariable Cox proportional hazard regression were applied to assess and quantify the association of the baseline clinical characteristics with in-hospital/ICU death. The associations of continuous variables were modeled using natural cubic splines. *Biomarker Analysis* - Comparisons between two independent groups were made using *t*-tests for normally distributed continuous variables or Wilcoxon rank-sum tests non-normally distributed continuous variables. When more than two groups were compared, either a one-way ANOVA with a Tukey or Bonferroni post-hoc test (where appropriate) for multiple testing correction, Kruskal-Wallis one-way analysis of variance with Dunn's multiple comparison correction. Two-way ANOVA was used to estimate how the mean quantitative variable changes according to time and group differences in leak experiments. Where appropriate, Benjamini-Hochberg false discovery rate (FDR) was utilized with adjusted P values (or Q value where stated) of $<0.05$ being considered statistically significant and indicated in the graphs as reported by the analysis software with significance thresholds of $P<0.05$, $P<0.01$, $P<0.001$, and $P<0.0001$ indicated as *, **, ***, **** respectively. MiRNA pathway analysis was conducted using BioCarta/KEGG/Reactome databases and tested for enrichment by a hypergeometric test with adjustment for multiple comparisons using the Benjamini-Hochberg FDR, with $P \leq 0.05$ considered to be statistically enriched in a gene set of interest.[22-24] Although many hypotheses were tested throughout the manuscript, no experiment-wide multiple test correction was applied. Unless indicated otherwise, graphs depict averaged values of independent data points with technical replicates and have error bars displayed as mean +/- standard deviation (±S.D.). Data were analyzed with GraphPad Prism 9.0.0 for MacOS (GraphPad Software, Inc., La Jolla, CA, USA; Biomarker Multiple Comparisons), R[25] (v4.0.3; Spearman Correlation Plots), and FIJI[21] (v2.1.0/1.53c; Quantifying Image Intensities). Final figures were assembled for publication purposes using Adobe Illustrator (v25.4.1).

Risk Assessment Using Machine Learning*:* We performed 250 experiments using repeated randomized stratified sub-sampling cross-validation into 80% training and 20% testing using Python (v3.8.8) and scikit-learn[26] (v.0.24.1). Categorical features were hot encoded, with missing variables recorded as additional categorical variables, having -1.0 for numerical features. For each experiment, a Random Decision Forest model was fit to the training dataset and evaluated on the independent testing set.[27] Model performance was assessed by the area under the receiver operating characteristic (AUROC) calculated on the testing set. Average AUROC and 95% confidence intervals were calculated across the 250 runs using the percentile method. Feature importance was estimated using permutation feature importance and aggregated across the 250 runs. The microRNA model had a multiphase selection, whereby all microRNA were inputted into the first phase (feature selection via collinearity), upon which the remaining 102 microRNA were used to train the model.

Data Deposit*:* The data generated in this study have been deposited in the National Center for Biotechnology Information’s Gene Expression Omnibus and are accessible through the GEO Series accession number (GSE; GSE178331, mRNA) and (GSE178246, miRNA).

### SUPPLEMENTAL TABLES

**Online Table I.** Key Reagents and Resources

| REAGENT or RESOURCE | SOURCE | IDENTIFER |
|---|---|---|
| Enzyme-Linked Immunosorbent Assays | | |
| Ang-2, sE-SEL, sICAM-1, sVCAM-1 | ProteinSimple | Cat#: SPCKC-PS-004112 |
| ET-1, IL-6, IL-8, sTREM-1 | ProteinSimple | Cat#: SPCKC-PS-004111 |
| High Sensitivity Cardiac Troponin I | Biomatik | Cat#: EKU09460 |
| MPO | R&D Systems | Cat#: DMYE00B |
| sSlit2 | Elabscience | Cat#: E-EL-H0931 |
| SARS-CoV-2 Spike (Trimer) Ig Total | ThermoFisher | Cat#: BMS2323 |
| Chemicals, Peptides, and Recombinant Proteins | | |
| Bovine Serum Albumin | BioShop | Cat#: ALBC0100 |
| Dexamethasone | BioShop | Cat#: DEX002.100 |
| EBM-2™ | Lonza | Cat#: 00190860 |
| EGM-2™ Bullet Kit | Lonza | Cat#: CC-3162 |
| FITC-40kDa Dextran | Sigma-Aldrich | Cat#: 53379 |
| Gelatin | ThermoFisher | Cat#: S006100 |
| Hanks' Balanced Salt Solution | Thermo | Cat#: 14025092 |
| Methanol | Sigma-Aldrich | Cat#: 322415 |
| Mounting Medium with DAPI | Vectashield | Cat#: H-1200 |
| Nangibotide | LifeTein | Cat#: Custom Order |
| Phosphate Buffered Saline | Gibco | Cat#: LS10010023 |
| Q-Peptide | Genscript | Cat#: SC1208 |
| RIPA Buffer (10x) | EMD Millipore | Cat#: 20-188 |
| Slit2-N | PreproTech | Cat#: 150-11 |
| Thrombin | Sigma-Aldrich | Cat#: 10602400001 |
| TNFα Human | Sigma-Aldrich | Cat#: T0157-10UG |
| TritonX-100 | Sigma-Aldrich | Cat#: T8787 |
| Tween20 | ThermoFisher | Cat#: 003005 |
| UltraPure Glycine | ThermoFisher | Cat#: 15527013 |
| Commercial Components | | |
| Vacutainer EDTA Tubes | BD Diagnostics | Cat#: 367525 |
| E-16 Plates | Agilent | Cat#: 300601150 |
| Black 96 well assay plate | Sigma-Aldrich | Cat#: M0312 |
| Costar TC-Treated Plates (24-well) | Millipore Sigma | Cat#: CLS3527-100EA |
| Coverslips No. 1, 24x50*mm* | VWR | Cat#: 4839081 |
| DNA LoBind Tubes | Eppendorf | Cat#: 22431021 |
| Mycoplasma Contamination Kit | ThermoFisher | Cat#: 4460623 |
| RNeasy Plus Micro Kit | Qiagen | Cat#: 74034 |
| Transwell Filters (12-well plate) | Corning | Cat#: 3462 |
| NEBNext® UltraTM Library Prep Kit | New England Biolabs | Cat#: E7645S |
| AMPure XP Beads | Beckman Coulter | Cat#: A63880 |
| Uracil-Specific Excision Reagent | New England Biolabs | Cat#: M5505S |
| Phusion High-Fidelity DNA polymerase | ThermoFisher | Cat#: F-530XL |
| HTG Lysis Buffer | HTG Diagnostic's | Cat#: SPP-Mi-04 |
| KAPA Library Quantification Kit | Roche | Cat#: 07960140001 |
| PhiX | Illumina | Cat#: FC-110-3001 |
| Cells | | |
| pHUVEC | Lonza | Cat#: C2519A |

| | | |
|---|---|---|
| | | Lot: 661173 |
| | | Cat#: C2519A<br>Lot: 636514 |
| **Key Instruments** | | |
| Cytation 5 | Biotek | Serial Number: 16041913 |
| DS-11 + Spectrophotometer | DeNovix | Serial Number: 760419B |
| ELLA | ProteinSimple | Serial Number: ELLA-16080112 |
| NextSeq | Illumina | Serial Number: A0877 |
| NovoSeq6000 | Illumina | Serial Number: |
| Fluoview 1000 Confocal IX81 | Olympus | Serial Number: 8B03839 |
| Real-Time Cell Analysis | xCelligence | Serial Number: 3211107167871 |
| **Software and Algorithms** | | |
| Adobe Illustrator | https://www.adobe.com/products/illustrator.html (Ver. 25.4.1) | |
| DeSeq2[17] | https://bioconductor.org/packages/release/bioc/html/DESeq2.html (Ver. 3.12) | |
| EdgeSeq Parser | HTG Diagnostics (Ver. 5.0.525.3181) | |
| FastQC[12] | https://www.bioinformatics.babraham.ac.uk/projects/fastqc/ (Ver. 0.11.2) | |
| FIJI[21] | https://imagej.net/software/fiji/downloads (Ver. 2.1.0/1.53c) | |
| FV10-ASW Viewer | https://www.olympus-lifescience.com/es/support/downloads (Ver. 4.2b) | |
| Fgsea (v 1.16.0)[19] | https://github.com/ctlab/fgsea#:~:text=fgsea%20is%20an%20R%2Dpackage,level%20split%20Monte%2DCarlo%20scheme. (Ver. 1.16.0) | |
| gProfilerR | https://biit.cs.ut.ee/gprofiler/ (Ver. 0.7.0) | |
| HTG Reveal | HTG Diagnostics (Ver. 3.0.0) | |
| MarkDuplicates | http://broadinstitute.github.io/picard (Ver. 2.18) | |
| miRPath[28] | http://snf-515788.vm.okeanos.grnet.gr/ (Ver. 3.0) | |
| MultiQC[15] | https://github.com/ewels/MultiQC (Ver. 1.9.0) | |
| Prism 9 | GraphPad Software ((Ver. 9.0.0 (86)) | |
| Python | https://www.python.org/ (Ver. 3.8.8) | |
| Qualimap[29] | https://github.com/EagleGenomics-cookbooks/QualiMap (Ver. 2.2.1) | |
| R[25] | https://www.r-project.org/ (Ver. 4.0.3) | |
| RNA-SeQC[30] | https://github.com/getzlab/rnaseqc (Ver. 2.4.0) | |
| RTCA | xCelligence (Ver. 2.0.0) | |
| Salmon[16] | https://combine-lab.github.io/salmon/ (Ver. 1.3.0) | |
| Scikit-learn[26] | https://scikit-learn.org/stable/ (Ver. 0.24.1) | |
| Simple Plex Runner | Protein Simple (Ver. 3.7.1.12) | |
| STAR[14] | https://github.com/alexdobin/STAR (Ver. 2.7.8) | |
| Sva[18] | https://bioconductor.org/packages/release/bioc/html/sva.html (Ver.3.38.0) | |
| Trimmomatic[13] | https://bioconductor.org/packages/release/bioc/html/tximport.html (Ver. 0.36) | |
| Tximport[13] | https://bioconductor.org/packages/release/bioc/html/tximport.html (Ver. 3.12) | |
| **Deposited Data** | | |
| Plasma microRNA transcriptome | Human participants | GSE: 178246 |
| Messenger RNA sequencing | Pooled HUVECs | GSE: 178331 |

Abbreviations: Ang-2 = Angiopoietin-2; Cat# = Catalogue number; DAPI = 4′,6-diamidino-2-phenylindole; EBM = Endothelial basal media; EGM = Endothelial growth medium; ET-1 = Endothelin-1; FITC = Fluorescein isothiocyanate; pHUVEC = Pooled human umbilical vein endothelial cells; Ig = Immunoglobulin; IL = Interleukin; kDa = Kilodalton; MPO = Myeloperoxidase; RIPA = Radioimmunoprecipitation assay buffer; SARS-CoV-2 = Severe acute respiratory syndrome coronavirus; sICAM = Soluble intercellular adhesion molecule-1; sSLIT-2 = Soluble slit guidance ligand 2; sTREM-1 = Soluble triggering receptor expressed on myeloid cells-1; sVCAM-1 = Soluble vascular cell adhesion molecule-1; TC = Tissue culture; TNFα = Tumor necrosis factor alpha.

**Online Table II.** Antibodies

| ANTIGEN | HOST | SOURCE | IDENTIFER |
|---|---|---|---|
| Human Vascular Endothelial-Cadherin | Mouse | R&D Systems | Cat#: 9381; RRID:AB_2260374 |
| Polyclonal Claudin-5 | Rabbit | Invitrogen | Cat#: 34-1600; RRID:AB_2533157 |
| Alexa Fluor 488 Anti-rabbit | Goat | ThermoFisher | Cat#: A11008; RRID:AB_143165 |
| Alexa Fluor 568 Anti-mouse | Donkey | ThermoFisher | Cat#: A10042; RRID:AB_2534017 |

Abbreviations: Cat# = Catalogue number; Research Resource Identifier (Antibody) = RRID:AB.

**Online Table III.** Admission Clinical Laboratory Findings

| Characteristics* | Disease Severity | | | | | |
|---|---|---|---|---|---|---|
| | Mild Negative (n=30) | Mild (n=27) | Moderate (n=39) | Severe (n=76) | Severe Negative (n=69) | P-value |
| Complete Blood Count | | | | | | |
| White blood cells – million/mm$^3$ | | | | | | |
| Median (IQR) | 8.0 (6.1-10.0) | 7.0 (5.2-11.2) | 7.0 (5.1-9.1) | 10.8 (8.3-13.8) | 11.9 (7.4-16.9) | **<0.001** |
| Distribution – no./total no. (%) | | | | | | |
| <4 | 3/30 (10.0) | 4/27 (14.8) | 4/36 (11.1) | 2/75 (2.7) | 4/69 (5.8) | 0.1954 |
| >11 | 3/30 (10.0) | 7/27 (25.9) | 6/36 (16.7) | 35/75 (46.7) | 36/69 (52.2) | **<0.0001** |
| Neutrophils – per mm$^3$ | | | | | | |
| Median (IQR) | 5.1 (3.8-7.0) | 4.7 (3.6-8.2) | 4.7 (3.2-6.5) | 9.0 (6.0-12.0) | 9.5 (6.0-15.0) | **<0.001** |
| <3,000 | 4/30 (13.3) | 5/27 (18.5) | 8/36 (22.2) | 1/75 (1.3) | 3/68 (4.4) | **0.0009** |
| >5,800 | 11/30 (36.7) | 10/27 (37.0) | 12/36 (33.3) | 22/75 (29.3) | 14/68 (20.6) | 0.3613 |
| Lymphocytes – per mm$^3$ | | | | | | |
| Median (IQR) | 1.4 (1.0-1.9) | 1.3 (0.9-2.1) | 1.2 (0.8-1.7) | 0.9 (0.6-1.2) | 0.9 (0.6-1.5) | **0.0023** |
| Distribution – no./total no. (%) | | | | | | |
| <1,500 | 16/30 (53.3) | 17/27 (63.0) | 25/36 (69.4) | 61/75 (81.3) | 52/68 (76.5) | **0.0343** |
| >3,000 | 1/30 (3.3) | 1/27 (3.7) | 2/36 (5.6) | 3/75 (4.0) | 4/68 (5.9) | 0.9693 |
| Monocytes – per mm$^3$ | | | | | | |
| Median (IQR) | 0.6 (0.4-0.8) | 0.6 (0.4-0.8) | 0.6 (0.3-0.7) | 0.5 (0.3-0.8) | 0.8 (0.3-1.0) | 0.5077 |
| Distribution – no./total no. (%) | | | | | | |
| <300 | 1/30 (3.3) | 4/27 (14.8) | 5/36 (13.9) | 14/75 (18.7) | 21/68 (30.9) | **0.0180** |
| >500 | 16/30 (53.3) | 15/27 (55.6) | 18/36 (50.0) | 32/75 (42.7) | 42/68 (61.8) | 0.2459 |
| Platelet count – thousand/mm$^3$ | | | | | | |
| Median (IQR) | 232 (179-292) | 247 (177-273) | 246 (179-292) | 227 (154-313) | 217 (151-268) | 0.6530 |
| Distribution – no./total no. (%) | | | | | | |
| <150,000 | 2/30 (6.7) | 4/27 (14.8) | 3/36 (8.3) | 17/75 (22.7) | 17/69 (24.6) | 0.1117 |
| >400,000 | 2/30 (6.7) | 1/27 (3.7) | 5/36 (13.9) | 6/75 (8.0) | 5/69 (7.2) | 0.6457 |
| Cardiac Laboratory Results | | | | | | |
| hs-CnT – ng/mL, median (IQR) | 2.50 (0.92-6.20) | 0.92 (0.92-6.50) | 1.90 (0.92-7.66) | 16.4 (7.42-56.0) | 23.0 (10.0-62.5) | **<0.0001** |
| Coagulation Laboratory Results | | | | | | |
| International normalized ratio | | | | | | |
| <0.9 | 0/11 (0.0) | 0/5 (0.0) | 0/17 (0.0) | 0/74 (0.0) | 0/65 (0.0) | 1.0000 |
| >1.1 | 5/11 (45.5) | 4/5 (80.0) | 12/17 (70.6) | 47/74 (63.5) | 45/65 (69.2) | 0.5329 |
| Partial thromboplastin time | | | | | | |
| <25 | 6/8 (75.0) | 2/5 (40.0) | 2/14 (14.3) | 0/74 (0.0) | 0/64 (0.0) | **<0.0001** |
| >40 | 0/8 (0.0) | 1/5 (20.0) | 3/14 (21.4) | 10/74 (13.5) | 4/64 (6.3) | 0.2930 |
| Supplemental Laboratory Results | | | | | | |
| Albumin, g/liter | | | | | | |
| <35 | 2/5 (40.0) | 4/9 (44.4) | 8/17 (47.1) | 59/74 (79.7) | 44/69 (63.8) | **0.0136** |
| >50 | 0/5 (0.0) | 0/9 (0.0) | 0/17 (0.0) | 1/74 (1.4) | 0/69 (0.0) | 0.8513 |
| Alanine aminotransferase, >40 U/liter | 1/14 (7.1) | 5/18 (27.8) | 3/24 (12.5) | 25/72 (34.7) | 17/66 (25.8) | **0.0791** |
| Aspartate aminotransferase, >40 U/liter | 3/14 (21.4) | 5/18 (27.8) | 4/24 (16.7) | 41/72 (56.9) | 26/66 (39.4) | **0.0019** |
| Creatine kinase, ≥200 U/liter | 2/13 (15.4) | 2/5 (40.0) | 3/36 (8.3) | 54/74 (73.0) | 35/63 (55.6) | **<0.0001** |
| Creatinine, ≥133 µmol/liter | 1/30 (3.3) | 4/27 (14.8) | 6/36 (16.7) | 23/75 (30.7) | 20/69 (29.0) | **0.0149** |
| D-dimer, ≥0.5 mg/liter | 3/8 (37.5) | 9/10 (90.0) | 10/12 (83.3) | 26/49 (53.1) | 8/56 (14.3) | **<0.0001** |
| Lactate Dehydrogenase – µkatals/liter | | | | | | |
| <2.34 | 4/8 (50.0) | 12/15 (80.0) | 8/12 (66.7) | 4/38 (10.5) | 1/60 (1.7) | **<0.0001** |
| >4.68 | 0/8 (0.0) | 0/15 (0.0) | 1/12 (8.3) | 5/38 (13.2) | 11/60 (18.3) | 0.2629 |
| Total bilirubin, >17.1 µmol/liter | 2/13 (15.4) | 1/16 (6.3) | 0/17 (0.0) | 17/71 (23.9) | 12/68 (17.6) | 0.1047 |
| Minerals, median (IQR) – mmol/liter | | | | | | |
| Potassium | 3.0 (3.8-4.4) | 3.9 (3.7-4.3) | 4.1 (3.7-4.4) | 4.2 (3.8-4.3) | 4.1 (3.7-4.8) | 0.4223 |
| Sodium | 139 (137-140) | 137 (136-141) | 138 (135-140) | 138 (135-143) | 137 (135-140) | 0.5582 |

* Summary statistics, where n-value is not provided, are based on the full population indicated in the column heading.

Bolded log-ranked P values are significant (P values <0.05). Abbreviations: hs-cTnI = High sensitivity cardiac troponin I; IQR = Interquartile range. Percentages may not add up to 100% due to rounding.

**Online Table IV.** Association of Baseline Clinical Characteristics to Mortality Amongst All Admitted COVID-19 Patients.

| Variables (Baseline) | Unadjusted Log-Rank P Value |
|---|---|
| Comorbidity - Gout | **0.003** |
| MiRNA - hsa-miR-6080 | **0.009** |
| Comorbidity - Coronary artery disease | **0.010** |
| MiRNA - Ang-2 | **0.015** |
| MiRNA - hsa-miR-199a-3p | **0.017** |
| MiRNA - hsa-miR-4793-5p | **0.027** |
| MiRNA - hsa-miR-181a-5p | **0.028** |
| MiRNA - hsa-miR-mir-30b-5p | **0.035** |
| MiRNA - hsa-miR-6750-5p | **0.036** |
| Clinical - Age of patient at hospital admission | **0.046** |
| Protein - MPO | 0.079 |
| History of stroke | 0.115 |
| Comorbidity - Heart failure | 0.149 |
| Comorbidity - Obesity | 0.151 |
| Protein - IL-8 | 0.156 |
| Comorbidity - CKD | 0.164 |
| MiRNA - hsa-miR-301a-3p | 0.167 |
| Protein - sVCAM-1 | 0.200 |
| MiRNA - hsa-miR-mir-30c-5p | 0.200 |
| CV Medication - Anticoagulant | 0.230 |
| Clinical - Male | 0.240 |
| History of dyslipidemia | 0.250 |
| Comorbidity - OSA | 0.250 |
| MiRNA - hsa-miR-146a-5p | 0.260 |
| MiRNA - hsa-miR-miR-30e-5p | 0.260 |
| Protein - IL-6 | 0.280 |
| MiRNA - hsa-miR-1 | 0.280 |
| Comorbidity - Malignancy | 0.280 |
| History of cardiac procedure/surgery | 0.300 |
| Comorbidity - GERD | 0.310 |
| History of any prior CV procedure | 0.310 |
| History of arrhythmia | 0.320 |
| Protein - sTREM-1 | 0.340 |
| Protein - E-Selectin | 0.350 |
| Comorbidity - Renal disease | 0.360 |
| MiRNA - hsa-miR-mir-339-3p | 0.380 |
| Comorbidity - Immunocompromised | 0.390 |
| Protein - sICAM-1 | 0.400 |
| CV medication - Number of medications | 0.430 |
| CV medication - ARB | 0.430 |
| MiRNA - hsa-miR-mir-4706 | 0.440 |
| Other CV condition - PFO | 0.440 |
| Comorbidity - Hypertension | 0.480 |
| History of myocardial infarction | 0.490 |
| Other CV condition - Aortic aneurysm | 0.490 |
| CV medication - Statin | 0.510 |
| Comorbidity - Diabetes | 0.520 |
| CV medication - ACE inhibitor | 0.520 |
| CV medication - CCB | 0.530 |
| Other CV condition - TIA | 0.550 |
| Other CV condition - Ischemic heart disease | 0.560 |
| Comorbidity - Vascular disease | 0.620 |
| Comorbidity - Pneumonia | 0.660 |
| Other CV condition - PAD | 0.680 |
| Comorbidity - COPD | 0.690 |
| MiRNA - hsa-miR-mir-26a-5p | 0.720 |
| Clinical - Patient ethnicity | 0.730 |
| CV medication - Beta blocker | 0.740 |
| MiRNA - hsa-miR-mir-30d-5p | 0.750 |
| CV medication - Diuretics | 0.760 |
| Comorbidity - CF | 0.800 |
| Other CV condition - Endocarditis | 0.800 |
| Other CV condition - Dilatated aortic root | 0.800 |
| Other CV condition - Becker’s muscle dystrophy | 0.800 |
| Comorbidity - Asthma | 0.850 |

| | |
|---|---|
| Comorbidity - Valvular heart disease | 0.930 |
| Other CV condition - VTE | 0.950 |
| Other CV condition - NSTEMI | 1.000 |

[*] Gout (n=3) was a small number of observations.

Bolded log-ranked P values are significant (P values <0.05). Abbreviations: ACE = Angiotensin-converting-enzyme inhibitors; Ang-2 = Angiopoetin-2; ARB = Angiotensin II receptor blockers; CCB = Calcium channel blocker; CKD = Chronic Kidney Disease; CF = Cystic Fibrosis; COPD = Chronic obstructive pulmonary disease; GERD = Gastroesophageal reflux disease; IL = Interleukin; miR = MicroRNA; MPO = Myeloperoxidase; NSTEMI = Non-ST-elevation myocardial infarction; OSA = Obstructive Sleep Apnea; PAD = Peripheral arterial disease; PFO = Patent foramen ovale; sICAM, Soluble intercellular adhesion molecule-1; sTREM-1, Soluble triggering receptor expressed on myeloid cells-1; sVCAM-1 = Soluble vascular cell adhesion molecule-1; TIA = Transient ischemic attack; VTE = Venous thromboembolism.

**Online Table V.** Intensive Care Unit-Level Clinical Characteristics and Clinical Outcomes

| Characteristics[*] | Severe (n=76) | Severe Negative (n=69) | P-value |
|---|---|---|---|
| Demographics | | | |
| Age, median (IQR) – yr. | 61 (52-71) | 61 (51-72) | 0.5137 |
| Distribution – no. (%) | | | |
| 18-40 yr. | 2 (2.6) | 11 (15.9) | **0.0071** |
| 41-64 yr. | 45 (59.2) | 26 (37.7) | **0.0126** |
| ≥65 yr. | 29 (38.2) | 32 (46.4) | 0.3999 |
| Male Sex, no./total no. (%) | 52/76 (68.4) | 46/69 (66.7) | 0.8602 |
| BMI, median (IQR)[†] | 28.1 (24.2-31.8) | 25.7 (21.8-32.5) | 0.3641 |
| Obesity[‡] – no. (%) | 38/61 (62.3) | 35/69 (50.7) | 0.2168 |
| Length of hospital stay, median (IQR) – days† | 13 (7-35) | 8 (3-15) | **<0.0001** |
| Max temperature, median (IQR), °C | 36.8 (36.3-37.8) | 36.4 (35.8-37.1) | 0.0797 |
| Mix temperature, median (IQR), °C | 36.7 (36.3-37.3) | 36.3 (35.6-37.1) | 0.0815 |
| Illness Severity | | | |
| APACHE | 21 (16-27) | 20 (16-29) | 0.8521 |
| SOFA score | | | |
| Median, (IQR) | 9 (4-10) | 7 (3-10) | 0.8112 |
| ≥2 – no. (%) | 12/38 (31.6) | 18/69 (26.1) | 0.3674 |
| ≥6 – no. (%) | 24/38 (63.2) | 44/69 (63.8) | >0.9999 |
| Respiratory metrics | | | |
| Proned – no. (%) | 8/38 (21.1) | 3/69 (4.3) | **0.0158** |
| Intubation – no. (%) | 53/76 (69.7) | 47/69 (68.1) | 0.8565 |
| NIV – mean, days† | 3.12 | 1.5 | 0.2177 |
| MIV – mean, days† | 10.84 | 7.61 | 0.2215 |
| ECMO – no. (%) | 21/76 (27.6) | 0 (0.0) | **<0.0001** |
| $FiO_2$, median % (IQR) | 0.53 (0.50-0.63) | 0.40 (0.30-0.50) | **<0.0001** |
| PF ratio, median (IQR), mm Hg/% | 130 (103-177) | 188 (146-289) | **0.0003** |
| PF ratio <300 mm Hg/% | 26/26 (100.0) | 38/49 (77.6) | **0.0126** |
| RR, no. (%), ≥22 breaths/min | 22/38 (57.9) | 33/65 (50.8) | 0.5425 |
| Cardiovascular metrics | | | |
| Heart rate, median (IQR) | 80 (67-101) | 82 (67-97) | 0.6911 |
| Blood Pressure, median (IQR), mmHg | | | |
| Max Systolic | 165 (141-179) | 140 (125-165) | **0.0242** |
| Max Diastolic | 75 (65-84) | 69 (63-80) | 0.1707 |
| Therapies – no. (%) | | | |
| Intravenous antibiotics | 11/38 (28.9) | 20/69 (29.0) | >0.9999 |
| Systemic glucocorticoids | 13/38 (34.2) | 37/67 (55.2) | 0.2016 |
| Remdesivir | 1/38 (2.6) | 0/69 (0.0) | 0.3679 |
| Fludrocortisone | 0/38 (0.0) | 5/69 (7.2) | 0.1583 |
| Outcomes – no. (%) | | | |
| Any secondary CV event | 13/76 (17.1) | 1/69 (1.4) | **0.0013** |
| ARDS during hospitalization | 43/76 (56.6) | 25/69 (36.2) | **0.0195** |
| Arrythmia during hospitalization | 8/76 (10.5) | 0/69 (0.0) | **0.0068** |

[*] Summary statistics, where n-value is not provided, are based on the full population indicated in the column heading.
† Body mass index is the weight in kilograms divided by the square of the height in meters.
‡ Obesity is classified according to World Health Organization guidelines (i.e., >25).

Bolded log-ranked P values are significant (P values <0.05). Abbreviations: APACHE = Acute physiologic assessment and chronic health evaluation; ARDS = Acute respiratory distress syndrome; BMI = Body mass index; CV = Cardiovascular; ECMO = Extracorporeal membrane oxygenation; $FiO_2$ = Fraction of inspired oxygen; IQR = Interquartile range; MIV = Mechanical ventilation; NIV = Non-invasive ventilation; PF = Ratio of arterial oxygen partial pressure to fractional inspired oxygen; RR = Respiratory rate; S.D. = Standard deviation; SOFA = Sequential Organ Failure Assessment. Percentages may not add up to 100% due to rounding.

**Online Table VI.** Association of Baseline Clinical Characteristics to Mortality Amongst All Severe COVID-19 Patients.

| Variables (Baseline) | Unadjusted Log-Rank P Value |
|---|---|
| Comorbidity – Gout | **<0.001** |
| MiRNA – -miR-30b-5p | **0.005** |
| MiRNA - hsa-miR-199a-3p | **0.006** |
| MiRNA - hsa-miR-181a-5p | **0.010** |
| Protein - VCAM-1 | **0.012** |
| MiRNA - hsa-miR-339-3p | **0.017** |
| MiRNA - hsa-miR-mir-30e-5p | **0.017** |
| Protein - Ang-2 | **0.020** |
| MiRNA - hsa-miR-146a-5p | **0.030** |
| MiRNA - hsa-miR-30c-5p | **0.035** |
| Comorbidity - CKD | **0.038** |
| History of coronary artery disease at baseline | **0.041** |
| MiRNA - hsa-miR-6080 | **0.042** |
| MiRNA - hsa-miR-6750-5p | 0.051 |
| MiRNA - hsa-miR-4793-5p | 0.060 |
| Protein - ICAM-1 | 0.096 |
| MiRNA - hsa-miR-301a-3p | 0.103 |
| CV medication - Anticoagulant | 0.103 |
| Clinical - Age of patient at hospital admission | 0.122 |
| Comorbidit–y - OSA | 0.126 |
| Clinical - Male | 0.130 |
| MiRNA - hsa-miR-4706 | 0.131 |
| Comorbidity - Stroke | 0.184 |
| Protein - IL-8 | 0.220 |
| Comorbidity - Heart failure | 0.220 |
| MiRNA - hsa-miR-26a-5p | 0.230 |
| Protein - sTREM-1 | 0.230 |
| History of arrhythmia | 0.240 |
| Comorbidity - Obesity | 0.260 |
| Comorbidity - COPD | 0.260 |
| MiRNA - hsa-miR-1 | 0.270 |
| History of any prior CV procedure | 0.310 |
| Protein - IL-6 | 0.320 |
| Protein - MPO | 0.320 |
| Comorbidity - GERD | 0.320 |
| Comorbidity - Dyslipidemia | 0.380 |
| Other CV condition - TIA | 0.380 |
| Comorbidity - Peripheral vascular disease | 0.410 |
| Comorbidity - Immunocompromised | 0.410 |
| Comorbidity - Valvular heart disease | 0.420 |
| Other CV condition - PFO | 0.440 |
| MiRNA - hsa-miR-30d-5p | 0.460 |
| CV medication - Statin | 0.480 |
| CV medication - Diuretics | 0.540 |
| CV medication - Number of medications | 0.550 |
| CV medication - ARB | 0.550 |
| Comorbidity - Renal disease | 0.560 |
| Other CV condition – Ischemic heart disease | 0.560 |
| Comorbidity - Hypertension | 0.590 |
| CV medication - CCB | 0.590 |
| History of myocardial infarction | 0.600 |
| Comorbidity - Malignancy | 0.600 |
| Other CV condition - Aortic aneurysm | 0.620 |
| CV medication - ACE inhibitor | 0.650 |
| Comorbidity - Pneumonia | 0.660 |
| Clinical - Patient ethnicity | 0.730 |
| Comorbidity - Diabetes | 0.780 |
| Protein - sE-Selectin | 0.810 |
| Comorbidity - Asthma | 0.810 |
| Other CV condition - VTE | 0.840 |
| CV medication - Beta blocker | 0.880 |

* Gout (n=3) was a small number of observations.

Bolded log-ranked P values are significant (P values <0.05). Abbreviations: ACE = Angiotensin-converting-enzyme inhibitors; Ang-2 = Angiopoetin-2; ARB = Angiotensin II receptor blockers; CCB = Calcium channel blocker; CKD = Chronic Kidney Disease; CF = Cystic Fibrosis; COPD = Chronic obstructive pulmonary disease; GERD = Gastroesophageal reflux disease; IL = Interleukin; miR = MicroRNA; MPO = Myeloperoxidase; OSA = Obstructive Sleep Apnea; PFO = Patent foramen ovale; sICAM, Soluble intercellular adhesion molecule-1; sTREM-1, Soluble triggering receptor expressed on myeloid cells-1; sVCAM-1 = Soluble vascular cell adhesion molecule-1; TIA = Transient ischemic attack; VTE = Venous thromboembolism.

#### SUPPLEMENTAL FIGURES

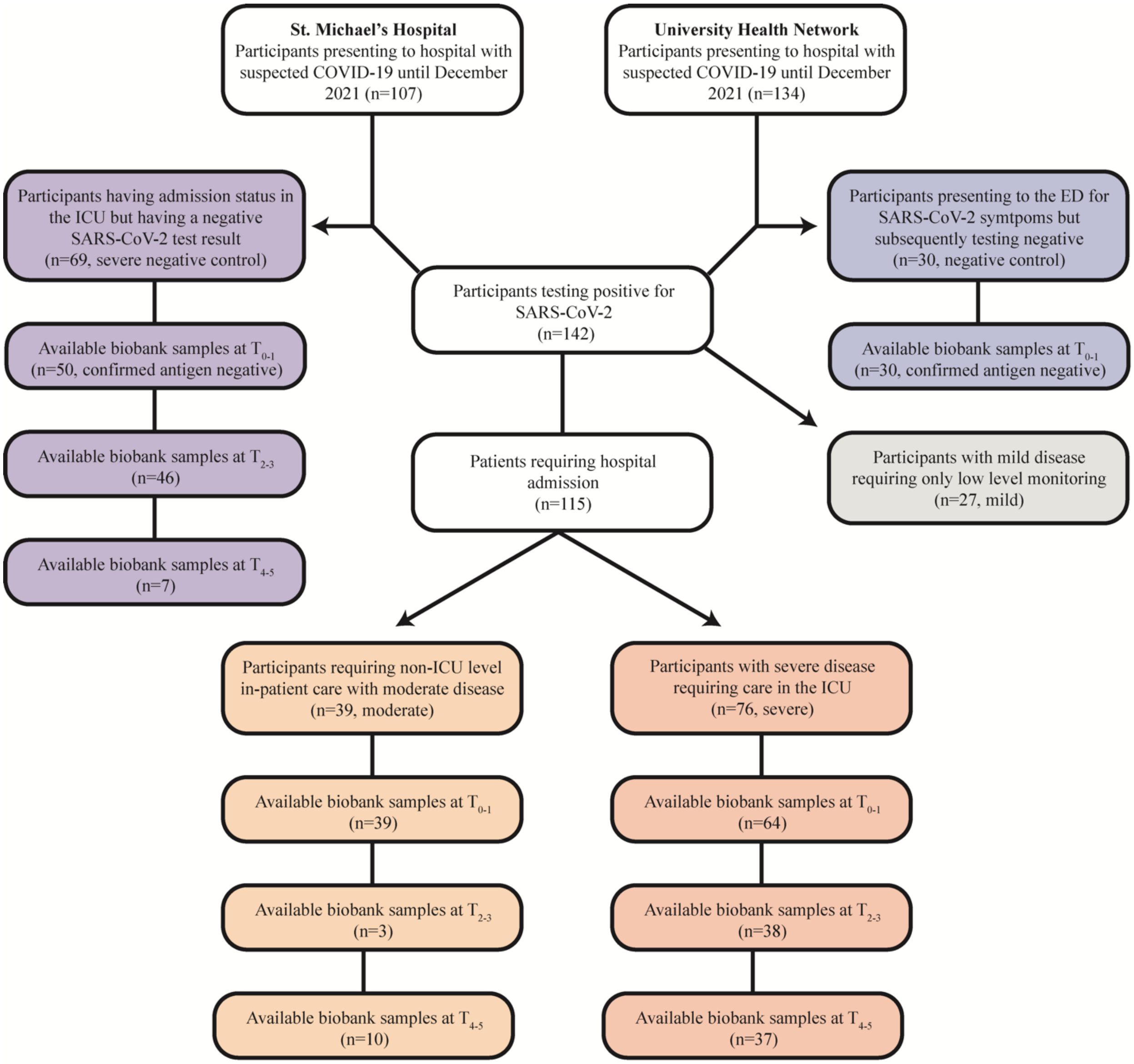

**Online Figure I; Related to Methods and Table 1:** Flow diagram of patients enrolled between the COLOBILI Study (St. Michael's Hospital) and the COVID Study (University Health Network). Abbreviations: COVID-19 = Coronavirus disease 2019; ED = Emergency department; ICU = Intensive care unit.

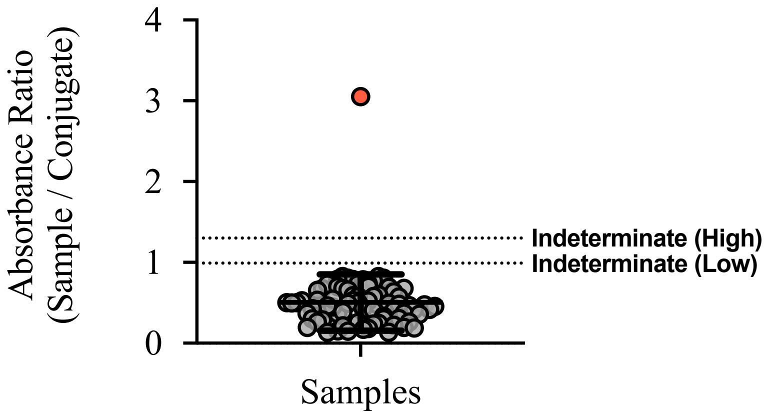

**Online Figure II; Related to Methods and Table 1:** Spike (trimer) antigen serology testing from patients having a negative SARS-CoV-2 polymerase chain reaction result. The data were analyzed using built-in low and high thresholds, whereby the area between the indeterminate-low and indeterminate-high constitutes an ambiguous result; n=80. Positive control is indicated on the graph (red circle). The graph depicts averaged values of independent technical duplicates data with center bars representing the mean and error bars representing standard deviation (±S.D.).

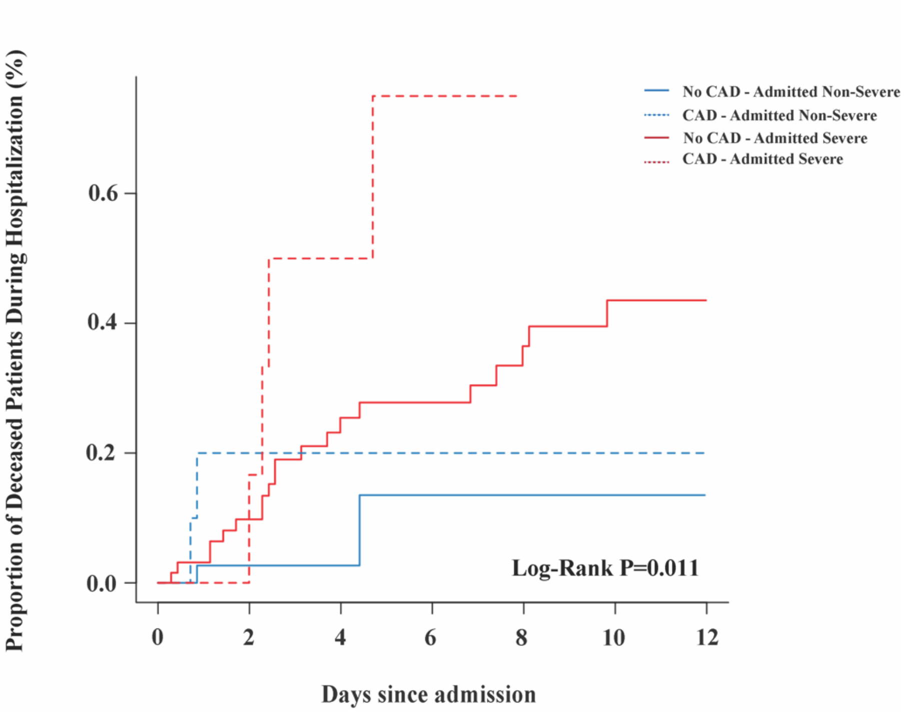

**Online Figure III; Related to Figure 1:** The association of coronary artery disease with mortality characterized in terms of proportion of deceased patients stratified by status. The data were analyzed using log-rank testing. Abbreviations: CAD = Coronary artery disease.

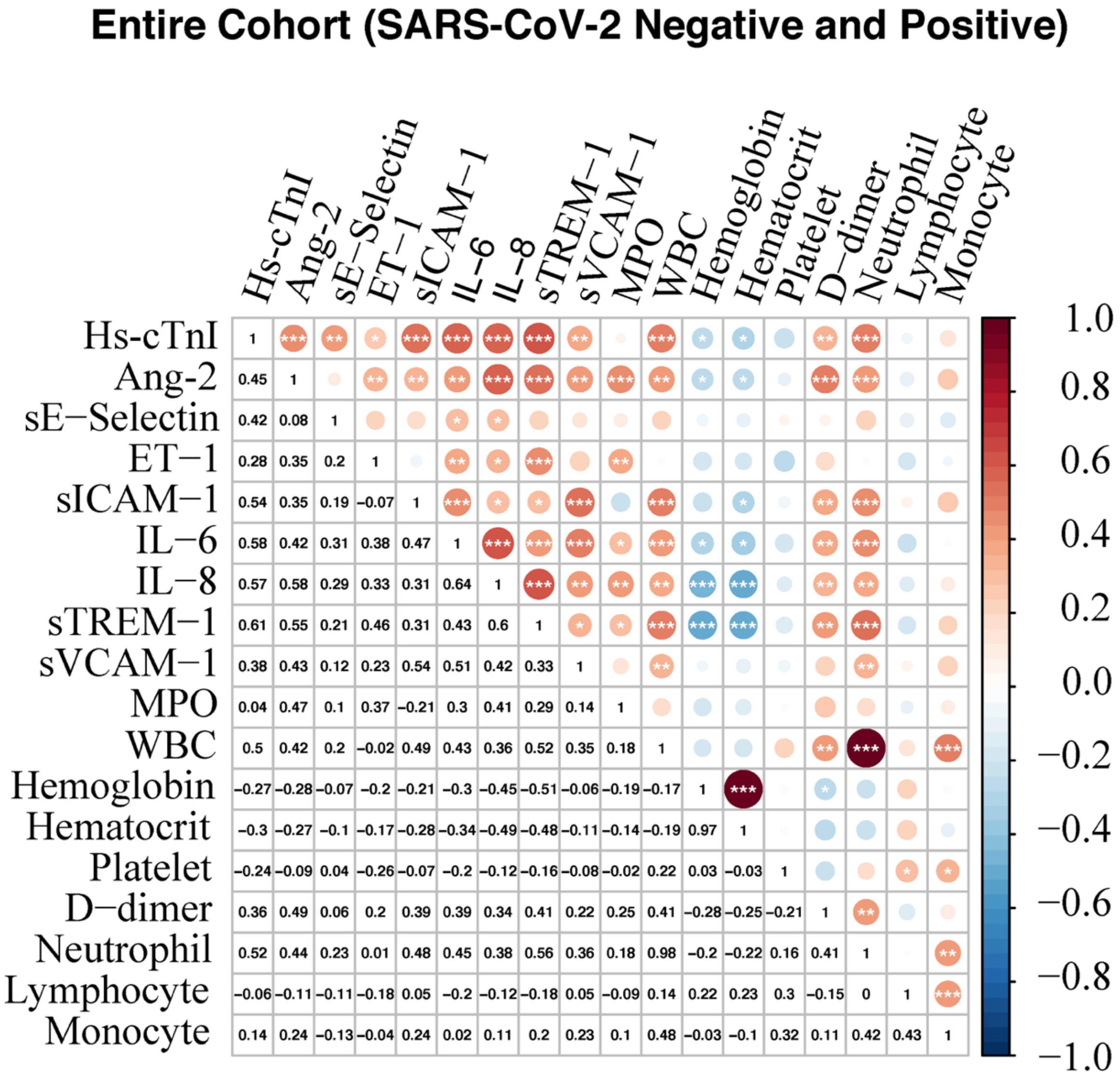

**Online Figure IV; Related to Figure 2:** Spearman correlations between $t_{0-1}$ concentrations of biomarkers amongst the entire cohort (SARS-CoV-2 negative and positive populations). Abbreviations: Ang-2 = Angiopoietin-2; ET-1 = Endothelin-1; Hs-CTnI = High-sensitivity cardiac troponin I; IL = Interleukin; MPO = Myeloperoxidase; sICAM = Soluble intercellular adhesion molecule-1; sTREM-1 = Soluble triggering receptor expressed on myeloid cells-1; sVCAM-1 = Soluble vascular cell adhesion molecule-1; WBC = White blood cells.

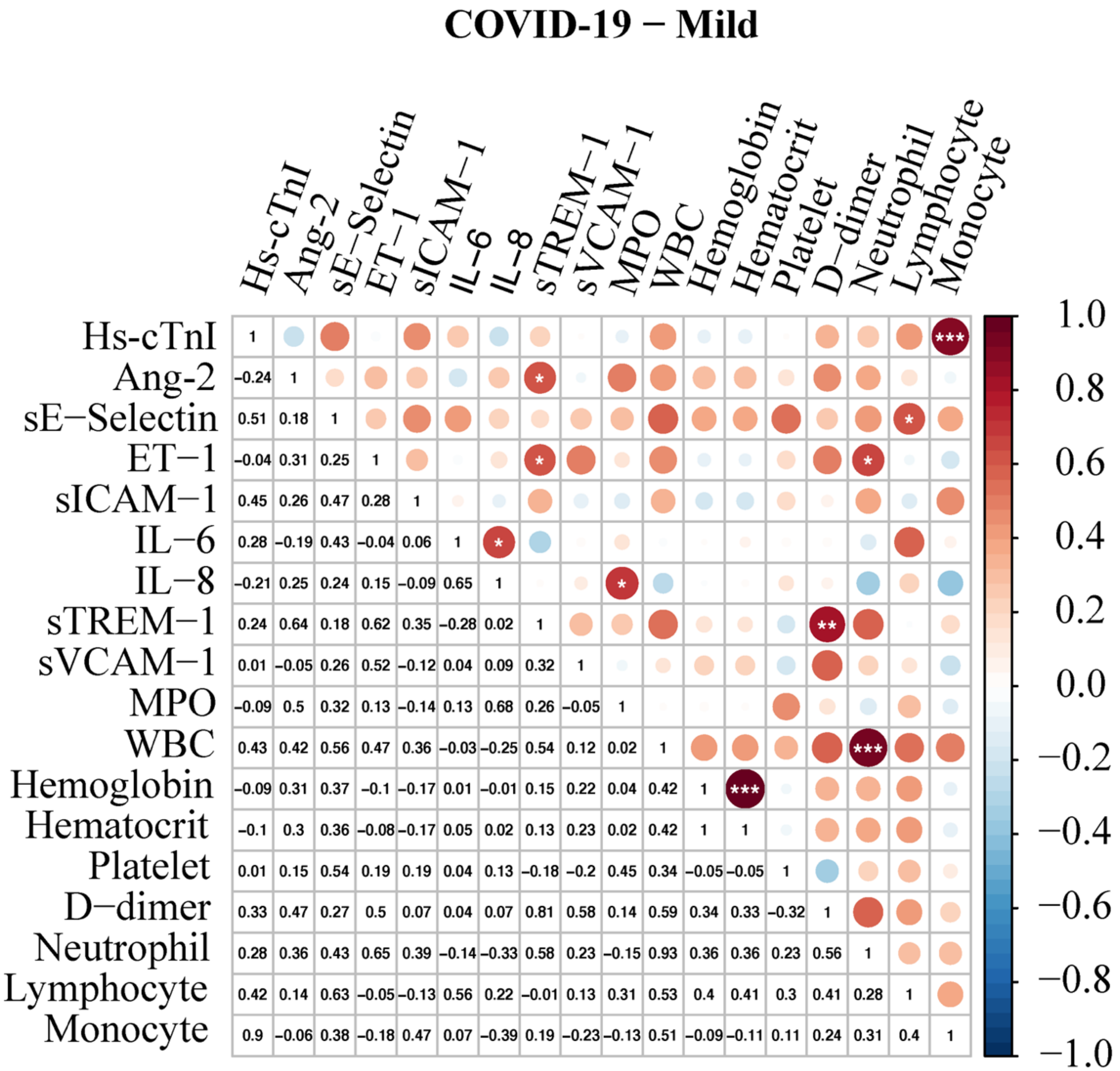

**Online Figure V; Related to Figure 2:** Spearman correlations between $t_{0\text{-}1}$ concentrations of biomarkers within the mild COVID-19 subgroup. Abbreviations: Ang-2 = Angiopoietin-2; ET-1 = Endothelin-1; Hs-cTnI = High-sensitivity cardiac troponin I; IL = Interleukin; MPO = Myeloperoxidase; sICAM = Soluble intercellular adhesion molecule-1; sTREM-1 = Soluble triggering receptor expressed on myeloid cells-1; sVCAM-1 = Soluble vascular cell adhesion molecule-1; WBC = White blood cells.

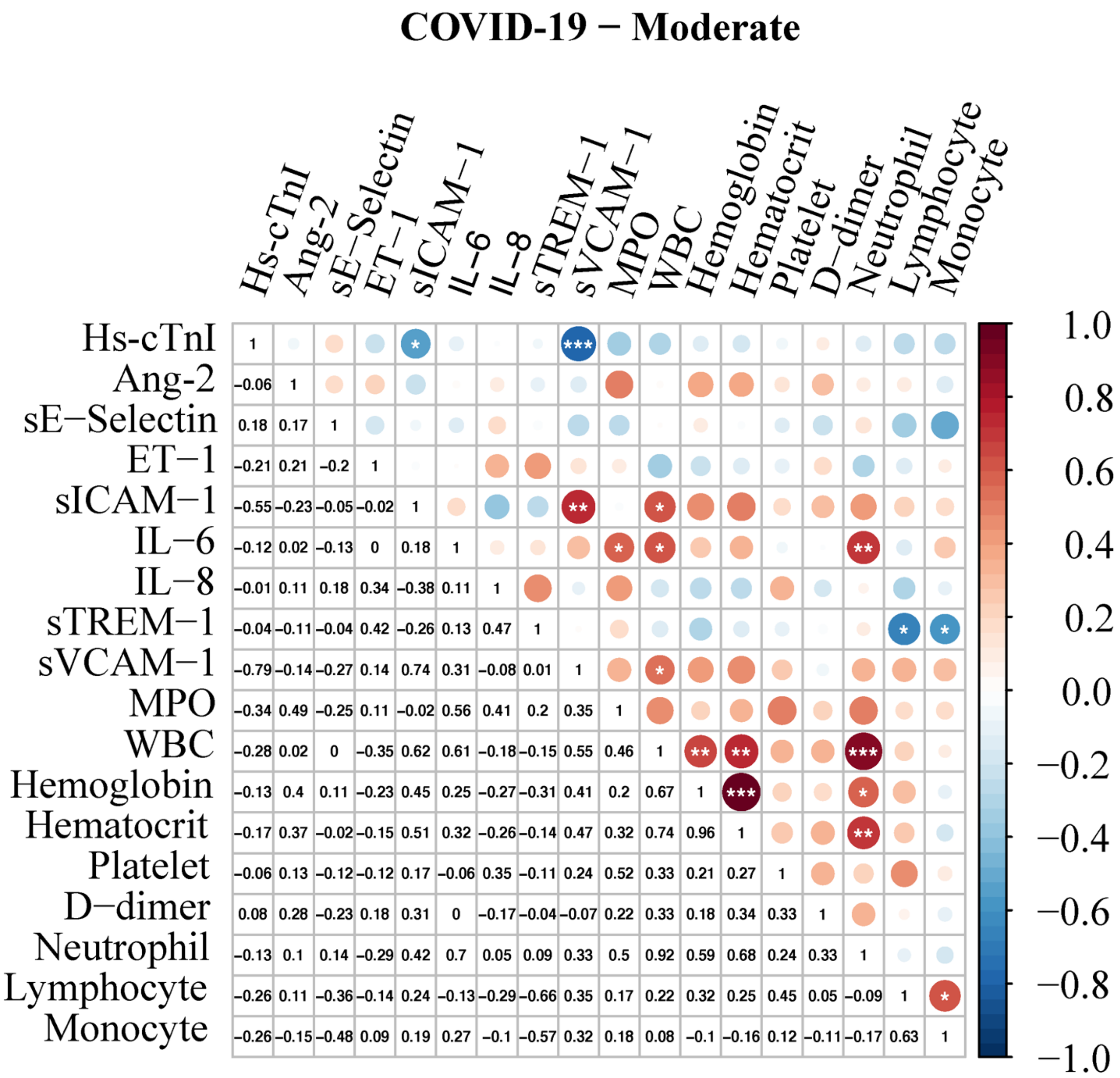

**Online Figure VI; Related to Figure 2:** Spearman correlations between $t_{0-1}$ concentrations of biomarkers within the severe COVID-19 subgroup. Abbreviations: Ang-2 = Angiopoietin-2; ET-1 = Endothelin-1; Hs-cTnI = High-sensitivity cardiac troponin I; IL = Interleukin; MPO = Myeloperoxidase; sICAM = Soluble intercellular adhesion molecule-1; sTREM-1 = Soluble triggering receptor expressed on myeloid cells-1; sVCAM-1 = Soluble vascular cell adhesion molecule-1; WBC = White blood cells.

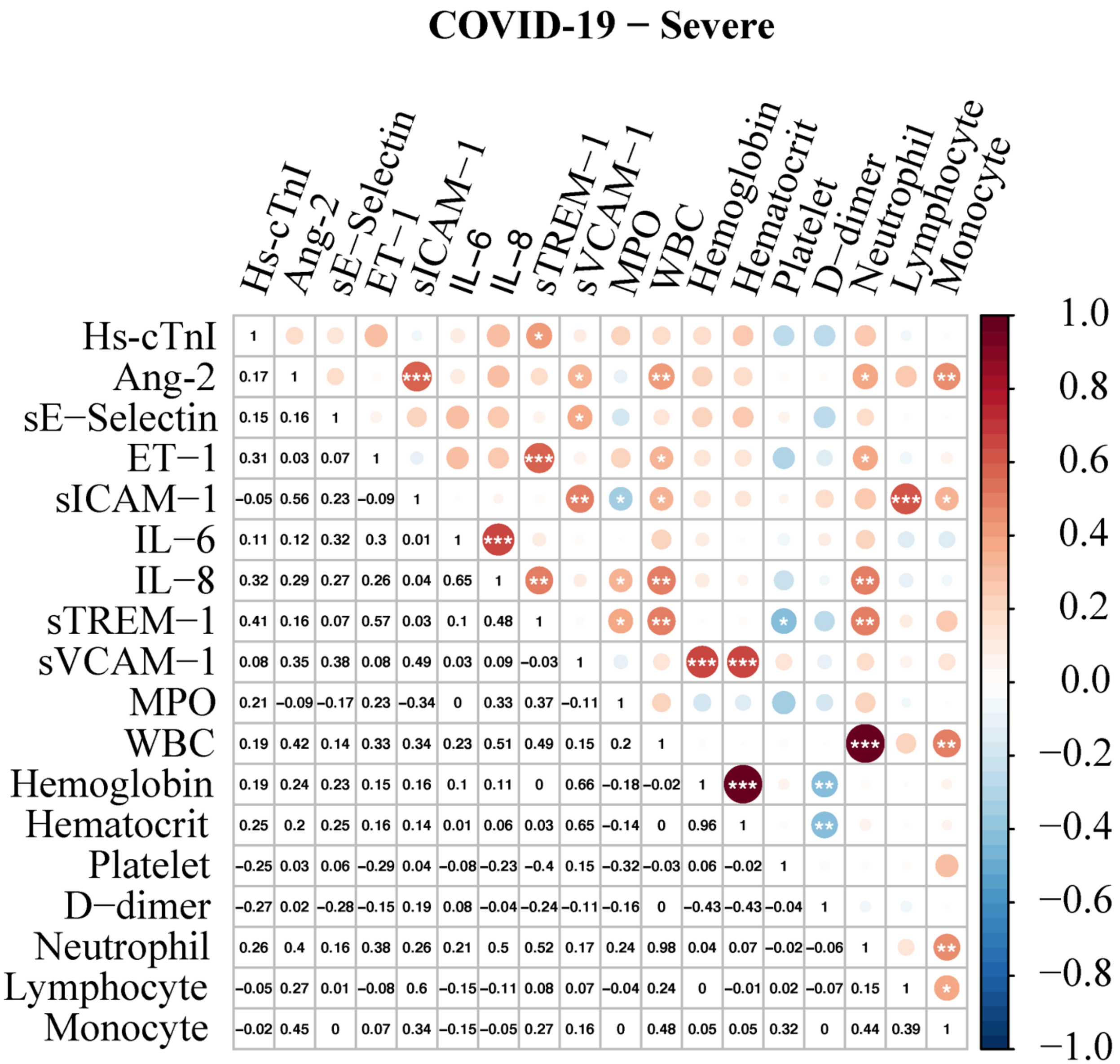

**Online Figure VII; Related to Figure 2:** Spearman correlations between $t_{0\text{-}1}$ concentrations of biomarkers within the COVID-19 subgroup. Abbreviations: Ang-2 = Angiopoietin-2; ET-1 = Endothelin-1; Hs-cTnI = High-sensitivity cardiac troponin I; IL = Interleukin; MPO = Myeloperoxidase; sICAM = Soluble intercellular adhesion molecule-1; sTREM-1 = Soluble triggering receptor expressed on myeloid cells-1; sVCAM-1 = Soluble vascular cell adhesion molecule-1; WBC = White blood cells.

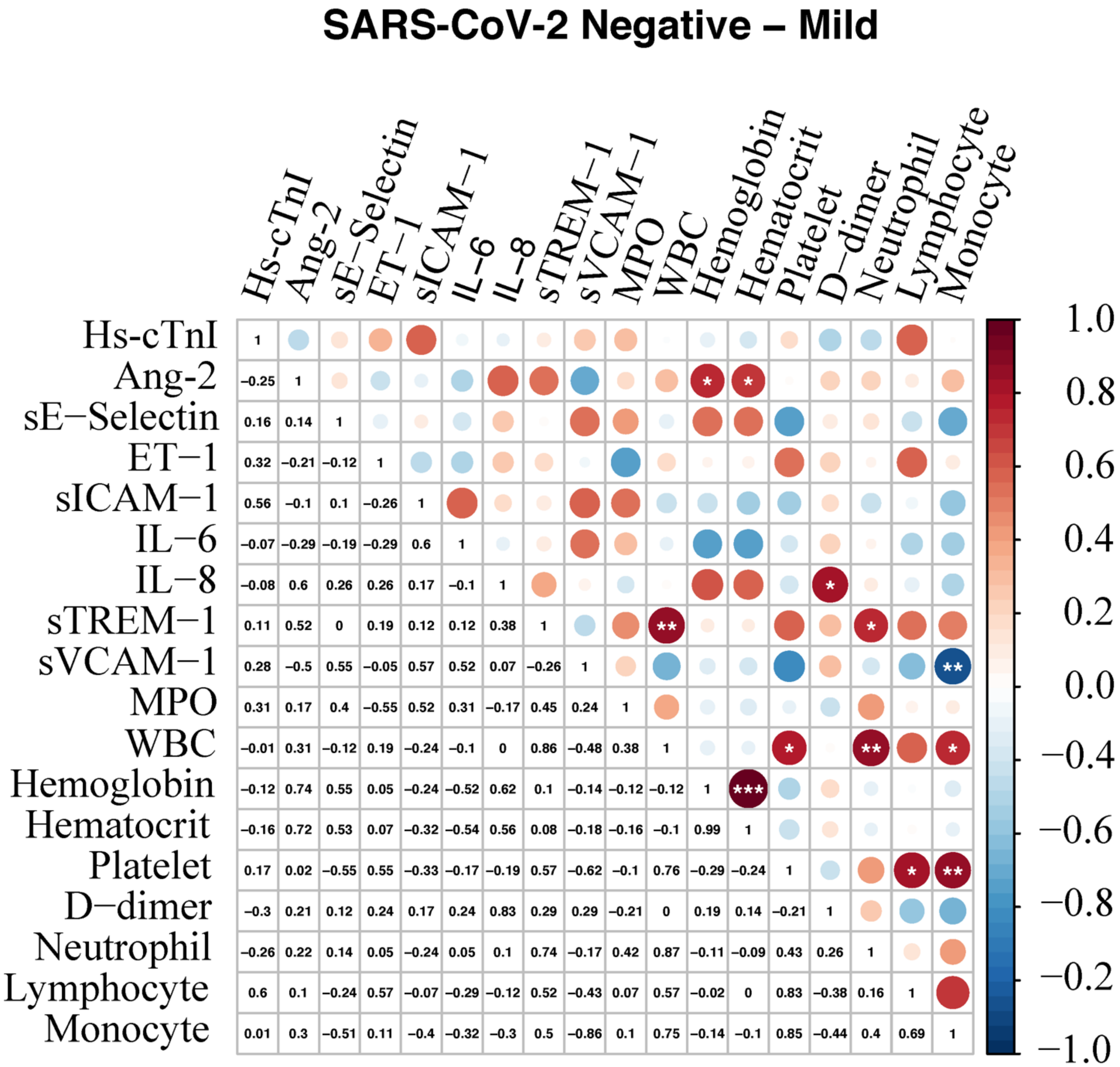

**Online Figure VIII; Related to Figure 2:** Spearman correlations between $t_{0\text{-}1}$ concentrations of biomarkers within the mild SARS-CoV-2 negative subgroup. Abbreviations: Ang-2 = Angiopoietin-2; ET-1 = Endothelin-1; Hs-cTnI = High-sensitivity cardiac troponin I; IL = Interleukin; MPO = Myeloperoxidase; sICAM = Soluble intercellular adhesion molecule-1; sTREM-1 = Soluble triggering receptor expressed on myeloid cells-1; sVCAM-1 = Soluble vascular cell adhesion molecule-1; WBC = White blood cells.

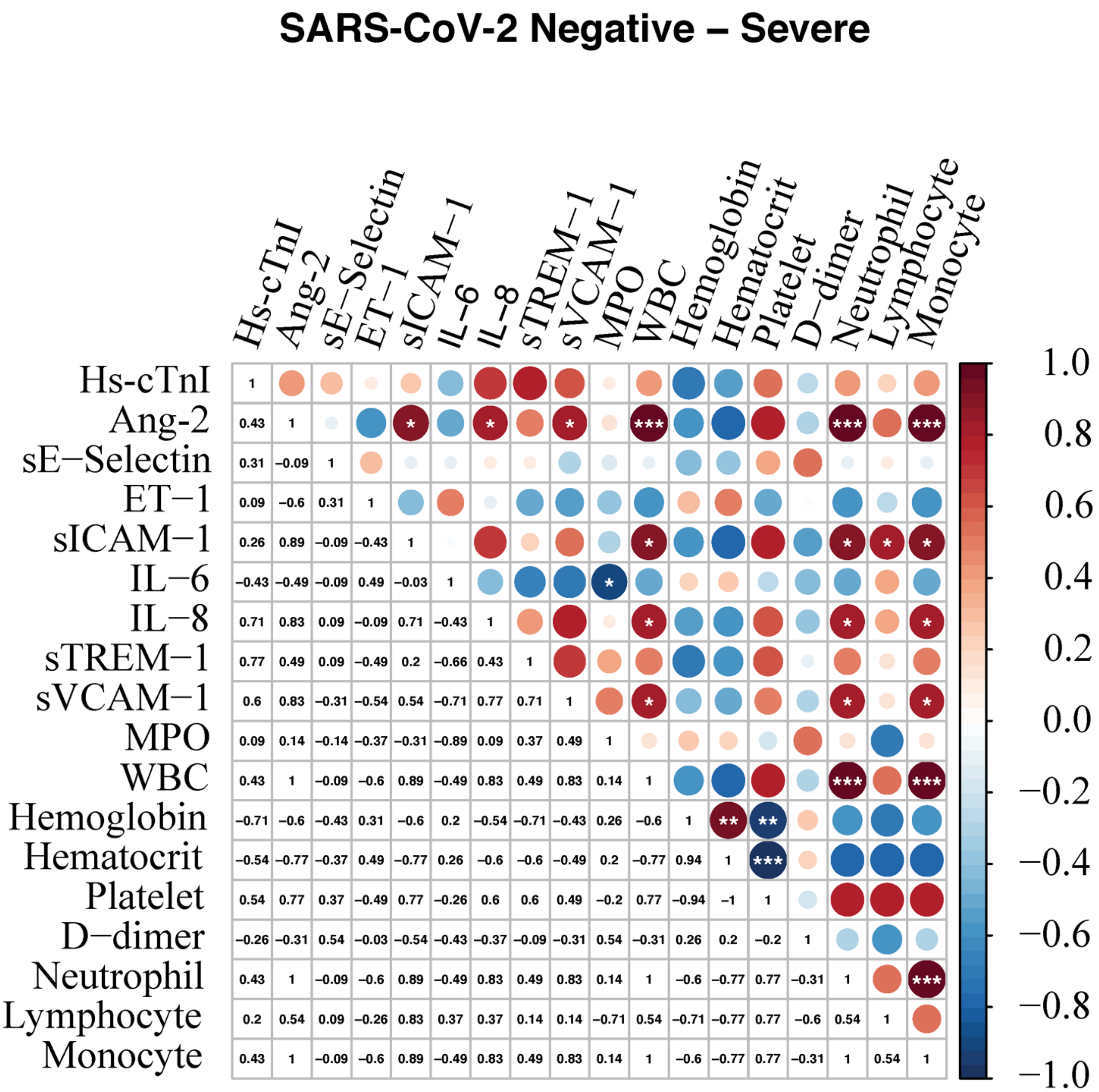

**Online Figure IX; Related to Figure 2:** Spearman correlations between $t_{0-1}$ concentrations of biomarkers within the severe SARS-CoV-2 negative subgroup. Abbreviations: Ang-2 = Angiopoietin-2; ET-1 = Endothelin-1; Hs-cTnI = High-sensitivity cardiac troponin I; IL = Interleukin; MPO = Myeloperoxidase; sICAM = Soluble intercellular adhesion molecule-1; sTREM-1 = Soluble triggering receptor expressed on myeloid cells-1; sVCAM-1 = Soluble vascular cell adhesion molecule-1; WBC = White blood cells.

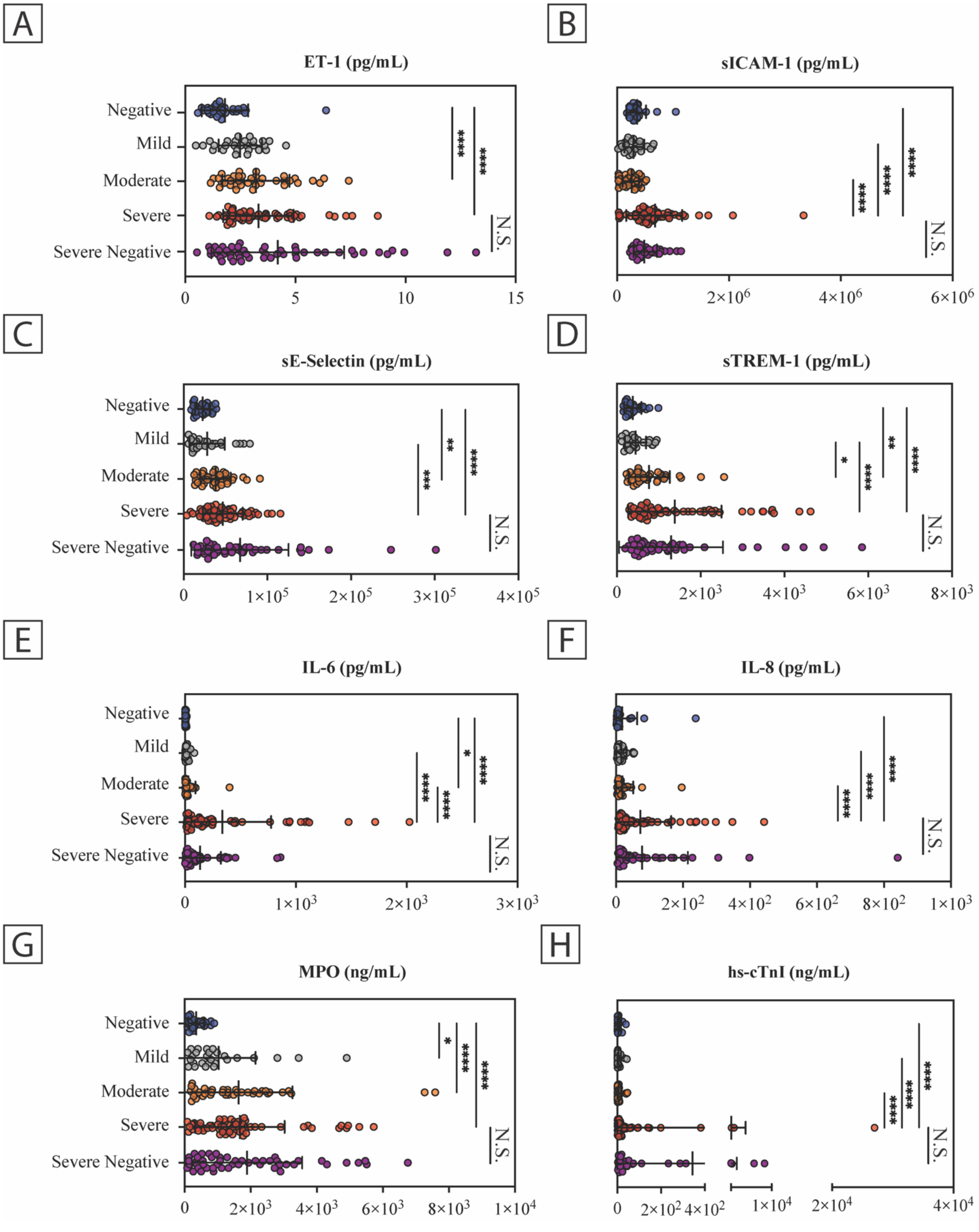

**Online Figure X; Related to Figure 2.** Plasma Concentration of Endothelial Dysfunction and Inflammatory Markers at $t_{0\text{-}1}$. (a) ET-1, (b) sICAM-1, (c) sE-Selectin, (d) sTREM-1, (e) IL-6, (f) IL-8, (g) MPO, and (h) hs-cTnI stratified among disease severity. Data shown are for all patients with an available $t_{0-1}$ sample (n=210), with values representing the mean and error bars are (±S.D.). P values for multiple group comparisons were determined by Kruskal-Wallis test with Dunn's multiple comparisons test. Severe negative comparisons are only shown in reference to the concordant severe group; testing was conducted with all groups. The graph depicts averaged values of independent technical triplicates data points with center bars representing the mean and error bars representing standard deviation (±S.D.). Abbreviations: ET-1 = Endothelin-1; hs-cTnI = High-sensitivity cardiac troponin I; sICAM = Soluble intercellular adhesion molecule-1; IL = Interleukin; MPO = Myeloperoxidase; sTREM-1 = Soluble triggering receptor expressed on myeloid cells 1; N.S. = non-significant.

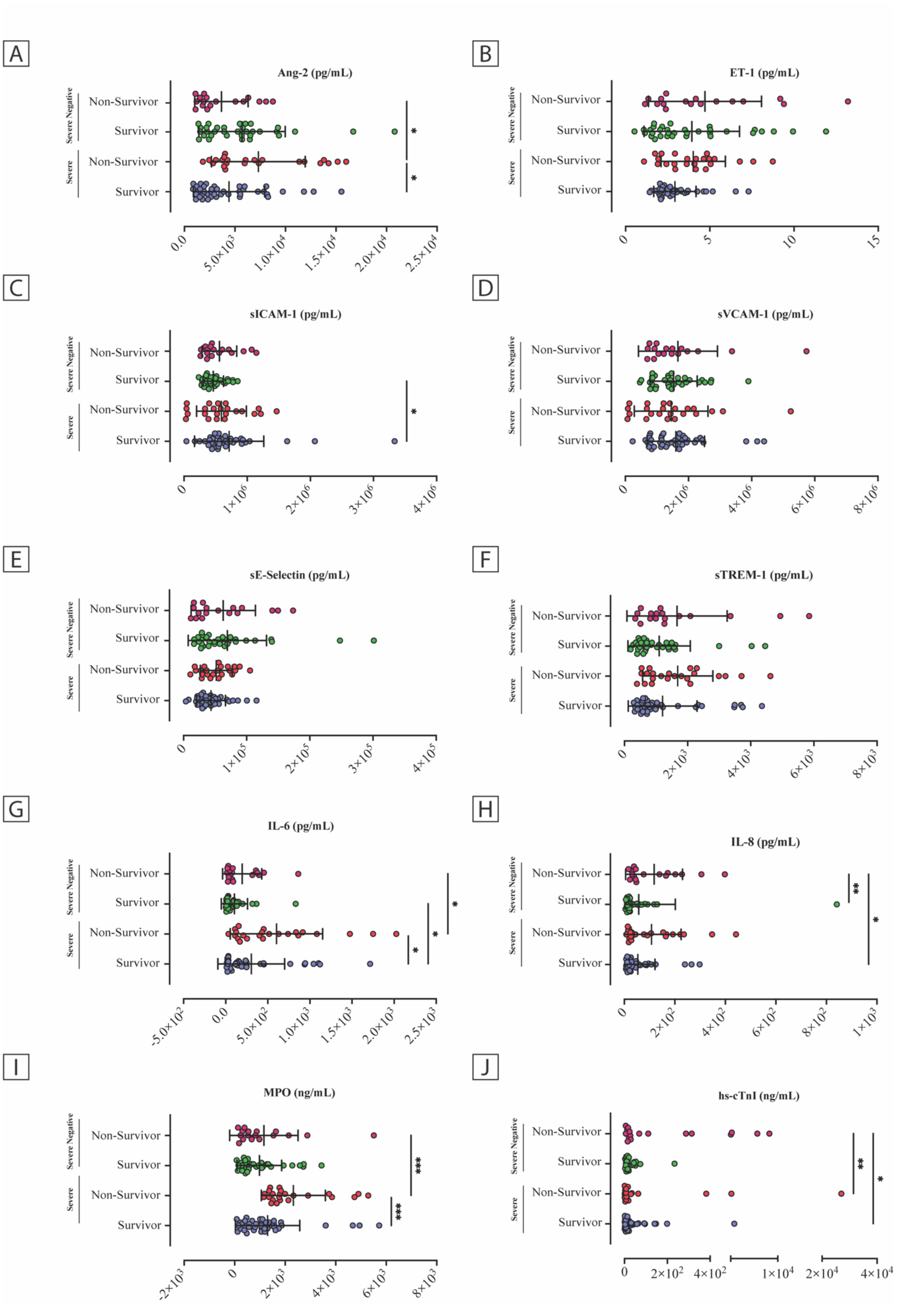
A
Ang-2 (pg/mL)
B
ET-1 (pg/mL)
C
sICAM-1 (pg/mL)
D
sVCAM-1 (pg/mL)
E
sE-Selectin (pg/mL)
F
sTREM-1 (pg/mL)
G
IL-6 (pg/mL)
H
IL-8 (pg/mL)
I
MPO (ng/mL)
J
hs-cTnI (ng/mL)
Severe Negative
Severe
Non-Survivor
Survivor

**Online Figure XI; Related to Figure 2:** Plasma Concentration of Endothelial Dysfunction and Inflammatory Markers at $t_{0-1}$ and ability to discriminate survival in ICU patients. Severe COVID-19 patients and severe negative patients (i.e., SARS-CoV-2 negative) (a) Levels of Angiopoietin-2, (b) Endothelin-1, (c) sICAM-1, (d) sVCAM-1, (e) sE-Selectin, (f) sTREM-1, (g) IL-6, (h) IL-8, (i) MPO, and (j) hs-cTnI stratified among disease severity. Data shown are for all severe patients with an available $t_{0-1}$ sample (n=114), with the center bars representing the mean and error bars representing standard deviation (±S.D.). P values for multiple group comparisons were determined by Kruskal-Wallis test with Dunn's multiple comparisons test. Abbreviations: Ang-2 = Angiopoietin-2; sICAM = Soluble intercellular adhesion molecule 1; IL = Interleukin; MPO = Myeloperoxidase; sTREM-1 = Soluble triggering receptor expressed on myeloid cells 1; sVCAM-1 = Soluble vascular cell adhesion molecule 1.

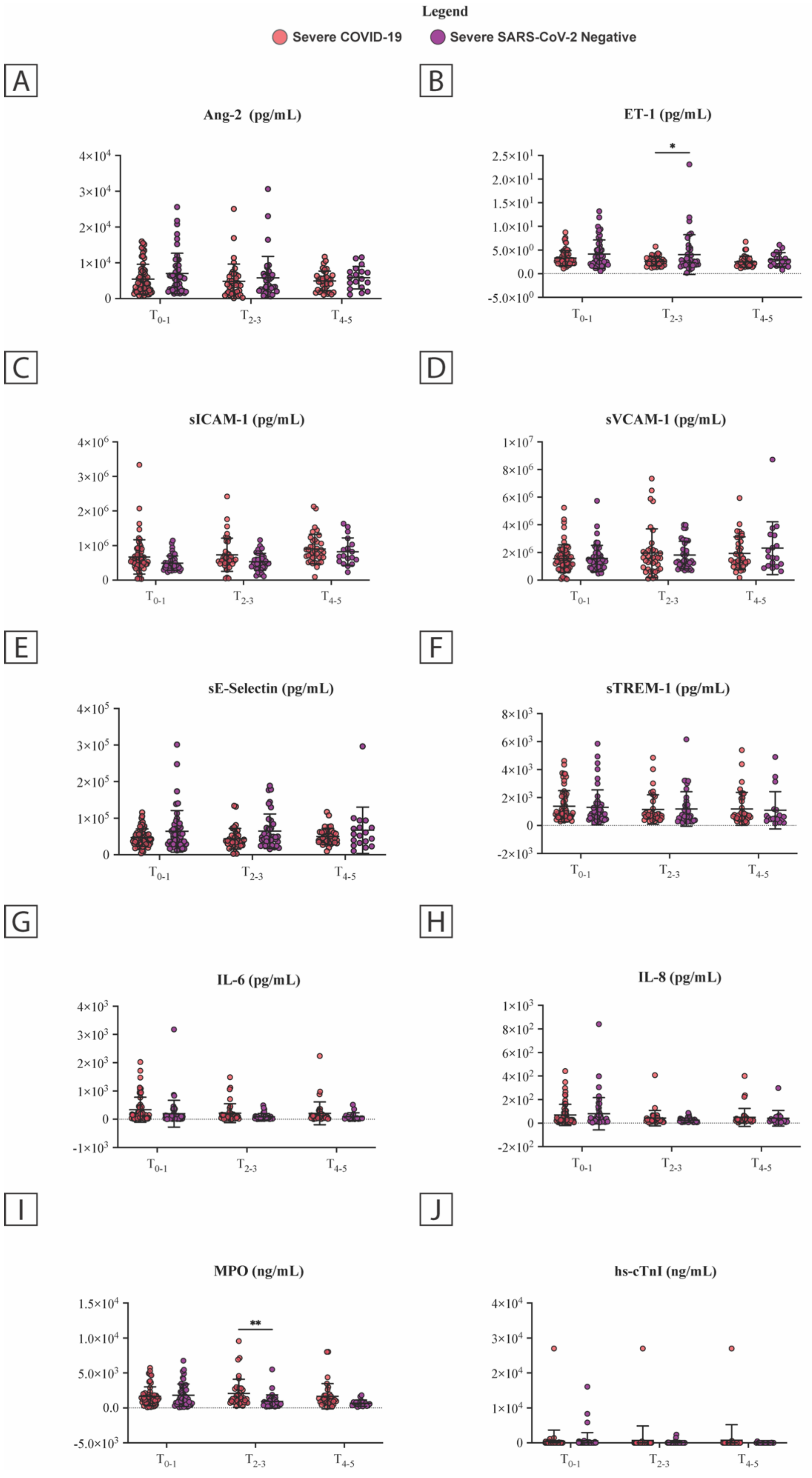
Legend
Severe COVID-19
Severe SARS-CoV-2 Negative
A
Ang-2 (pg/mL)
B
ET-1 (pg/mL)
C
sICAM-1 (pg/mL)
D
sVCAM-1 (pg/mL)
E
sE-Selectin (pg/mL)
F
sTREM-1 (pg/mL)
G
IL-6 (pg/mL)
H
IL-8 (pg/mL)
I
MPO (ng/mL)
J
hs-cTnI (ng/mL)
$T_{0-1}$
$T_{2-3}$
$T_{4-5}$

**Online Figure XII; Related to Figure 2:** Plasma Concentration of Endothelial Dysfunction and Immunological Markers at $t_{0-1}$ in ICU patients. (a) Levels of Angiopoietin-2, (b) Endothelin-1, (c) sICAM-1, (d) sVCAM-1, (e) sE-Selectin, (f) sTREM-1, (g) IL-6, (h) IL-8, (i) MPO, and (j) hs-cTnI stratified among disease severity. Data shown are for all patients with an available $t_{0-1}$ (n=114), $t_{2-3}$ (n=84), and $t_{4-5}$ (n=44), with center bars representing the mean and error bars representing standard deviation (±S.D.). P values for multiple group comparisons were determined by 2-way ANOVA with Sidak's multiple comparisons test. Abbreviations: sICAM = Soluble intercellular adhesion molecule 1; IL = Interleukin; MPO = Myeloperoxidase; sTREM-1 = Soluble triggering receptor expressed on myeloid cells 1; sVCAM-1 = Soluble vascular cell adhesion molecule 1.

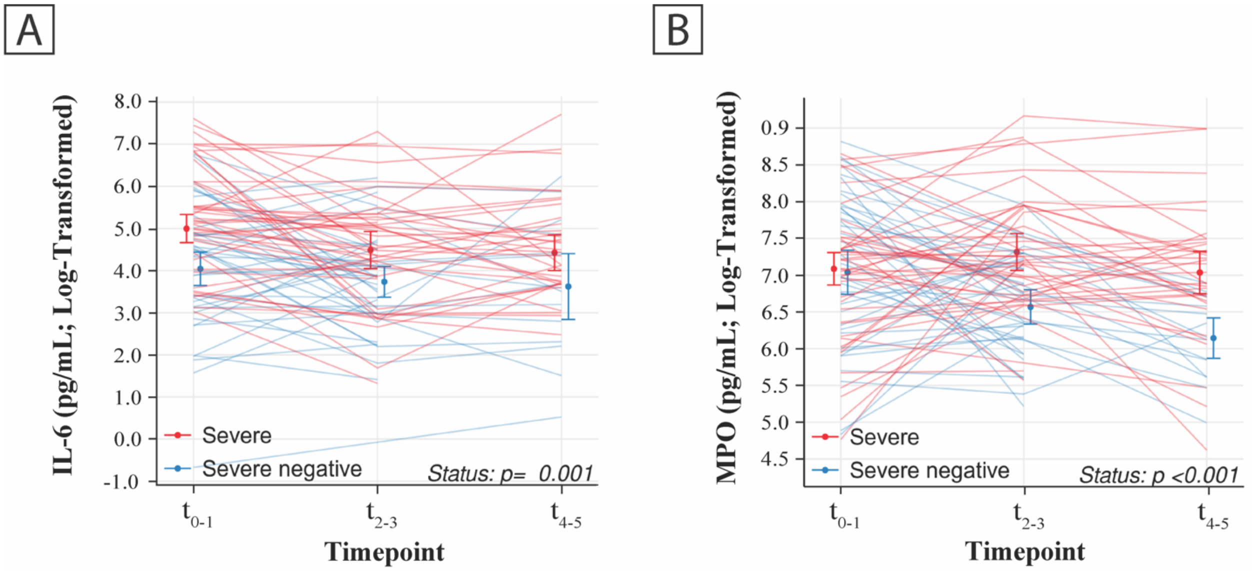

**Online Figure XIII; Related to Figure 2:** Plasma Concentration of (a) IL-6 and (b) MPO, longitudinally between severe COVID-19 patients and severe SARS-CoV-2 negative patients. The error bars represent the mean and its 95% confidence intervals estimated using generalized estimating equation with an independent working correlation matrix. The standard errors were estimated using robust sandwich estimator. Abbreviations: IL = Interleukin; MPO = Myeloperoxidase.

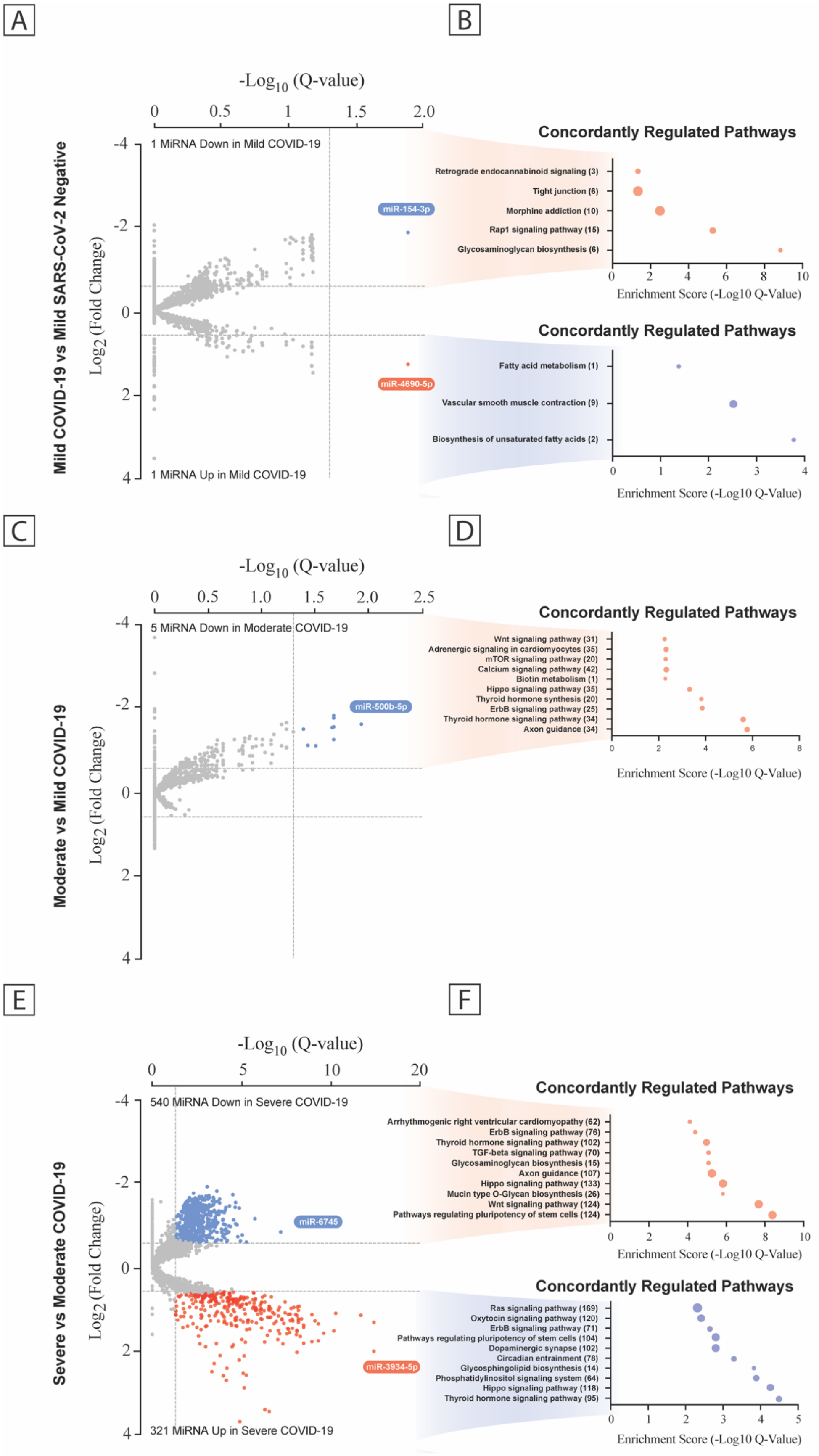

A
B
-Log10 (Q-value)
Mild COVID-19 vs Mild SARS-CoV-2 Negative
Log2 (Fold Change)
1 MiRNA Down in Mild COVID-19
1 MiRNA Up in Mild COVID-19
miR-154-3p
miR-4690-5p
Concordantly Regulated Pathways
Retrograde endocannabinoid signaling (3)
Tight junction (6)
Morphine addiction (10)
Rap1 signaling pathway (15)
Glycosaminoglycan biosynthesis (6)
Enrichment Score (-Log10 Q-Value)
Concordantly Regulated Pathways
Fatty acid metabolism (1)
Vascular smooth muscle contraction (9)
Biosynthesis of unsaturated fatty acids (2)
Enrichment Score (-Log10 Q-Value)
C
D
-Log10 (Q-value)
Moderate vs Mild COVID-19
Log2 (Fold Change)
5 MiRNA Down in Moderate COVID-19
miR-500b-5p
Concordantly Regulated Pathways
Wnt signaling pathway (31)
Adrenergic signaling in cardiomyocytes (35)
mTOR signaling pathway (20)
Calcium signaling pathway (42)
Biotin metabolism (1)
Hippo signaling pathway (35)
Thyroid hormone synthesis (20)
ErbB signaling pathway (25)
Thyroid hormone signaling pathway (34)
Axon guidance (34)
Enrichment Score (-Log10 Q-Value)
E
F
-Log10 (Q-value)
Severe vs Moderate COVID-19
Log2 (Fold Change)
540 MiRNA Down in Severe COVID-19
321 MiRNA Up in Severe COVID-19
miR-6745
miR-3934-5p
Concordantly Regulated Pathways
Arrhythmogenic right ventricular cardiomyopathy (62)
ErbB signaling pathway (76)
Thyroid hormone signaling pathway (102)
TGF-beta signaling pathway (70)
Glycosaminoglycan biosynthesis (15)
Axon guidance (107)
Hippo signaling pathway (133)
Mucin type O-Glycan biosynthesis (26)
Wnt signaling pathway (124)
Pathways regulating pluripotency of stem cells (124)
Enrichment Score (-Log10 Q-Value)
Concordantly Regulated Pathways
Ras signaling pathway (169)
Oxytocin signaling pathway (120)
ErbB signaling pathway (71)
Pathways regulating pluripotency of stem cells (104)
Dopaminergic synapse (102)
Circadian entrainment (78)
Glycosphingolipid biosynthesis (14)
Phosphatidylinositol signaling system (64)
Hippo signaling pathway (118)
Thyroid hormone signaling pathway (95)
Enrichment Score (-Log10 Q-Value)

**Online Figure XIV; Related to Figure 3 and 4:** Plasma MicroRNA Transcriptome Across the Disease Severity Subgroups. Volcano plots of differentially expressed miRNA between patient groups (a, c, e) with predicted KEGG terms (with enrichment score below and number of genes to the right) for pathways of deregulated microRNAs shown beside each corresponding region of the volcano plot (b, d, f). Data is displayed as FDR adjusted P values (Q values) vs the $\log_2$ fold change, with dashed lines are drawn to define restriction boundaries. See Supplementary Data Files III and IV for a full list of differentially expressed miRNA along with a full list of predicted KEGG pathways. Abbreviations: COVID-19 = Coronavirus Disease 2019; FDR = False discovery rate; KEGG = Kyoto Encyclopedia of Genes and Genomes; MiRNAs = MicroRNAs; SARS-CoV-2 = Severe acute respiratory syndrome coronavirus.

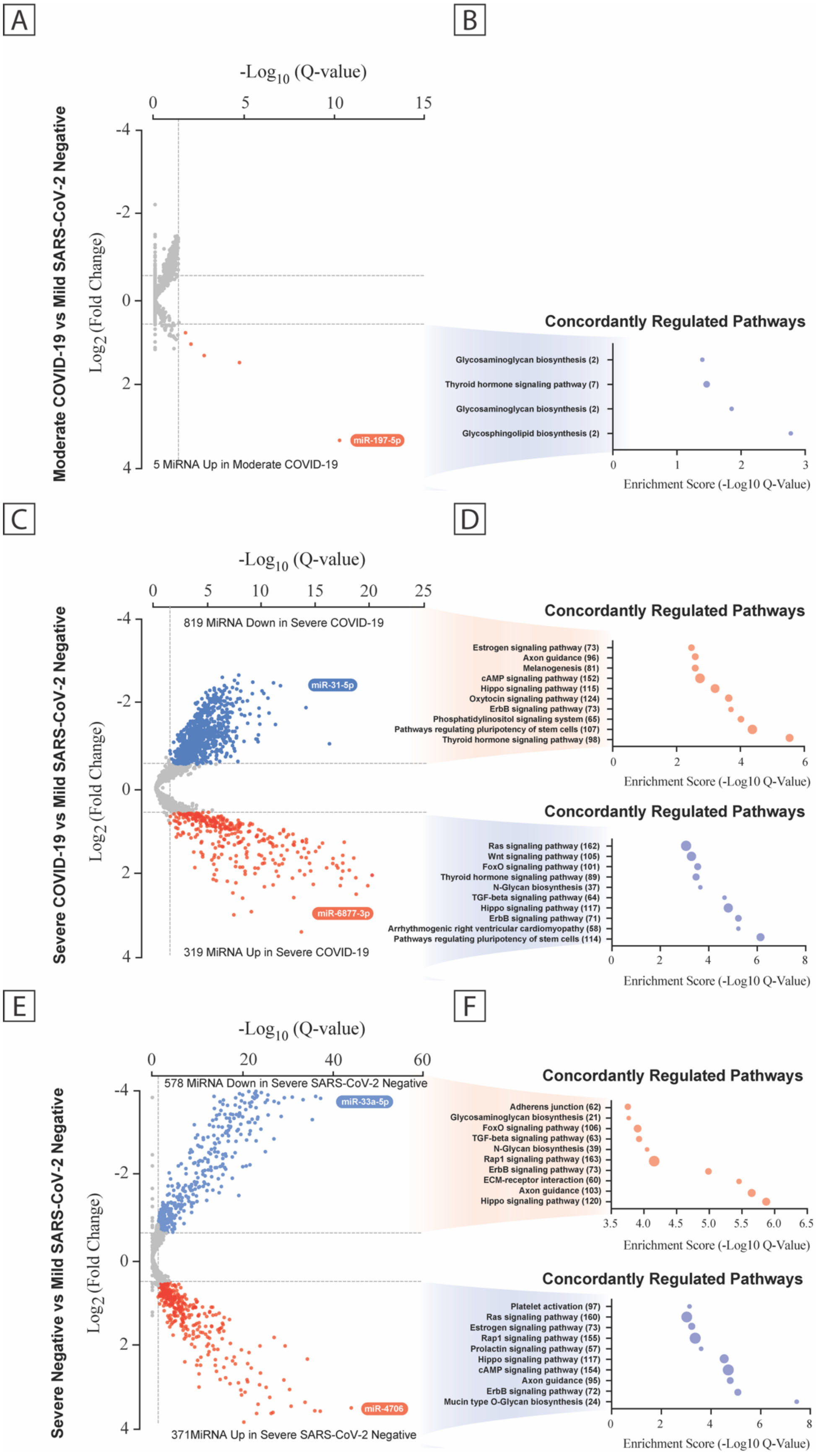
A
B
C
D
E
F
-Log10 (Q-value)
Log2 (Fold Change)
Moderate COVID-19 vs Mild SARS-CoV-2 Negative
miR-197-5p
5 MiRNA Up in Moderate COVID-19
Concordantly Regulated Pathways
Glycosaminoglycan biosynthesis (2)
Thyroid hormone signaling pathway (7)
Glycosaminoglycan biosynthesis (2)
Glycosphingolipid biosynthesis (2)
Enrichment Score (-Log10 Q-Value)
Severe COVID-19 vs Mild SARS-CoV-2 Negative
819 MiRNA Down in Severe COVID-19
miR-31-5p
miR-6877-3p
319 MiRNA Up in Severe COVID-19
Estrogen signaling pathway (73)
Axon guidance (96)
Melanogenesis (81)
cAMP signaling pathway (152)
Hippo signaling pathway (115)
Oxytocin signaling pathway (124)
ErbB signaling pathway (73)
Phosphatidylinositol signaling system (65)
Pathways regulating pluripotency of stem cells (107)
Thyroid hormone signaling pathway (98)
Ras signaling pathway (162)
Wnt signaling pathway (105)
FoxO signaling pathway (101)
Thyroid hormone signaling pathway (89)
N-Glycan biosynthesis (37)
TGF-beta signaling pathway (64)
Hippo signaling pathway (117)
ErbB signaling pathway (71)
Arrhythmogenic right ventricular cardiomyopathy (58)
Pathways regulating pluripotency of stem cells (114)
Severe Negative vs Mild SARS-CoV-2 Negative
578 MiRNA Down in Severe SARS-CoV-2 Negative
miR-33a-5p
miR-4706
371MiRNA Up in Severe SARS-CoV-2 Negative
Adherens junction (62)
Glycosaminoglycan biosynthesis (21)
FoxO signaling pathway (106)
TGF-beta signaling pathway (63)
N-Glycan biosynthesis (39)
Rap1 signaling pathway (163)
ErbB signaling pathway (73)
ECM-receptor interaction (60)
Axon guidance (103)
Hippo signaling pathway (120)
Platelet activation (97)
Ras signaling pathway (160)
Estrogen signaling pathway (73)
Rap1 signaling pathway (155)
Prolactin signaling pathway (57)
Hippo signaling pathway (117)
cAMP signaling pathway (154)
Axon guidance (95)
ErbB signaling pathway (72)
Mucin type O-Glycan biosynthesis (24)

**Online Figure XV; Related to Figure 3 and 4:** Plasma MicroRNA Transcriptome Across the Disease Severity Subgroups. Volcano plots of differentially expressed miRNA between patient groups (a, c, e) with predicted KEGG terms (with enrichment score below and number of genes to the right) for pathways of deregulated microRNAs shown beside each corresponding region of the volcano plot (b, d, f). Data is displayed as FDR adjusted P values (Q values) vs the $log_2$ fold change, with dashed lines are drawn to define restriction boundaries. See Supplementary Data Files III and IV for a full list of differentially expressed miRNA along with a full list of predicted KEGG pathways. Abbreviations: COVID-19 = Coronavirus Disease 2019; FDR = False discovery rate; KEGG = Kyoto Encyclopedia of Genes and Genomes; MiRNAs = MicroRNAs; SARS-CoV-2 = Severe acute respiratory syndrome coronavirus.

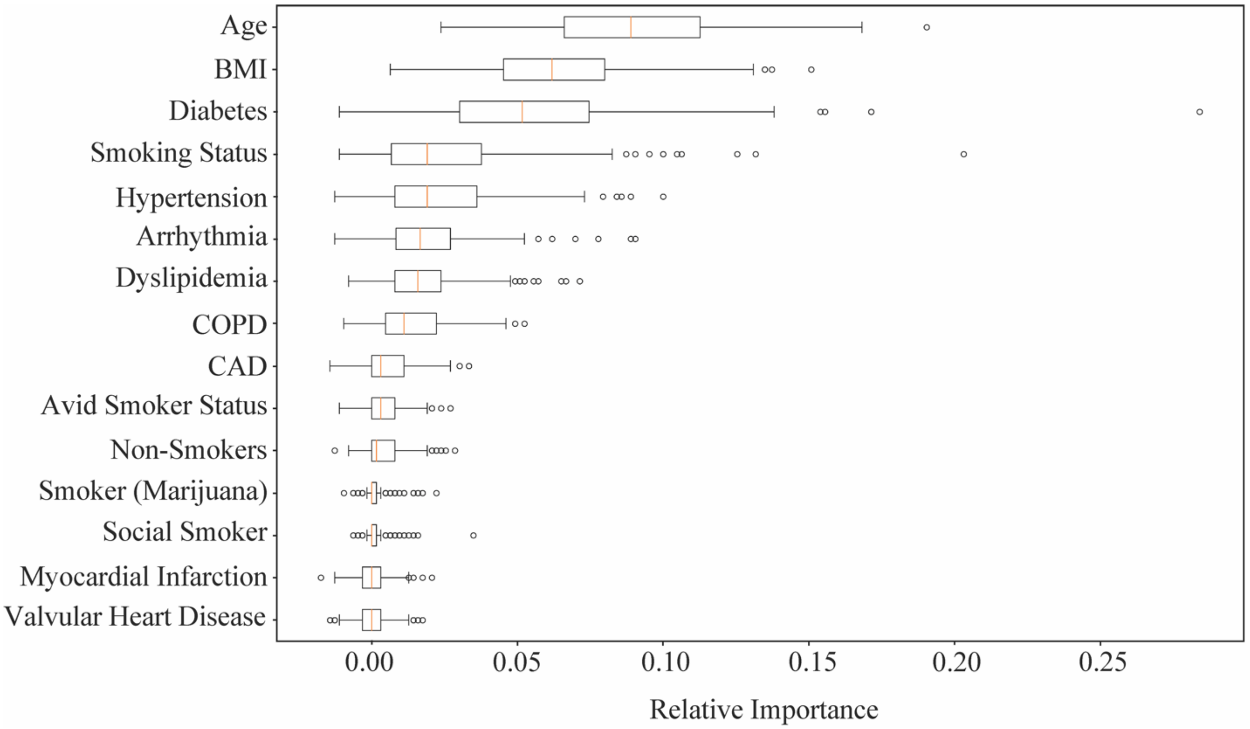

**Figure XVI; Related to Figure 4*:*** Feature importance of a machine learning model incorporating clinical data. All clinical metrics are at time of admission with preexisting conditions defined according to those listed in the methods. Abbreviations: BMI = Body mass index; CAD = Coronary Artery Disease; COPD = Chronic obstructive pulmonary disease.

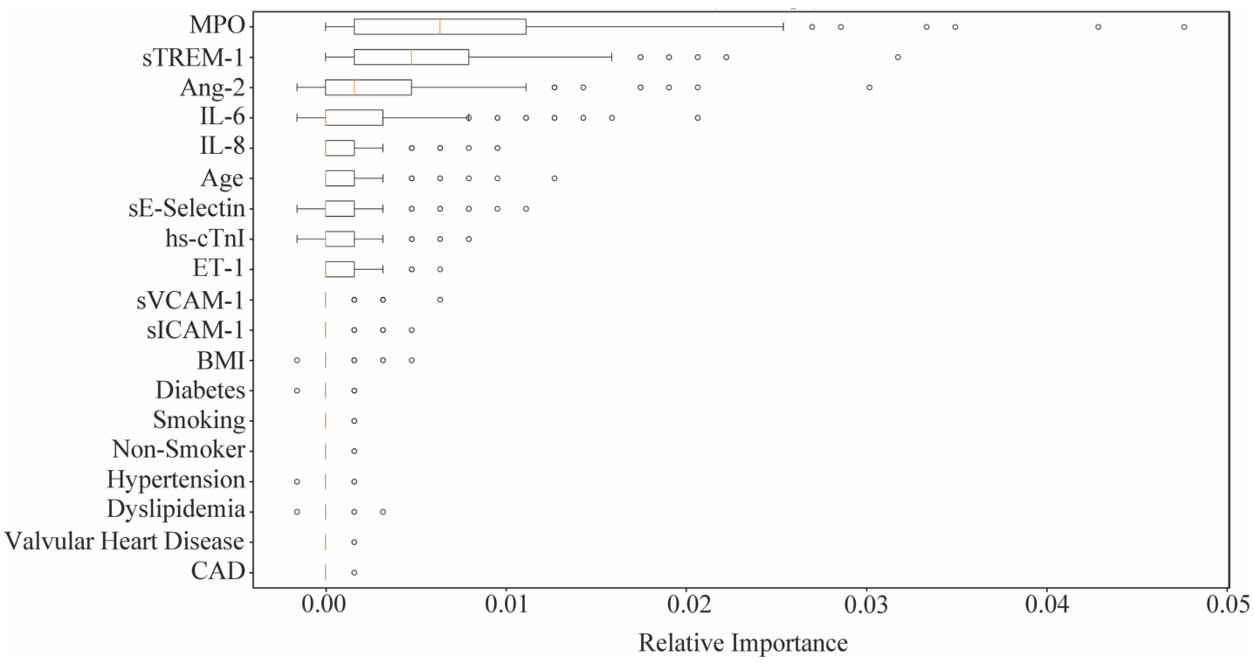

**Online Figure XVII; Related to Figure 4*:*** Feature importance of a machine learning model incorporating both clinical data and protein expression metrics. All clinical metrics are at time of admission with preexisting conditions defined according to those listed in the methods. Abbreviations: Ang-2 = Angiopoietin-2; BMI = Body mass index; CAD = Coronary Artery Disease; ET-1 = Endothelin-1; Hs-cTnI = High-sensitivity cardiac troponin I; IL = Interleukin; MPO = Myeloperoxidase; sICAM = Soluble intercellular adhesion molecule-1; sTREM-1 = Soluble triggering receptor expressed on myeloid cells-1; sVCAM-1 = Soluble vascular cell adhesion molecule-1.

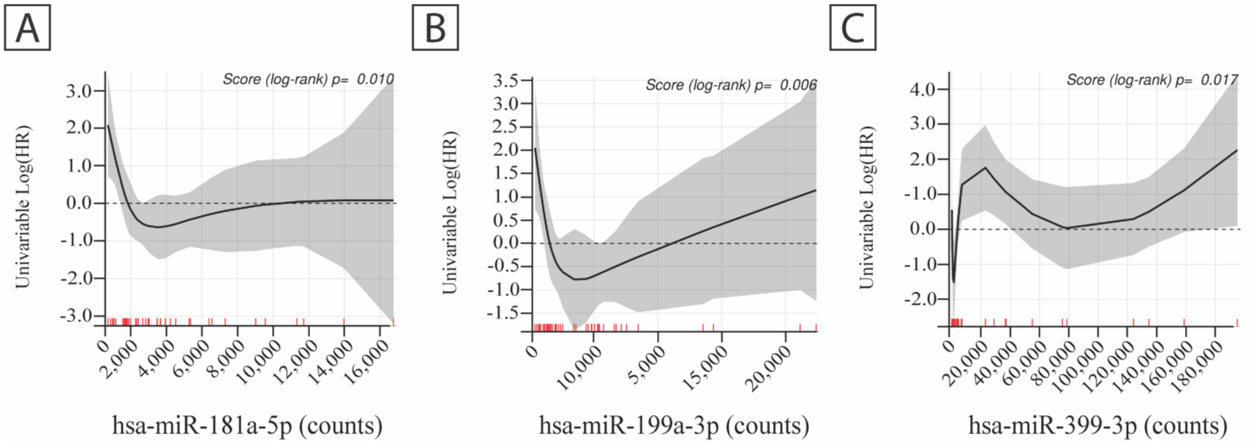

**Online Figure XVIII; Related to Figure 5: Association of Biomarkers with In-Hospital Mortality for Severe COVID-19 Patients.** Univariable log hazard ratios of candidate microRNAs (a) hsa-miR-181a-5p, (b) hsa-miR-199a-3p, and c) hsa-miR-339-3p.

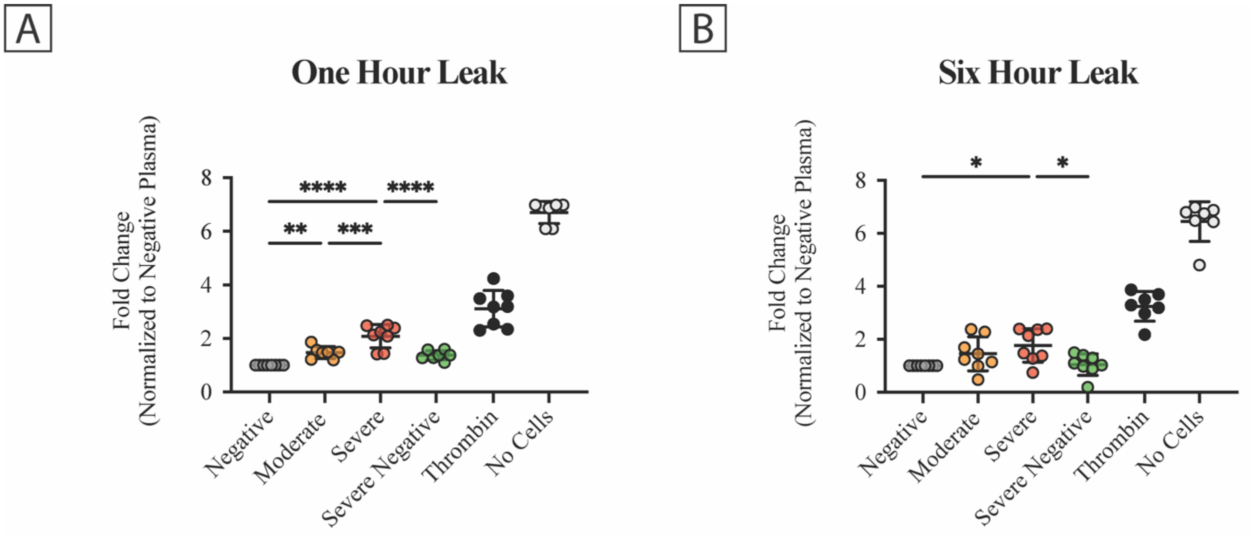

**Online Figure XIX; Related to Figure 5:** $T_{0-1}$ COVID-19 Patient Plasma Selectively Induces Acute Increases in Endothelial Permeability. (a) Permeability of pHUVEC monolayers was measured by 40 kDa FITC extravasation from the apical to the basolateral surface one-hour post-co-incubation. Treatment groups were normalized to the negative control. Center line represents the mean and error bars are (±S.D.). P values determined by one-way ANOVA test with Tukey's multiple comparisons test. Moderate vs negative: **$P=9.5\times10^{-3}$; Severe vs negative ****$P=5.6\times10^{-8}$; Severe vs moderate (P): ***$P=6.5\times10^{-4}$; Severe vs severe negative: ****$P=9.8\times10^{-5}$. (b) Permeability of pHUVEC monolayers was measured by 40 kDa FITC extravasation from the apical to the basolateral surface six hours post-co-incubation. Treatment groups were normalized to the negative control. Center bars represent the mean and error bars represent the standard deviation (±S.D.). P were values determined by one-way ANOVA test with Tukey's multiple comparisons test. Severe vs negative: *$P=2.1\times10^{-2}$; Severe vs severe negative: *$P=3.2\times10^{-2}$; n=7-8 per group. Thrombin treatment was included as a barrier disrupting positive control.

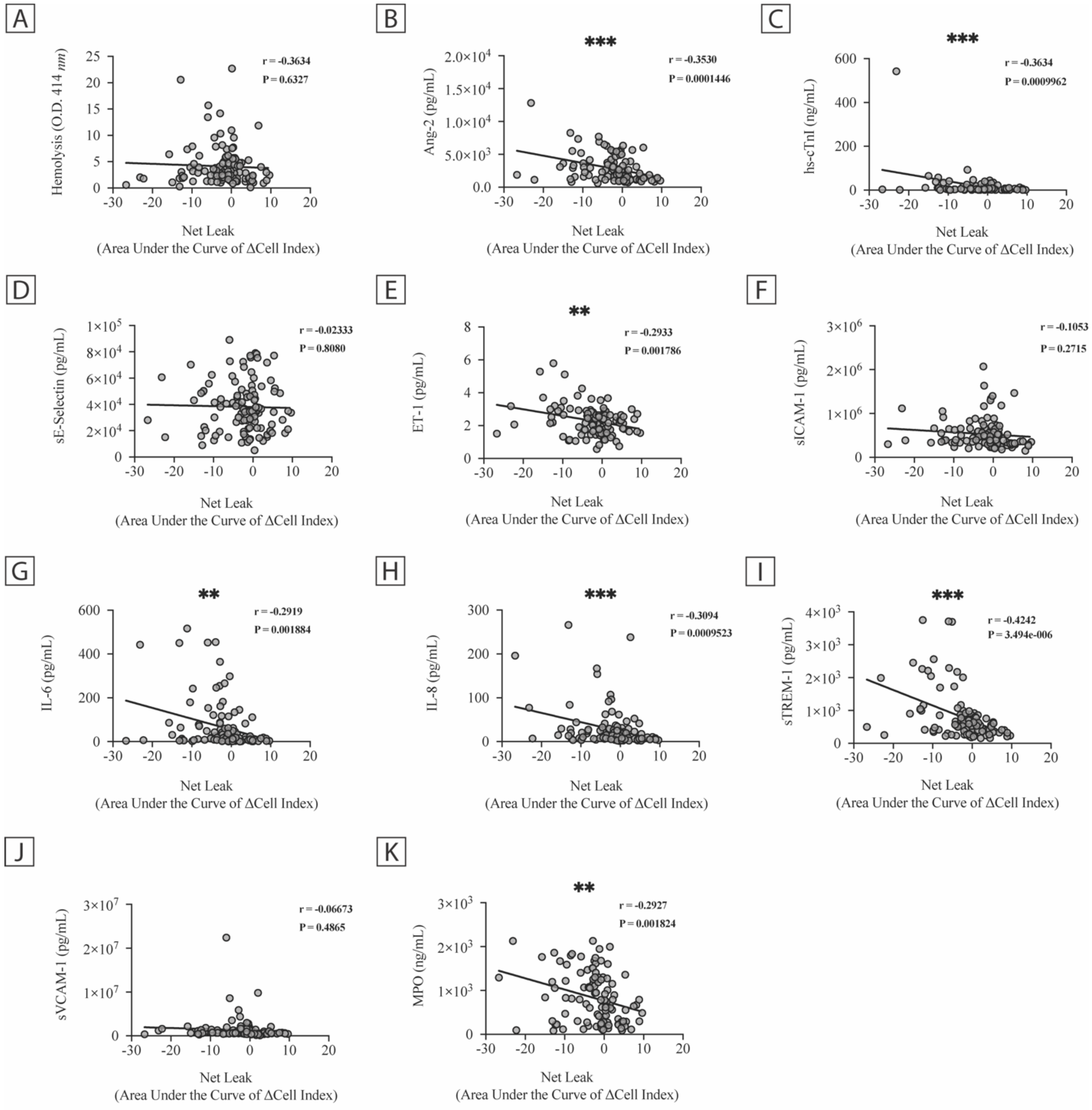

**Online Figure XX; Related to Figure 5:** Correlation of $t_{0-1}$ Plasma Cardiovascular Biomarkers in COVID-19 positive patients to Induction of Endothelial Permeability. Pearson correlations between (a) hemolysis, (b) Ang-2, (c) hs-cTnI, (d) sE-Selectin, (e) ET-1, (f) sICAM-1, (g) IL-6, (h) IL-8, (i) sTREM-1, (j) sVCAM-1, and (k) MPO to the change in pHUVEC TEER after six-hours co-incubation; n=111 per correlation. Leak is defined through negative values on the x-axis. Abbreviations: Ang-2 = Angiopoietin-2; ET-1 = Endothelin-1; Hs-cTnI = High-sensitivity cardiac troponin I; IL = Interleukin; MPO = Myeloperoxidase; sICAM = Soluble intercellular adhesion molecule-1; sTREM-1 = Soluble triggering receptor expressed on myeloid cells-1; sVCAM-1= Soluble vascular cell adhesion molecule-1.

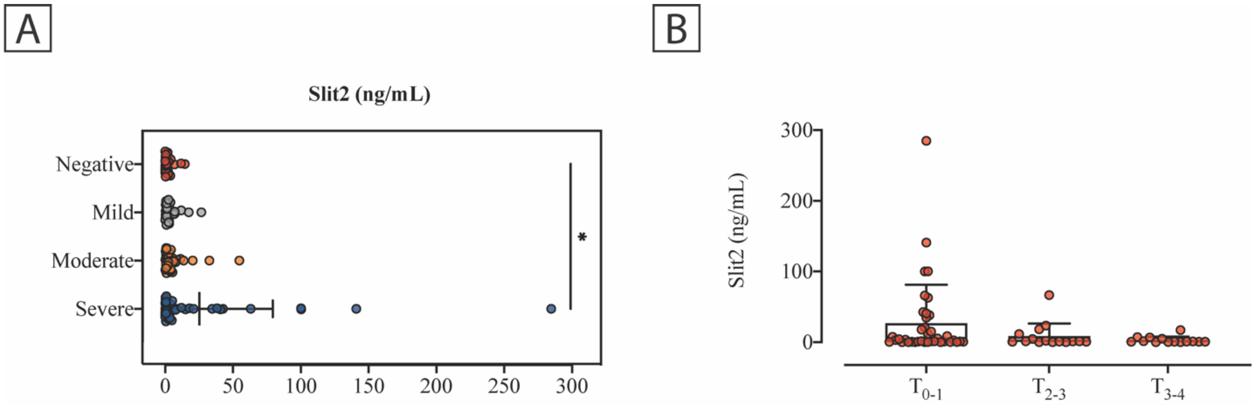

**Online Figure XXI; Related to Figure 6:** Endogenous sSlit2 is upregulated in severe COVID-19 patient plasma. (a) Endogenous sSlit2 at $t_{0-1}$ across the severity of COVID-19 (n=27-40, severe vs negative, *P=0.0279). (b) Endogenous sSlit2 at longitudinal intervals in patients with severe COVID-19 (n=14-38). Center bars represent the mean and error bars represent the standard deviation (±S.D.). P values were determined by one-way ANOVA test with Tukey's multiple comparisons test.

### SUPPLEMENTAL DATA FILE ANNOTATIONS

**Supplementary Data File I.** Quality control table for all RNA-sequencing experiments used in this study.

**Supplementary Data File II.** R documentation file for the analysis of the RNA-sequencing experiments.

**Supplementary Data File III.** R code for the analysis of the RNA-sequencing experiments.

**Supplementary Data File IV.** Full list of differentially expressed miRNA with pairwise comparisons between COVID-19 cohorts and the negative controls.

**Supplementary Data File V.** Full list of pathway enrichments for miRNA-sequencing experiment between COVID-19 cohorts and the negative controls.

**Supplementary Data File VI.** Full list of differentially expressed genes with pairwise comparisons between COVID-19 cohorts and the negative controls.

**Supplementary Data File VII.** Full list of pathway enrichments for mRNA-sequencing experiment between COVID-19 cohorts and the negative controls.

**Supplementary Data File VIII.** Gene set enrichment analysis for mRNA-sequencing experiment between COVID-19 cohorts and the negative controls.
